# Effects of concurrent transcranial direct and alternating current stimulation on human motor skill learning: a systematic review and Bayesian meta-analysis

**DOI:** 10.1101/2025.11.19.25340588

**Authors:** August Lomholt Nielsen, Maria Isbrandt Gad, Lasse Jespersen, Jonas Rud Bjørndal, Mikkel Malling Beck, Lasse Christiansen, Anke Ninja Karabanov, Jesper Lundbye-Jensen

**Author notes:** Correspondence: August Lomholt Nielsen, Movement and Neuroscience, Department of Nutrition, Exercise and Sports, University of Copenhagen, Nørre Allé 51, DK2200 Copenhagen, Denmark.

## Abstract

Motor learning is important throughout human lifespan for acquisition, refinement and preservation of motor skills and for regaining functional abilities through rehabilitation training. Therefore, it is essential to develop strategies and paradigms to facilitate processes of motor learning. Weak-current transcranial electrical stimulation (TES) can be applied with the aim of augmenting effects of concurrent motor practice. Two dominant application modalities of TES, transcranial direct or alternating current stimulation (tDCS and tACS), can modulate and entrain endogenous neural activity, respectively. Here, we investigate effects of tDCS and tACS applied during motor practice on core features of motor skill learning, namely speed and accuracy, based on seventy-three studies employing a broad variety of motor tasks. We used Bayesian hierarchical random-effects modeling to estimate pooled effects and explore potential moderators, such as number of sessions, stimulation intensity, and outcome measure. We found medium positive effects of tDCS applied to target the motor cortex (n=48), cerebellum (n=16), or other cortical areas (n=17, mainly the prefrontal cortex or parietal cortex) on increases in speed and accuracy during motor practice, with smaller effects observed for consolidation between sessions. Multiple practice sessions with concurrent tDCS demonstrated favorable effects. In contrast, tACS yielded negligible effects. In conclusion, we provide evidence of positive effects of applying tDCS but not tACS during motor practice to enhance practice-related improvements and retention of motor skill, speed and accuracy. The accumulation of positive effects with repeated exposure, as well as cross-domain effects, supports the use of tDCS in real-world applications, including neurorehabilitation.

**Highlights:**

- *tDCS enhances within-session motor learning, rather than between-session consolidation*
- *Multi-session tDCS yields greater motor learning than single-session interventions*
- *tDCS effects were not linked to target area, intensity, or task characteristics*
- *tACS reveals no significant effects on motor learning at any tested frequency*

## 1 | Introduction

The ability to learn new motor skills underpins many aspects of human function, ranging from everyday tasks to athletic performance and neurorehabilitation. Behavioral improvements driven by repeated motor practice depend on changes within the central nervous system that involve the formation of new neural connections and reorganization of neural circuits [1], [2]. Motor learning refers to the process of acquiring new motor skills or enhancing existing ones, and is typically characterized by improvements in movement accuracy and execution speed [3]. Conceptually, motor skill learning involves identifying an appropriate movement goal, given specific constraints, selecting the correct motor plan to achieve that goal, and executing the movement with increasing precision [4]. Efforts to optimize motor skill learning have led to the exploration of neuromodulatory interventions aimed at promoting neural plasticity. Among these, transcranial electrical stimulation (TES) has emerged as a promising non-invasive technique to modulate brain activity and support motor learning and control [5], [6]. The literature suggests that TES can enhance motor function and facilitate motor learning in healthy individuals, with potential applications in injury prevention and rehabilitation [7]. Among the TES modalities, Transcranial Direct Current Stimulation (tDCS) [8] and Transcranial Alternating (sinusoidal) Current Stimulation (tACS) [9] have received significant attention because of their ease of use and low cost. A wide range of motor learning paradigms have been employed in laboratory settings to investigate the behavioral effects of tDCS/tACS, including tasks focused on motor adaptation, sequence learning, de novo skill acquisition, and motor acuity [10], [11], [12], [13].

A central aspect of motor learning is the trade-off between movement speed and accuracy, which is often examined by fixing one variable while measuring changes in the other, or by modeling their interaction explicitly. This is particularly relevant in TES studies, where the stimulation effects may depend on the specific motor task and experimental parameters. The sequential finger tapping task (SFTT) and sequential visual isometric pinch force task (SVIPT) are among the most frequently used paradigms for experimentally investigating motor skill learning [14]. Using these paradigms, TES has been found to shift the speed-accuracy trade-off curve or enhance performance in one dimension [15], [16], [17].

Motor learning unfolds over multiple temporal stages, typically classified as *online learning* (performance gains during active practice) and *offline learning*, which refers to improvements or consolidation occurring between sessions [18]. While most TES studies have focused on single-session designs, growing interest has emerged in the effects of repeated practice combined with multiple TES sessions [10], [19], [20], [21]. This is particularly relevant from a practical perspective because acquiring and mastering new motor skills typically requires repeated exposure and extended practice. Evidence suggests that TES may enhance neural mechanisms involved in online [22] as well as offline learning [10], [23].

The neural mechanisms underlying motor skill learning involve dynamic interactions across multiple brain regions, which undergo functional reorganization in response to practice and experience [18], [24]. These plastic changes depend on the temporal and spatial coordination of neural firing, which serves as a prerequisite for strengthening synaptic connections through well-established mechanisms such as long-term potentiation (LTP) and long-term depression (LTD) processes [1], [2] ultimately contributing to the refinement and optimization of motor programs. Despite extensive research, the precise mechanisms by which TES modulates neural activity and learning remain incompletely understood [5]. The effects of TES are influenced by multiple interacting factors, including non-uniform stimulation across cortical areas, individual differences in brain anatomy (e.g., gyrification), the functional brain state at the time of stimulation, and concurrent neuroinflammatory or neuromodulatory processes [25], [26]. As such long-standing notion that anodal stimulation facilitates, whereas cathodal stimulation inhibits, the cortical region underneath the electrode does not encompass the complexity of the effects on neural activity beneath and between the stimulation electrodes [27], [28]. The primary motor cortex (M1) has been the most frequently targeted region in TES studies, likely because of its central role in motor skill acquisition and consolidation [6]. Other studies have explored the effects of stimulation of the Cerebellum (CB) and higher cortical areas, such as the prefrontal, premotor, and posterior parietal cortices, to influence specific processes involved in motor learning [29], [30].

Despite the substantial body of research on tDCS and tACS, the need for a comprehensive and updated meta-analysis has become evident owing to the publication of new studies and the evolution of research methodologies. Our meta-analysis aimed to address several key issues. First, we included recent studies that have emerged since previous meta-analyses, thereby providing a more comprehensive evaluation of the effects of tDCS and tACS on motor learning. Importantly, we focused exclusively on studies that employed a motor learning paradigm, ensuring that the interventions were assessed within the context of both skill acquisition (online learning) and retention (offline learning) rather than merely motor performance. Secondly, to isolate the direct effects of brain stimulation on motor learning, we included only those studies in which stimulation was applied concurrently with motor practice. This criterion helps to exclude potential homeostatic mechanistic influences that might arise if stimulation is applied prior to or after practice sessions. Third, we ensured that the motor learning paradigm encompasses both accuracy and speed, given that both components are critical to understanding the full impact of tDCS and tACS on motor skill learning. Finally, our meta-analysis employed Bayesian statistical methods, which offer a robust framework for quantifying the effects of tDCS and tACS, as they allow the integration of prior knowledge with observed data and explicitly model between-study variability. This approach is particularly well suited for synthesizing evidence from heterogeneous study designs and capturing the uncertainty inherent in complex motor learning paradigms.

Specifically, it addresses the following questions: What are the effects of concurrent tDCS/tACS on motor skill learning in able-bodied young adults? How do specific stimulation parameters, such as electrode placement applied within tDCS/tACS protocols, influence motor skill learning when stimulation is applied concurrently with motor practice? Do the observations and quantitative assessments across studies allow recommendations for stimulation protocols to promote learning through effects on acquisition within-session or between-session and retention?

## 2 | Method

### 2.1 | Protocol and registration

The systematic review process was conducted using Covidence (covidence.org). The procedure commenced with the importation of studies in Covidence, initiating the systematic review protocol. The first phase involved screening titles and abstracts. Studies that met the predefined inclusion and exclusion criteria during the initial screening were then subjected to a comprehensive full-text review. Subsequently, data were extracted from studies that qualified for inclusion. For detailed information on the data extraction process, please refer to Section 2.5.

This systematic review was conducted following the PRISMA guidelines (PRISMA [prisma-statement.org]) [31]. The literature search was performed in November 2023, and the data extraction phase was completed in April 2024. The registration protocol for this review can be accessed at: https://www.crd.york.ac.uk/PROSPERO/display_record.php?RecordID=515752, ID: CRD42024515752.

### 2.2 | Literature search

The literature search was conducted through multiple public databases. PubMed, Web of Science, Scopus, SPORTDicus, CINAHL, Cochrane Central Register of Controlled Trials (CENTRAL), and MEDLINE were scanned. To encompass all studies of interest, a nuanced search strategy was developed, focusing on the type of intervention, namely, tDCS and tACS, and the primary outcome of the intervention, which was motor skill learning.

Searching in the electronic databases with various transcranial electrical stimulation terms was used using the Boolean operator “OR.” These terms encompassed [tDCS], [transcranial direct current stimulation], [anodal transcranial direct current stimulation], [transcranial electric stimulation], [tACS], [anodal transcranial alternating current stimulation], or [transcranial alternating current stimulation]. To further ensure a comprehensive focal point of motor skill learning, phrases like [motor skill learning], [motor learning], [motor practice], [motor skill acquisition], [motor acquisition], [motor sequence], [visuomotor], [learning curve], [sequence learning], and [serial learning] were employed using the Boolean operator “OR.” These two distinct focus points were paired using the Boolean operator “AND” to create a search matrix that integrated nuances of TES and motor learning.

### 2.3 | Eligibility criteria

Studies were selected according to predefined inclusion and exclusion criteria. To be eligible for inclusion, studies had to meet the following conditions: I) they investigated the effects of anodal tDCS or tACS against sham tDCS/ACS (placebo stimulation) in young healthy adults on motor learning. This decision reflects the substantial predominance of anodal electrode placement near the target area (anodal tDCS) and the cathode at a reference area in motor learning research [6]; II) tDCS/ACS had to be applied concurrently with the motor skill training session(s) alone, meaning that no other forms of TES were combined with it; III) the study had to explicitly investigate the change in the speed-accuracy trade-off either by concurrently evaluating both speed and accuracy metrics or systematically tracking one parameter while holding the other constant (described further in next section); IV) eligible study designs included randomized controlled trials (RCTs), crossover RCTs, and non-randomized controlled trials, regardless of blinding level (single-, double-, triple-, or unblinded).

Data from interventions were excluded if: I) the stimulation was not applied concurrently with motor practice; II) the intervention comprised mental training, motor imagery, and motor observational training; III) the outcome measures did not evaluate the behavioral effects of stimulation (real/sham) and concurrent motor practice; IV) the intervention lacked adequate methodological description of the outcome measures evaluated; V) the study lacked a control group(s); VI) the study disregarded one component of the speed–accuracy trade-off in their outcome assessment; and VII) the intervention combined stimulation methods other than tDCS and tACS.

#### 2.3.1 | Selection of outcome measures

In this review, the term “motor skill learning” was defined as a shift in the speed-accuracy trade-off. This definition served as a foundation for establishing the inclusion criteria for the selection of studies as listed in the previous section. This encompassed a broad spectrum of motor learning paradigms, but specifically targeted those demonstrating quantifiable changes in motor performance, characterized by either a heightened precision in task execution or a reduction in the time required to reach a specified level of accuracy. Only studies that disclosed information about the relationship between temporal and spatial behavioral changes were included (i.e., speed-accuracy tradeoff). This means that studies that only provide information about e.g., correct responses and no additional information about the temporal dynamics of the task were excluded. This includes studies that discarded data to analyze only one of the two components without further argumentation. The basis of this choice was to investigate the change in either speed, accuracy, or both, and to ensure that performance improvements in one of the parameters are not a result of lowering the other parameter. The included tasks spanned surgical tasks, serial visual isometric pinch tasks (SVIPT), finger tapping tasks, serial reaction time tasks (SRTT), and the Purdue pegboard test. The primary outcomes were movement time, completion time, absolute error, and relative error. The inclusion criteria for the studies in this review encompassed both learning paradigms involving varying degrees of motor demands, considering temporal and spatial dimensions. The motor demands were categorized into two main groups: those with greater motor demands, which pertain to the quality of movement execution during the task itself, and those with minor motor demands, which encompass tasks that place equal emphasis on motor execution as well as cognitive and perceptual dimensions. An illustrative example of a task that falls into the category of minor motor demands is the SRTT, wherein participants selectively press a key in response to a stimulus in a spatially concurrent manner. While it is recognized that the SRTT has some limitations, such as not precisely determining motor action acuity, it was included in this review because of its significance in understanding action selection and the explicit fragmented learning observed in SRTT. However, it is important to approach the results related to SRTT with careful consideration and caution because of the poor determination of precise motor execution, but rather inhibitory control [4]. Tasks involving adaptation as a learning paradigm were excluded from the review. Such tasks often involve individuals adjusting to perturbations or disruptions in their motor execution environment (e.g., manipulating objects with altered dynamics or navigating through visually distorted fields). While these tasks involve motor behavior changes, according to our definition, they primarily assess an individual’s capacity to adapt to new conditions rather than their ability to acquire or refine motor skills [4].

### 2.4 | Study selection

Two independent reviewers, M. I. G. and A. L. N., screened the titles, abstracts, and subsequently, the full texts. In the event of disagreement between reviewers, a third reviewer (L. J.) was consulted to assess contentious studies. The researchers M. I. G. and A. L. N. read the selected studies twice to enhance familiarity with the data before extracting and synthesizing the findings.

### 2.5 | Data extraction, analysis, and synthesis

Reviewers M. I. G. and A. L. N. independently extracted data from each study using a predefined Excel spreadsheet. This spreadsheet was coded for variables such as sample size, participant characteristics (sex and age), tDCS/ACS stimulation protocol details, exercise protocol, number of sessions required by the study, and performance outcomes (improvement/no improvement), based on the recommendations of Popay et al. (2006) [32] (see Table S1). The extracted effect sizes (see section 2.7) were divided into online and offline learning effects. Online learning was defined as the performance change from baseline to a time point within half an hour after motor practice, reflecting immediate learning effects. Offline learning refers to performance changes occurring beyond three hours after practice, including delayed retention tests. While most studies assessed offline effects after overnight sleep (≥24 hours post-practice), one study evaluated retention at a 3-hour delay, which was also categorized as offline based on established criteria for delayed consolidation [33]. Studies investigating the effects of multiple training sessions combined with stimulation have been conducted over several days. When data permitted, online learning was defined as the accumulated sum of within-session gains across sessions or, alternatively, as the total learning gain from baseline to post-practice. For offline learning, between-session gains were similarly aggregated across sessions, defined as either the cumulative improvement between sessions or the difference between post-practice and retention test performance. Studies were categorized and ordered according to the task paradigm (See Table S1). If values were not explicitly stated in the studies, they were extracted using Webplot digitizer Version 4.4 (https://apps.automeris.io/wpd/). Data extraction was performed twice to ensure accuracy and reduce bias. M. I. G. and A. L. N. resolved any ambiguities through discussions.

### 2.6 | Evaluation of bias risk

Both M.I.G and A.L.N, independently evaluated study biases based on the Cochrane manual’s criteria [34]. They systematically assessed key domains of bias, including selection, performance, detection, attrition, and reporting. Each domain was evaluated and rated as having a low, unclear, or high risk of bias based on the quality and transparency of its methodological reporting. Any disagreements between the authors were resolved through discussion until mutual agreement was reached. This evaluation was applied consistently during all search updates. The risk of bias was described as follows: 1) random sequence generation (selection bias); 2) allocation concealment (selection bias); 3) blinding of participants and personnel (performance bias); 4) blinding of outcome assessment (detection bias); 5) incomplete outcome data (attrition bias); and 6) selective reporting (reporting bias). The Cochrane quality assessment can be found in the Supplementary Material (S2).

### 2.7 | Statistical analyses

The extracted data were synthesized using a meta-analytic approach based on Bayesian statistics. The data are presented as estimated effect sizes from the Bayesian posterior distribution, highlighting the key findings and trends across the included studies. Furthermore, we employed the specific framework proposed by Ince et al. (2021) [35], which incorporates Bayesian prevalence to elucidate the perspective of statistical prevalence. The method evaluates the observed effects and assesses the likelihood of obtaining similar results if the experiment is replicated. The motivation for including Bayesian prevalence is that it is possible to quantify how typical or uncommon an observed effect is and the uncertainty around this estimate [35].

In each article, the size of the intervention effect was calculated according to the difference in performance outcomes between experimental and control conditions. The intervention effect was measured by calculating the standardized mean difference (SMD) of continuous data. The results are reported with 95% confidence intervals (95% CI). A separate meta-analysis was performed for each subgroup to investigate the broader effects across various domains, including online and offline learning, stimulation target areas, and the number of tDCS/ACS sessions. This analysis was conducted carefully considering the diverse outcome measures within each study, such as speed, accuracy, and skill assessment, and modeled as a random effect (see section 2.7.1). Owing to the diverse populations and varying tasks utilized in the studies, a random effects model was employed to account for statistical heterogeneity and inter-study dependencies more accurately [34], [36]. Cochrane guidelines report standardized mean difference (SDM) using Cohens Effect Size to represent small (≤0.2), moderate (≤0.5) large (≤0.8), and very large (>0.8) effect sizes [34], [37] (available at: http://handbook.cochrane.org) [34], [37]. Coheńs (1988) [37] general guidelines for interpreting effect sizes can serve as a benchmark; however, these guidelines may undervalue the significance of small or medium effect sizes that possess practical or clinical relevance within a specific research domain [38]. An examination of prior relevant research provides some indication of the magnitude obtained in previous studies on different types of outcomes that can guide current findings to be placed in an appropriate context. No empirical analysis has been conducted in this research area to establish what constitutes small, medium, or large effects in this context. In other research areas, it has been demonstrated that Coheńs effect size categorization system is largely inappropriate because it is not based on the actual distribution of effect sizes reported in the literature [39]. An empirical distribution goes beyond the purpose of this paper, but we discuss the Bayesian estimated effect sizes based on previous relevant effect sizes and consider a threshold of 0.3 to be notably meaningful, and values below are considered less meaningful.

#### 2.7.1 | Meta-analytic model

A random-effects model was employed using a Bayesian multilevel hierarchical structure to provide a pooled estimate of the overall effect size by synthesizing the weighted results from the included studies. This model accommodates the estimation of an average effect from heterogeneous studies, enabling the derivation of more generalizable inferences while adequately accounting for inherent between-study variability and potential outliers in the data [40], [41], [42], [43], [44]. The multilevel structure of the random-effects model accommodates variations due to differences in study design or population, thereby explicitly modeling heterogeneity in the results. The credible interval for the pooled estimate incorporates both within-study sampling error and between-study variance, thus providing a comprehensive measure of uncertainty [45]. In this specific case, we employed a random-effects model with a nested structure: multiple effect sizes (level 2) were nested within individual studies (level 3). This hierarchical approach allowed us to estimate two sources of variance, the variance between effect sizes within a study (level 2) and the variance between studies (level 3).

To estimate the pooled effect size and its credible intervals, a hierarchical Bayesian robust regression model was fitted to the data using a Student’s t-likelihood. This model consists of multiple levels, allowing for the incorporation of the correlation among multiple within-study effect sizes, which prevents the underestimation of uncertainty [42]. The use of a Student’s t-distribution in the robust regression model mitigates the impact of outliers, which could otherwise disproportionately influence the results [46]. Our prior beliefs were modeled as weakly informative priors, meaning that the influence of the priors on the posterior probability density was less weighted in the model but weighted toward the likelihood [44]. Extreme effect sizes were discounted as unlikely. The commonly used standardized scale, for example, Cohen’s d, assumes the *d* scale for effect sizes, and between-study variance > 0:25 and population average > 1:0 are unlikely to occur [37]. A mean of 0 and a standard deviation of 0.5 were chosen for the overall effect. This prior implies that we grant an approximation of a 95% prior probability that the true estimated effect size lies between -2 and 2. We set the prior heterogeneity of the effects (i.e., the standard deviation of random effects) with a mean of 0 and a standard deviation of 0.5. For the degrees of freedom parameter ν of the Student’s t-distribution, a gamma distribution with a shape parameter of 2 and a scale parameter of 0.1 was chosen to ensure that the model remains robust to the influence of outliers [41]. Prior predictive checks were conducted to verify that the model’s predictions for plausible effect size values excluded only implausibly extreme values based on the specified priors [47]. The models were fitted using Hamiltonian Monte Carlo samplers with 4 parallel chains with 20,000 iterations each, 200 warm-up samples, and an adapt delta of 0.99. Model validation was assessed by 1) performing prior and posterior predictive checks, 2) ensuring Rhat statistics to be close to 1, 3) plotting prior against posterior estimates and assessing whether the posteriors had lower variance than the priors, 4) ensuring no divergences in the process of estimation, and 5) checking that the number of effective bulk and tail samples was above 200. Leave-one-out (loo) information criteria and stacking weights were calculated to determine which model exhibited the lowest pointwise out-of-sample prediction error and was generalized most effectively to new data [48].

A separate model was constructed to explore the potential influence of the number of tDCS sessions as a moderator. By including the number of sessions as a covariate, the model was designed to isolate and evaluate its impact on the overall effect size, thereby providing insights into whether the frequency of tDCS sessions plays a significant role in the observed outcomes (see Section 3.7).

In addition, separate models were developed to incorporate outcome measures, task types, target area (model for other higher cortical areas), and stimulation intensities as moderators. These models investigated how different outcome measures and task or stimulation characteristics employed in each study influenced the estimated effect, allowing for a more nuanced understanding of how specific study parameters affect the overall efficacy of tDCS in motor skill learning.

A separate analysis was conducted to evaluate the publication bias of the included studies. The framework provided by Mathur and VanderWeele (2020) [49] includes a quantitative sensitivity analysis and inspection of a significance funnel plot, which were used to assess the publication bias. Selected studies are considered as a representation of samples from an underlying population of published and unpublished studies, where the probability of selection for significant studies is higher [49]. The potential presence of publication bias is assessed by estimating the degree of severity of publication bias required to attenuate the point estimate or credible interval to the null. Publication bias analysis was only carried out for tDCS studies due to the low number of included studies considering tACS. All statistical evaluations were performed using R 4.3.2 (R Core Team, 2023). The R package “metafor” [50] was used to estimate within-study effect sizes and to compute SMD using a frequentist approach, serving as a reference point for Bayesian inference. For the Bayesian analysis, we used the R package “brms” [51] to estimate posterior distributions via Hamiltonian Monte Carlo implemented in Stan via Stan Development Team (R package version 2.26.24 - https://mc-stan.org/) [52], using RStudio 4.2 (RStudio Team, 2024).

An additional analysis was conducted to estimate the population-level prevalence of statistically significant effects, where we applied a Bayesian prevalence method (see Supplementary Material, S8) inspired by [35]. Briefly, each study contributed a binary outcome (significant versus non-significant) under the standard NHST framework. We modeled the number of significant studies with a binomial likelihood and a uniform Beta(1,1) prior, yielding a posterior from which we extracted the maximum a posteriori (MAP) estimate of the true-positive prevalence. Under this prior, the MAP coincides with the sample proportion. We also computed 96% highest posterior density intervals (HPDIs) to capture uncertainty in the prevalence estimate. Unlike frequentist confidence intervals, HPDIs directly reflect the most credible prevalence values, given the observed data.

## 3 | Results

### 3.1 | Overview

The database search process produced 2110 unique records for screening, which resulted in 314 full-text records that were assessed for eligibility (Fig. 1). After the final study selection, 73 studies were included, of which 64 investigated the effects of tDCS and 9 investigated the effects of tACS on motor learning. An overview of the included studies and their characteristics (stimulation type and target area) is shown in Fig. S1. As is evident from Figure S1, an increasing number of studies have been published since 2009.

**Figure 1:**
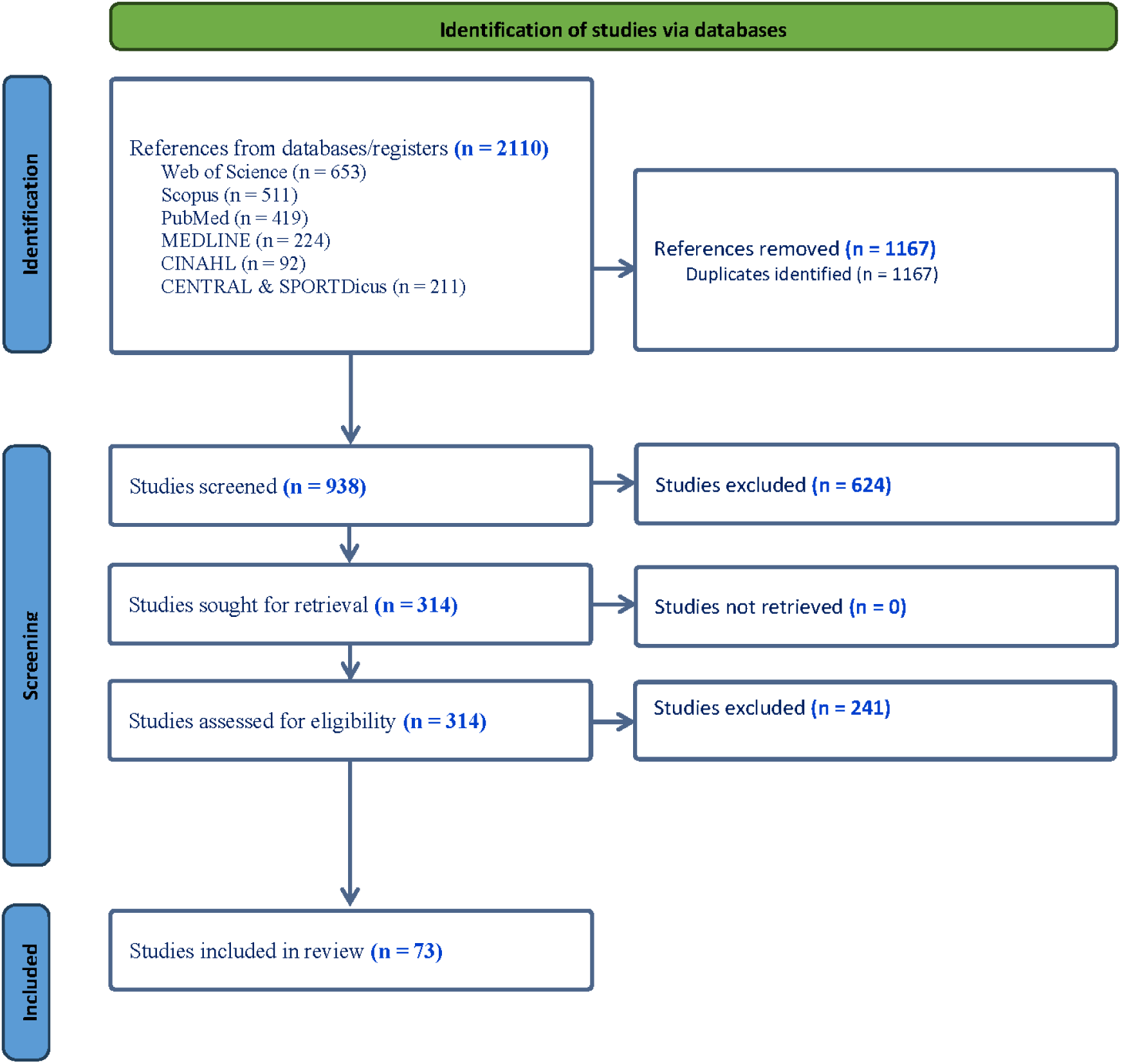
Flowchart of the included studies.

### 3.2 | Study characteristics

Table S4-7 provides an overview of the study characteristics, including study information, stimulation protocol, and motor skill learning procedures. The sample comprises 73 articles published between 2009 and 2023, with a notable increase in publication rate since 2009 (Fig. S1). The studies exhibit considerable variability in terms of TES specifications and motor learning protocols. Among the included studies, 64 employed tDCS and 9 utilized tACS. Specifically, 91 tDCS intervention groups were compared to control groups, and 20 tACS intervention groups were similarly compared to a control group. Within the tDCS domain, 57 groups targeted the M1, 16 groups focused on the CB, and another 17 groups applied tDCS to other higher cortical areas (HCA), such as the supplementary motor area (SMA), dorsolateral prefrontal cortex (DLPFC), and posterior parietal cortex (PPC). One group received tDCS applied to M1 and CB simultaneously. The majority of studies employed a stimulation intensity of 1-2 mA, with 26 and 28 groups receiving 1 and 2 mA, respectively; however, the intensities ranged from 0.3 mA to 4 mA. The electrode materials varied, but all the studies utilized carbon rubber electrodes. Specifically, 51 studies used sponges with saline solution as the contact medium, and 13 studies used conductive gel (e.g., Ten20 conductive paste). Electrode placement was predominantly based on the international 10-20 EEG system (29 studies) or the Transcranial Magnetic Stimulation (TMS) hotspot (17 studies) in studies using M1 as target area. Four studies utilized both the 10-20 EEG system and the TMS hotspot, while two studies used MRI coordinates for electrode placement. A total of 14 different devices were used across the included studies. The NeuroConn DC stimulator (NeuroConn, Ilmenau, Germany) was the most commonly used, followed by the Soterix 1X1 Low-Intensity Smart ScanTM tDCS device (SOTERIX MEDICAL INC), Stimulator (Oasis Pro of Mind-Alive Inc. model, Edmonton, AB, Canada), Phoresor II Auto - model PM850 Iomed, tDCS Brain Stimulation Device – ApeX Type A USA Model China, Caputron ActivaTrek’s “ActivaDose 2”, Intelect Advanced Therapy System, Starstim system (Neuroelectrics, Barcelona, Spain), Magstim Company Limited Whitland Wales UK, TCT Research tDCS Stimulator, TST Kowloon Hong Kong, ActivaDose Iontophoresis Delivery Unit produced by ActivaTek Inc., Chattanooga Ionto Iontophoresis System Hixson TN USA, and NewRonika, Italy HDCstim MI2011C002156.

Seventeen studies employed computational simulations, with SimNIBS [53] being the most frequently used simulation program, followed by ROAST, tDCSExploreTM (Soterix Medical Inc., New York, NY, USA), and StimViewer. Among those reporting the type of control/sham stimulation, 85 groups used a ramp-like stimulation method. Specifically, 70 groups incorporated a fade-in/fade-out phase lasting 8-40 seconds, while 14 groups used a fade-in phase followed by immediate cessation of stimulation. One study used 0.1 mA as sham stimulation.

The most commonly employed motor learning paradigm was variations of the Finger Tapping Task (FTT), which was used in 27 studies. Additionally, 11 studies utilized the Sequential Visual Isometric Pinch Task (SVIPT), 9 studies employed laparoscopic/surgical tasks and/or the Purdue Pegboard Task (PPT), 13 studies utilized various tracing tasks such as manipulandum, Vienna, or arc pointing tasks, two studies used dart throwing, one study used cup stacking, and one study administered a balance tracking task.

Regarding outcome measures, 27 studies assessed skill measures that incorporated both temporal and spatial parameters. Eleven studies independently investigated speed (e.g., reaction time or completion time), and eight studies focused solely on accuracy or precision (e.g., error rate or distance from the target). Sixteen studies examined both accuracy and speed separately, without integrating them into a single skill measure. Two studies independently explored all three dimensions: skill, accuracy, and speed.

In the application of tACS, 17 groups received stimulation of M1, two groups received stimulation of CB, and one group received stimulation of M1 and CB. tACS was administered in various frequency bands: delta (1 Hz) in one intervention group, alpha band (10 Hz) in four groups, beta-band (20-35 Hz) in five groups, gamma-band (40-100 Hz) in eight groups, and frequencies higher than the gamma band in the two groups. Stimulation intensity varied, with four studies using 2 mA peak-to-peak and five studies using 1 mA peak-to-peak. All studies utilized carbon rubber electrodes. Specifically, five studies used sponges with saline solution as the contact medium, and four studies employed a conductive gel (e.g., Ten20 conductive paste). Electrode placement was determined based on the international 10-20 EEG system in five studies, the TMS hotspot in two studies, and both methods in two studies. Regarding the devices used, seven studies employed the NeuroConn DC stimulator (NeuroConn, Ilmenau, Germany), and two studies utilized devices from Soterix Medical Inc., New York, NY, USA. Three studies incorporated computational simulations: two used SimNIBS and one used tDCSExploreTM (Soterix Medical Inc., New York, NY, USA).

Among those reporting the type of control/sham stimulation, a 30-second stimulation period was the most commonly used.

The most frequently employed motor learning paradigm was variations in the FTT, which was used in five studies. Additionally, one study employed the SVIPT, one study used a bimanual tracing task, and one study utilized a tracing task (manipulandum). In terms of outcome measures, three studies investigated speed (e.g., reaction time or completion time) independently, two studies focused solely on accuracy or precision, and four studies examined both accuracy and speed separately.

### 3.3 | Quantitative analyses

The pooled effect sizes in the different subdivided domains were based on 203 observed measures of Hedges’ g based on 62 studies applying tDCS (with two studies excluded due to insufficient data for effect size extraction) and 32 extracted effect sizes from 9 studies investigating the effects of tACS application.

### 3.4 | Online learning

The following table presents a summary of the results of the meta-analyses conducted for the three target areas. Table 1 shows an overall estimate of 0.41, with a between-study variance of 0.11 for online learning in the groups receiving M1 tDCS. tDCS of CB demonstrated a SMD of 0.39, with a between-study variance of 0.32 for online learning. Finally, for higher cortical areas, a SMD of 0.38 was found with a between-study variance of 0.15. The statistical probability of having a SMD below 0.3, considered as less or no meaningful, was 3% for M1, 30.7% for CB, and 26.6% for HCA. In our analysis, the Pareto K diagnostic values for the M1 and CB observations were below 0.7, indicating that the LOO-CV approximation was reliable [48]. The specific results of our LOO-CV analysis showed an elpd of M1 [-82.2], CB [-22.4], and HCA [-19.1], with an effective number of parameters (p_loo) of M1 [16.9], CB [10.6], and HCA [5.5]. These diagnostics collectively indicate a robust model with good predictive accuracy and a minimal risk of overfitting. Because the LOO Pareto K diagnostics from the baseline model of HCA online learning identified the potential influence of extreme values in one of the cases, we performed leave-one-out cross-validation on the baseline model, which showed that model generalizability did not increase by adding ‘target are’ as a moderator. Adjusting priors also did not improve the predictive performance of the model. Using stimulation intensity as a moderator, modeled with ≤1 mA vs. >1 mA, did not influence the posterior estimate considerably (stimulation intensity ≤1: -0.03±0.09, [-0,2; 0.15]).

**Table 1:** Summary of Bayesian Meta-analyses of the effects of tDCS on online motor learning across the Primary Motor Cortex (M1), the Cerebellum (CB), and Higher Cortical Areas (HCA). Standardized Mean Difference (SMD), between-study heterogeneity (Tau), convergence diagnostics (Rhat), 95% credible intervals, effective sample sizes for both bulk and tail distributions representing the number of independent draws available from the central and extreme regions of the posterior distribution, respectively, Leave-One-Out Cross-Validation (LOO) metrics, Bayesian LOO estimate of the expected log pointwise predictive density (elpd), Pareto K diagnostic values, and Empirical Cumulative Distribution Function (ECDF) analysis.

| Online learning | SMD bias-corrected | Tau ($\tau$) | Rhat | 95% credible interval | Effective bulk and tail samples | LOO elpd/p_loo/ pareto-k | ECDF < 0.3 |
| --- | --- | --- | --- | --- | --- | --- | --- |
| **MI** | 0.41 | 0.11 | 1 | [0.3; 0.52] | 70391 \|\| 53953 | -82.2 \|\| 16.9 \|\| K<0.7 | 3% |
| **CB** | 0.39 | 0.32 | 1 | [0.01; 0.66] | 45065 \|\| 44870 | -22.4 \|\| 10.6 \|\| K<0.7 | 30.7% |
| **HCA** | 0.38 | 0.15 | 1 | [0.13; 0.63] | 88934 \|\| 55641 | -19.1 \|\| 5.5 \|\| K(<0.7)=95%,<br>K(0.7-1)=5% (bad) | 26.6% |

#### 3.4.1 | Online motor learning – Motor Cortex

The pooled effect size based on 85 measures of Hedges’ g from 57 intervention groups in 48 studies (Fig. 2) revealed an overall estimate of 0.41 with a 95% CI [0.3, 0.52], with a between-study variance of 0.11 [0.11, 0.43], and within-study variance of 0.12 [0.00, 0.29]. According to Cohen’s (1988) [37] criteria for effect size estimates, this pooled result indicated a medium positive effect. A SMD of this size implies that there is approximately a 61% (common language effect size) probability that randomly chosen individuals will perform better with tDCS compared to sham in terms of online motor learning [54].

**Figure 2:**
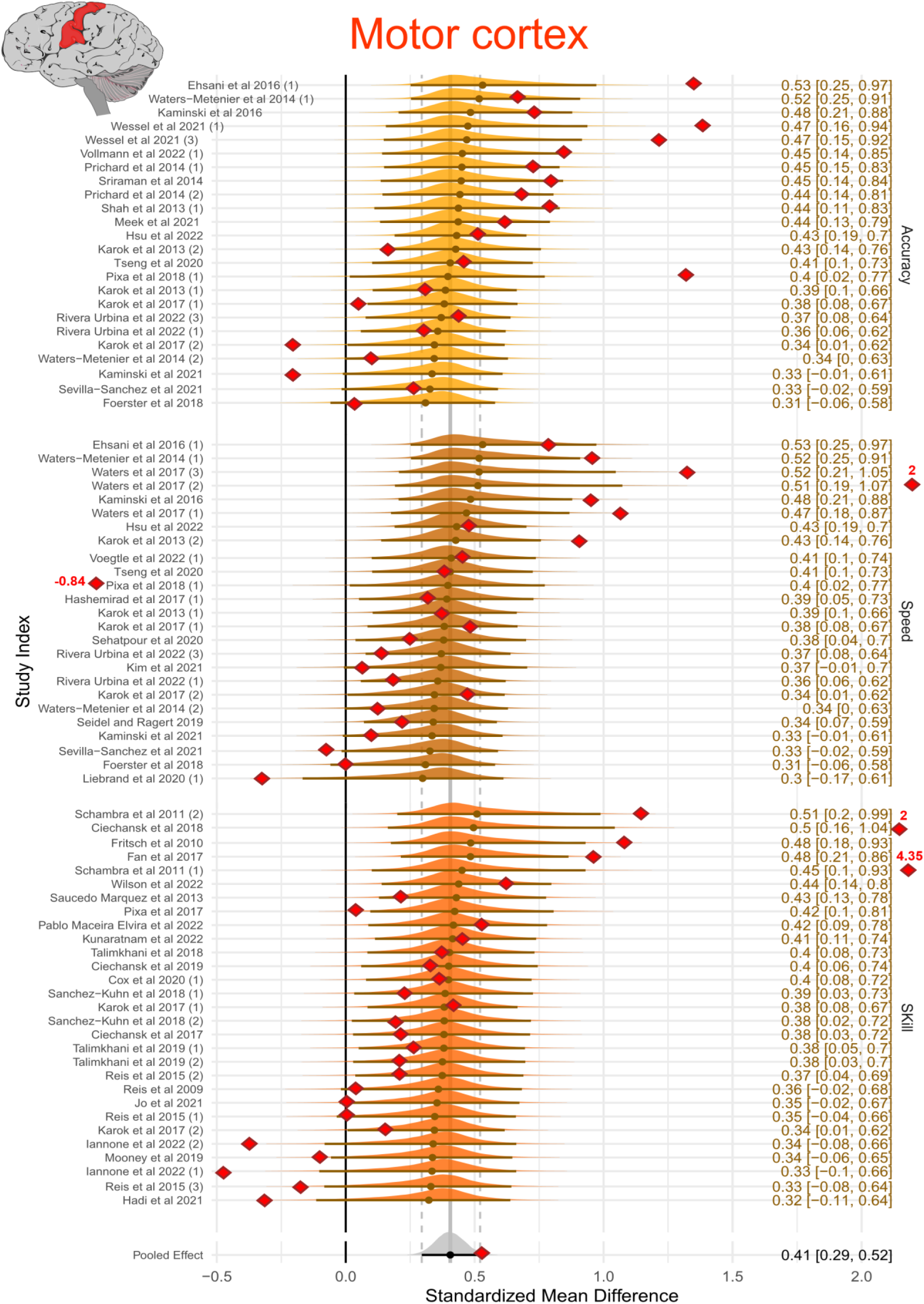
Estimated effect sizes for experimental studies of online motor learning that applied tDCS to the primary motor cortex - M1. Studies appearing multiple times with numerical differentiation (e.g., 1, 2, 3) represent distinct experimental groups or intervention arms within the same study. The colored distributions represent the posterior probability distributions for each effect size estimated. Values on the right display the posterior mean standardized effect size (Hedges’ g) along with the corresponding 95% credible intervals. Red squares depict the observed (raw) effect sizes, providing a visual comparison to the model-based estimates. The pooled standardized mean difference (SMD) is shown both as a grey posterior distribution (Bayesian estimate) and a red square (Frequentist estimate), serving as a reference point across studies.

#### 3.4.2 | Online motor learning – Cerebellum

The pooled effect size of CB stimulation on online learning was based on 22 measures of Hedges’ g from 16 intervention groups in 16 studies (Fig. 3) and revealed an overall estimate of 0.39 with 95% CI [0.08, 0.73], with a between-study variance of 0.32 [0.02, 0.8] and within-study variance of 0.23 [0.01, 0.66]. According to Cohen’s (1988) [37] criteria for effect size estimates, this pooled result indicates a small-to-medium positive effect. An SMD of this size implies that there is an approximately 60% (common language effect size) probability that randomly chosen individuals will exhibit more online learning with real tDCS stimulation compared to sham.

**Figure 3:**
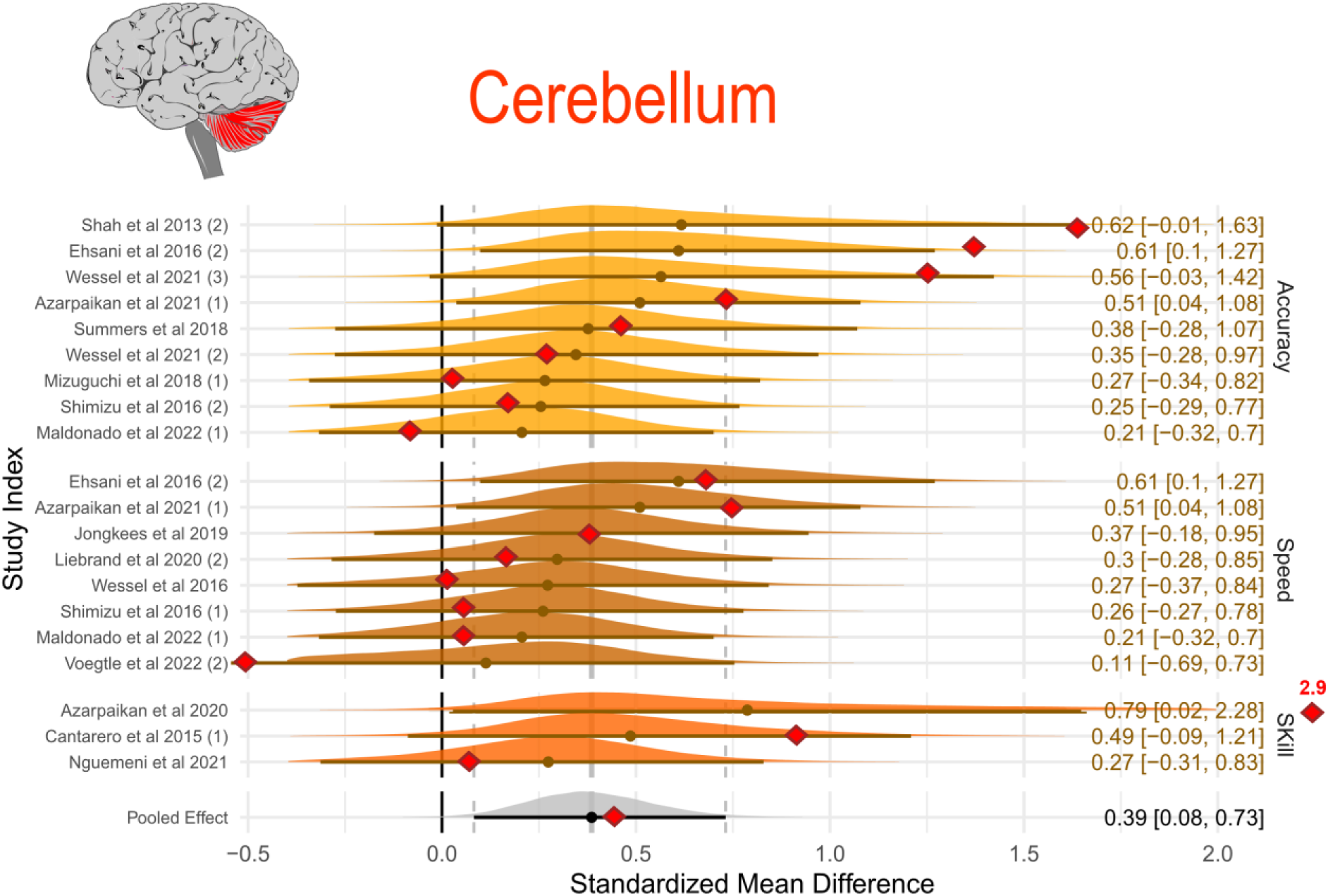
Estimated effect sizes for experimental studies of online motor learning that applied tDCS to CB. Studies appearing multiple times with numerical differentiation (e.g., 1, 2, 3) represent distinct experimental groups or intervention arms within the same study. The colored distributions represent the posterior probability distributions for each effect size estimated. Values on the right display the posterior mean standardized effect size (Hedges’ g) along with the corresponding 95% credible intervals. Red squares depict the observed (raw) effect sizes, providing a visual comparison to the model-based estimates. The pooled standardized mean difference (SMD) is shown both as a grey posterior distribution (Bayesian estimate) and a red square (Frequentist estimate), serving as a reference point across studies.

#### 3.4.3 | Online motor learning – Higher cortical areas

The pooled effect size of tDCS targeting HCA on online learning based on 20 measures of Hedges’ g from 17 intervention groups in 17 studies (Fig. 4) revealed an overall estimate of 0.38 with 95% CI [0.13, 0.63], with a between-study variance of 0.15 [0.01, 0.43] and within-study variance of 0.14 [0.01, 0.41]). According to Cohen’s (1988) [37] criteria for effect size estimates, this pooled result indicates a small to medium positive effect. A standardized mean difference of this size implies an approximately 60% probability that randomly chosen individuals will perform better with tDCS compared to sham.

**Figure 4:**
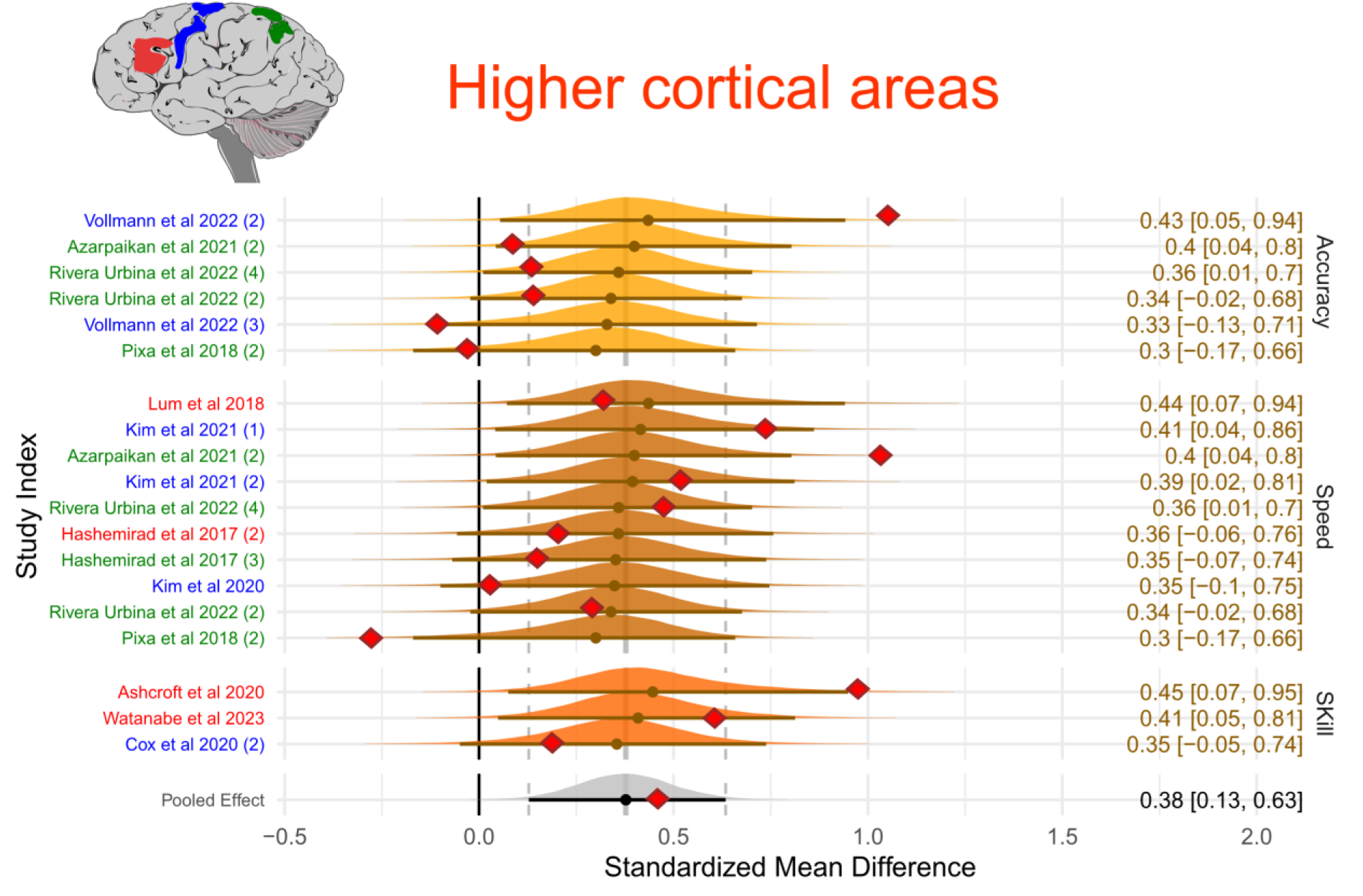
Estimated effect sizes for online motor learning that apply tDCS to HCA across experimental studies. Studies appearing multiple times with numerical differentiation (e.g., 1, 2, 3) represent distinct experimental groups or intervention arms within the same study. Colors assigned to each study correspond to the specific cortical target area stimulated. The colored distributions represent the posterior probability distributions for each effect size estimated. Values on the right display the posterior mean standardized effect size (Hedges’ g) along with the corresponding 95% credible intervals. Red squares depict the observed (raw) effect sizes, providing a visual comparison to the model-based estimates. The pooled standardized mean difference (SMD) is shown both as a grey posterior distribution (Bayesian estimate) and a red square (Frequentist estimate), serving as a reference point across studies.

### 3.5 | Offline learning

The following table presents a summary of the results of the meta-analyses conducted for offline learning. Table 2 shows an overall estimate of 0.23, with a between-study variance of 0.1 M1 tDCS. tDCS targeting CB demonstrated an SMD of 0.16, with a between-study variance of 0.38 for offline learning and -0.1 SMD, with a between-study variance of 0.2 for application at HCA. The likelihood of observing an SMD below 0.3, which is deemed a small effect size, was 87.7% for M1, 67.6% for CB, and 93.3% for HCA. The Pareto K diagnostic values for M1 and HCA observations were below 0.7, indicating that the LOO-CV approximation was reliable. The specific results of our LOO-CV analysis showed an elpd of M1 [-30.7], CB [-15.8], and HCA [-7.8], with an effective number of parameters (p_loo) of M1 [6.5], CB [6.6], and HCA [2.3]. Since the LOO Pareto K diagnostics from the baseline model of CB offline learning identified the potential influence of extreme values in one of the cases, we performed leave-one-out cross-validation on the baseline model, which showed that model generalizability did not increase after adjusting. After further adjusting for the potential effect of distant delayed retention testing, we added ‘delayed retentioń as a moderator, which did not improve the predictive performance of the model. Using stimulation intensity as a moderator, modeled with ≤1 mA vs >1 mA, did not influence the posterior estimate considerably (stimulation intensity ≤1: -0.01±0.12, [-0,24; 0.22]).

**Table 2:** Summary of Bayesian Meta-analyses of the effects of tDCS on offline motor learning targeting the Primary Motor Cortex (M1), the Cerebellum (CB), and Higher Cortical Areas (HCA). Standardized Mean Difference (SMD), between-study heterogeneity (Tau), convergence diagnostics (Rhat), 95% credible intervals, effective sample sizes for both bulk and tail distributions, Leave-One-Out Cross-Validation (LOO) metrics, Bayesian LOO estimate of the expected log pointwise predictive density (elpd), Pareto K diagnostic values, and Empirical Cumulative Distribution Function (ECDF) analysis.

| Offline learning | SMD bias-corrected | Tau ($\tau$) | Rhat | 95% credible interval | Effective bulk and tail samples | LOO elpd/$p_{loo}$/pareto-k | ECDF < 0.3 |
| --- | --- | --- | --- | --- | --- | --- | --- |
| **M1** | 0.23 | 0.1 | 1 | [0; 0.27] | 110800 \|\| 55407 | -30.7 \|\| 6.5 \|\| K<0.7 | 87.7% |
| **CB** | 0.16 | 0.43 | 1 | [-0.52; 0.87] | 58258 \|\| 49317 | -16 \|\| 6.7 \|\| K(<0.7)=50%, K(0.7-1)=50% (bad) | 67.6% |
| **HCA** | -0.01 | 0.2 | 1 | [-0.43; 0.42] | 48197 \|\| 44186 | -7.8 \|\| 2.3 \|\| K<0.7 | 93.3% |

#### 3.5.1 | Offline motor learning – Motor cortex

The pooled effect size based on 54 measures of Hedges’ g from 47 intervention groups in 38 studies (Fig. 5) reveals an overall estimate of 0.23 with 95% CI [0.1, 0.35], with a between-study variance of 0.1 [0.0, 0.27] and within-study variance of 0.12 [0.00, 0.26]. According to Cohen’s (1988) [37] criteria for effect size estimates, this pooled result indicates a small positive effect. A SMD of this size implies an approximately 56 % probability that randomly chosen individuals will perform better with tDCS.

**Figure 5:**
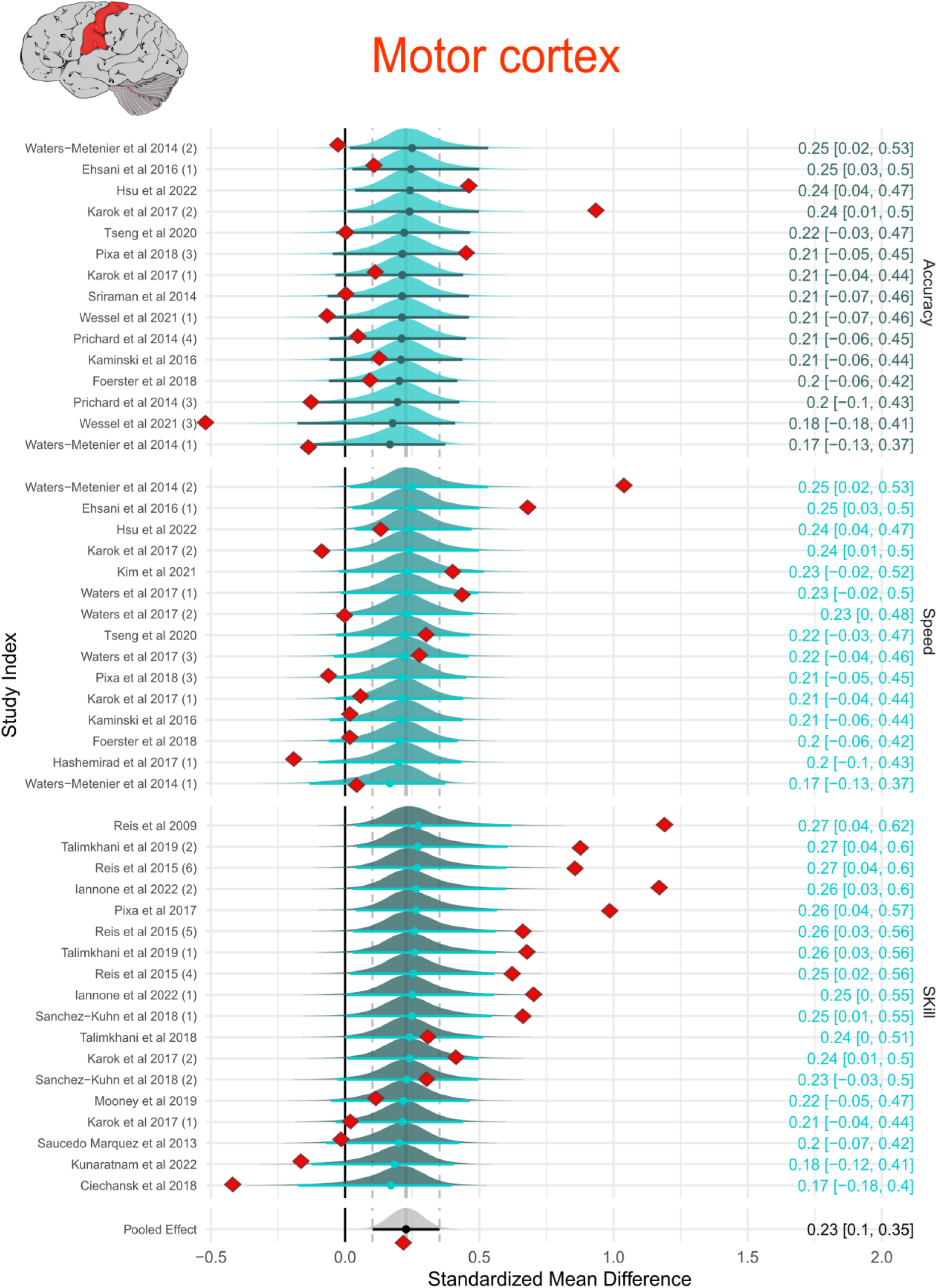
Estimated effect sizes for offline motor learning that apply tDCS to M1 across experimental studies. Studies appearing multiple times with numerical differentiation (e.g., 1, 2, 3) represent distinct experimental groups or intervention arms within the same study. The colored distributions represent the posterior probability distributions for each effect size estimated. Values on the right display the posterior mean standardized effect size (Hedges’ g) along with the corresponding 95% credible intervals. Red squares depict the observed (raw) effect sizes, providing a visual comparison to the model-based estimates. The pooled standardized mean difference (SMD) is shown both as a grey posterior distribution (Bayesian estimate) and a red square (Frequentist estimate), serving as a reference point across studies.

#### 3.5.2 | Offline motor learning – Cerebellum

The pooled effect size of CB offline learning based on 10 measures of Hedges’ g from 8 intervention groups in 7 studies (Fig. 6) revealed an overall estimate of 0.16 with a 95% CI [-0.53, 0.87], with a between-study variance of 0.43 [0.01, 1.15] and within-study variance of 0.38 [0.01, 1.15]. According to Cohen’s (1988) [37] criteria for effect size estimates, this pooled result indicates a small positive effect. A SMD of this size implies that there is an approximately 54% probability that randomly chosen individuals will perform better with tDCS.

**Figure 6:**
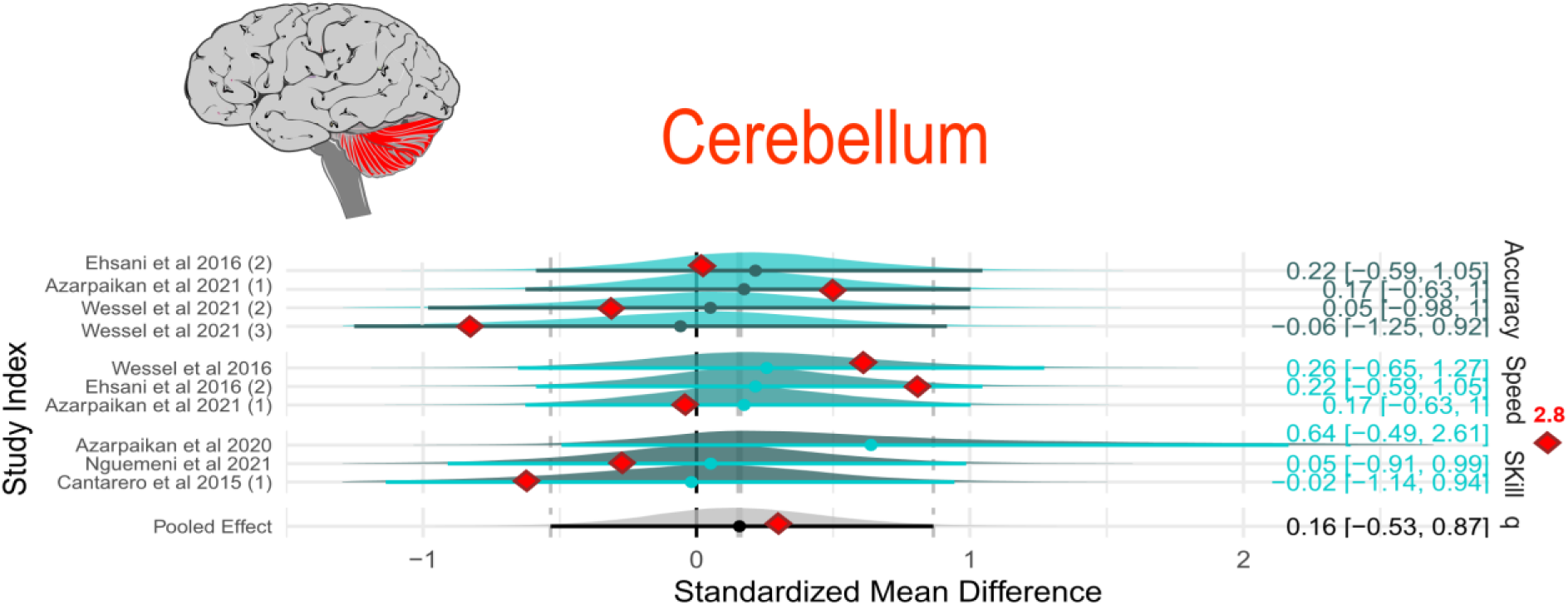
Estimated effect sizes for offline motor learning that apply tDCS to M1 across experimental studies. Studies appearing multiple times with numerical differentiation (e.g., 1, 2, 3) represent distinct experimental groups or intervention arms within the same study. The colored distributions represent the posterior probability distributions for each effect size estimated. Values on the right display the posterior mean standardized effect size (Hedges’ g) along with the corresponding 95% credible intervals. Red squares depict the observed (raw) effect sizes, providing a visual comparison to the model-based estimates. The pooled standardized mean difference (SMD) is shown both as a grey posterior distribution (Bayesian estimate) and a red square (Frequentist estimate), serving as a reference point across studies.

#### 3.5.3 | Offline motor learning – Higher cortical areas

The pooled effect size of HCA offline learning based on 9 measures of Hedges’ g from 7 intervention groups in 6 studies (Fig. 7) revealed an overall negative estimate of -0.01 with 95% CI [-0.43, 0.42], with a between-study variance of 0.19 [0.01, 0.64] and within-study variance of 0.21 [0.01, 0.59]. According to Cohen’s (1988) [37] criteria for effect size estimates, this pooled result indicates a negative effect.

**Figure 7:**
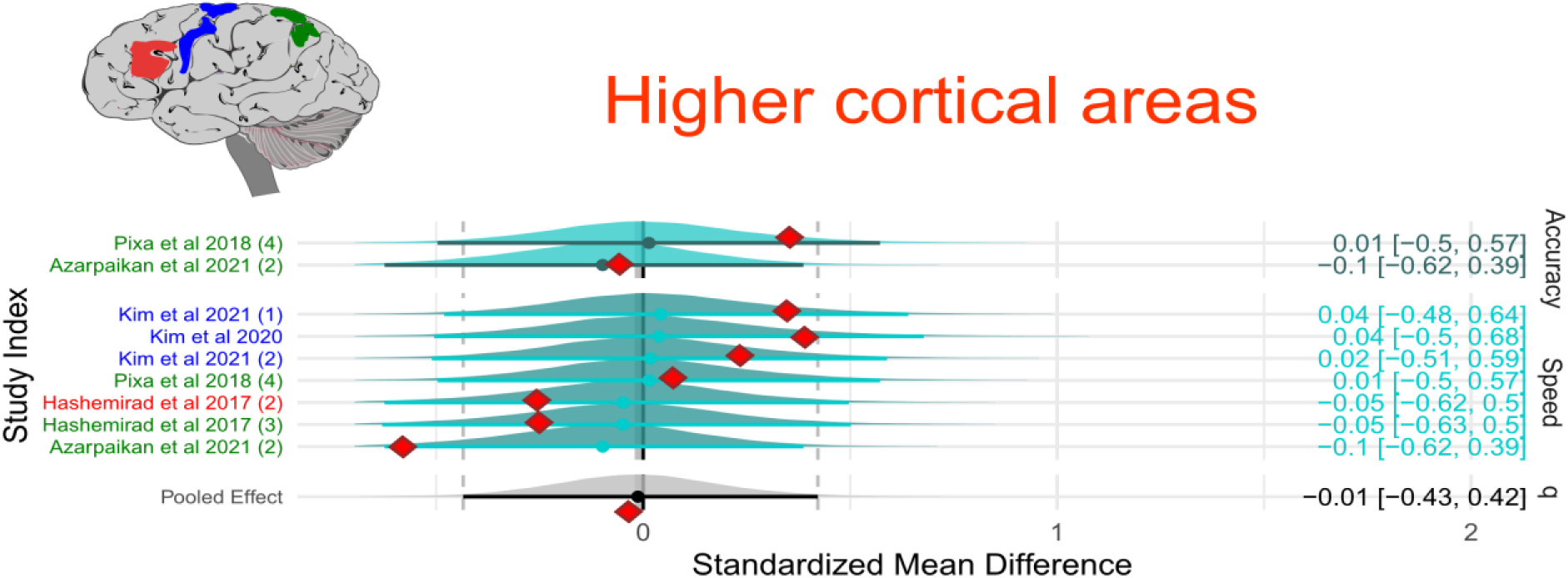
Estimated effect sizes for offline motor learning that apply tDCS to HCA across experimental studies. Studies appearing multiple times with numerical differentiation (e.g., 1, 2, 3) represent distinct experimental groups or intervention arms within the same study. Colors assigned to each study correspond to the specific cortical target area stimulated. The colored distributions represent the posterior probability distributions for each effect size estimated. Values on the right display the posterior mean standardized effect size (Hedges’ g) along with the corresponding 95% credible intervals. Red squares depict the observed (raw) effect sizes, providing a visual comparison to the model-based estimates. The pooled standardized mean difference (SMD) is shown both as a grey posterior distribution (Bayesian estimate) and a red square (Frequentist estimate), serving as a reference point across studies.

### 3.6 | Single sessions vs multiple sessions

27 studies used a motor practice paradigm comprising more than one session, and 37 studies investigated the effects of tDCS after a single practice session. The largest number of sessions was 20, but the most frequent practice sessions fell within the 3-5 session range.

We modeled the number of sessions with tDCS applied concurrently with motor practice (e.g., binarization for single vs. multiple sessions) and evaluated a separate model that included the number of sessions as a moderator. Multiple sessions had an overall positive influence on the estimate (Estimate: 0.33; 95% CI: [0.3, 0.69]), whereas a single session had a negative impact on the estimate (Estimate: - 0.11; 95% CI: [-0.34, 0.11]) This coefficient indicates a change in the effect size associated with a single session compared to multiple sessions (Fig. 8). The marginal effect difference between single and multiple sessions was associated with 0.4-point greater predicted estimates, and this difference pertained to training efficacy and the impact of repeated stimulation sessions on skill acquisition. Bayesian meta-analysis produces a posterior distribution of the difference in the probabilities of the number of sessions categorized in this analysis as single vs. multiple sessions, as shown in Fig. 8C [55]. The top panel of Fig. 8C shows the marginal difference in the probabilities ωT −ωC (bottom panel as the joint distribution), which represents the average difference in effect sizes between single and multiple sessions, expressed in terms of pooled standard mean deviations across the studies.

**Figure 8:**
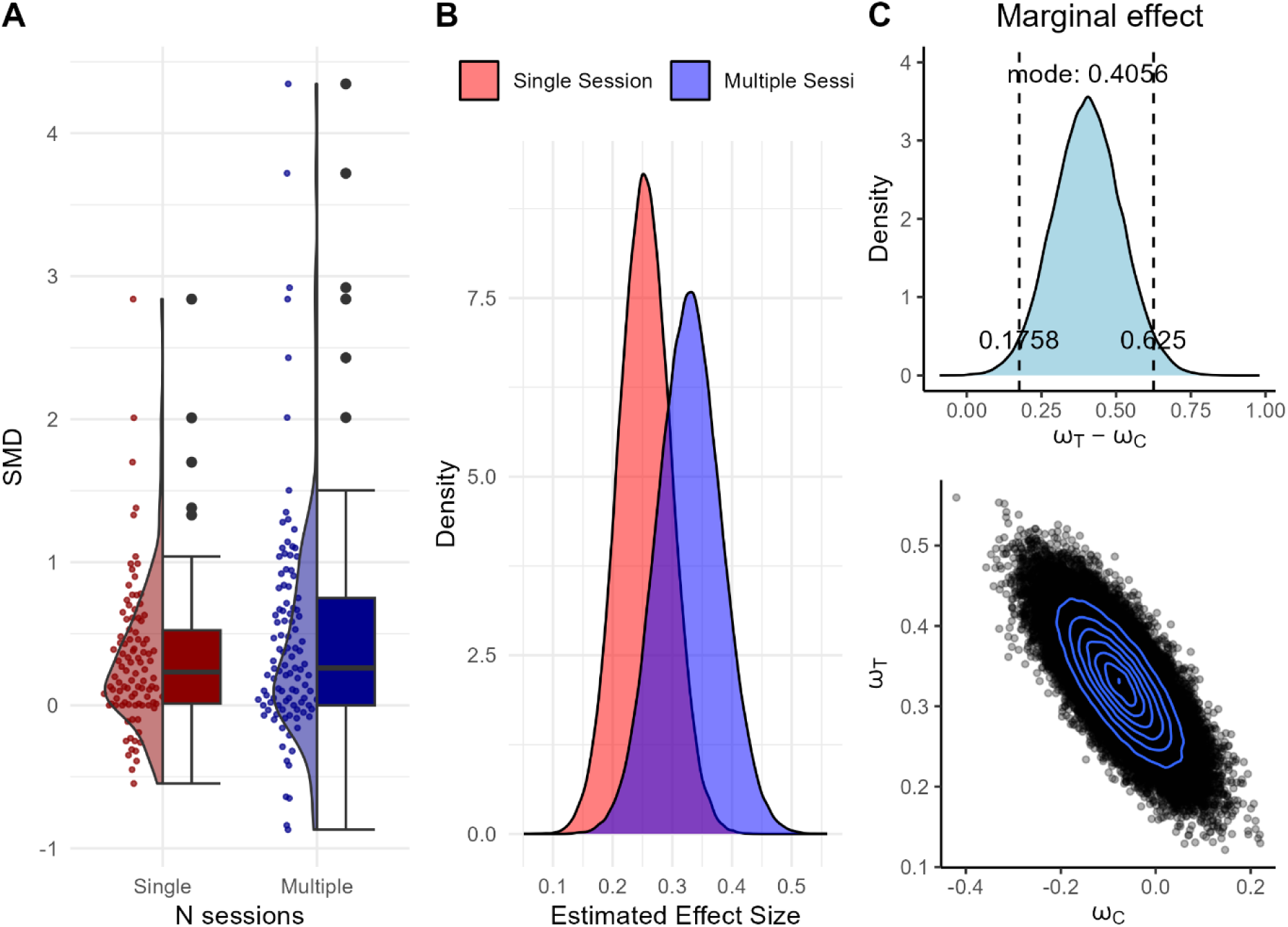
Single- vs multisession tDCS: A) SMD for each study divided by the number of tDCS sessions. B) Estimated effect sizes for online motor learning that applied tDCS to M1 across experimental studies. The shaded areas indicate the posterior probability density of each estimate. Model-based influence of the number of tDCS and motor practice sessions on the estimated effect size. C, top: the marginal difference of the probabilities, ωT (multi-session) − ωC (single-session). The blue shaded area illustrates the estimated density, with vertical dashed lines marking the bounds of the 95% Highest Density Interval (HDI). and bottom: Joint posterior distribution of the group-level probability estimates for single-session and multisession tDCS. Each dot represents a posterior draw from the model, and the overlaid blue contour lines indicate regions of highest density, reflecting the correlation and uncertainty between the two estimates.

### 3.7 | Transcranial Alternating Current Stimulation

Table 3 and Figure 9 reveal an overall estimate of 0.11, with a between-study variance of 0.13 for tACS across all the frequency domains. The statistical probability of having an SMD below 0.3 considered as less or no meaningful, was 97.8%. The specific results of our LOO-CV analysis showed an elpd of [-20.8], with an effective number of parameters (p_loo) of [12.3]. Because the LOO Pareto K diagnostics from the tACS Bayesian model identified the potential influence of extreme values in three cases, we performed leave-one-out cross-validation on the baseline model, which showed that model generalizability did not increase by adding ‘tACS frequencies applied’ as a moderator, meaning that the individual frequency band could not explain the model estimate. Adjusting priors did also not improve model predictive performance.

**Table 3:** Summary of Bayesian Meta-analyses of the effects of tACS on online motor learning. Standardized Mean Difference (SMD), between-study heterogeneity (Tau), convergence diagnostics (Rhat), 95% credible intervals, effective sample sizes for both bulk and tail distributions, Leave-One-Out Cross-Validation (LOO) metrics, Bayesian LOO estimate of the expected log pointwise predictive density (elpd), Pareto K diagnostic values, and an Empirical Cumulative Distribution Function (ECDF) analysis.

| tACS | SMD<br>bias-corrected | Tau<br>($\tau$) | Rhat | 95%<br>credible<br>interval | Effective<br>bulk and tail<br>samples | LOO<br>elpd/$p_{loo}$/pareto-k | ECDF<br>< 0.3 |
| --- | --- | --- | --- | --- | --- | --- | --- |
| Overall | 0.11 | 0.13 | 1 | [-0.07; 0.3] | 9011 \|\| 2625 | -20.8 \|\| 12.3 \|\| K(<0.7)=90.6%,<br>K(0.7-1)=9.4% (bad) | 97.8% |

**Figure 9:**
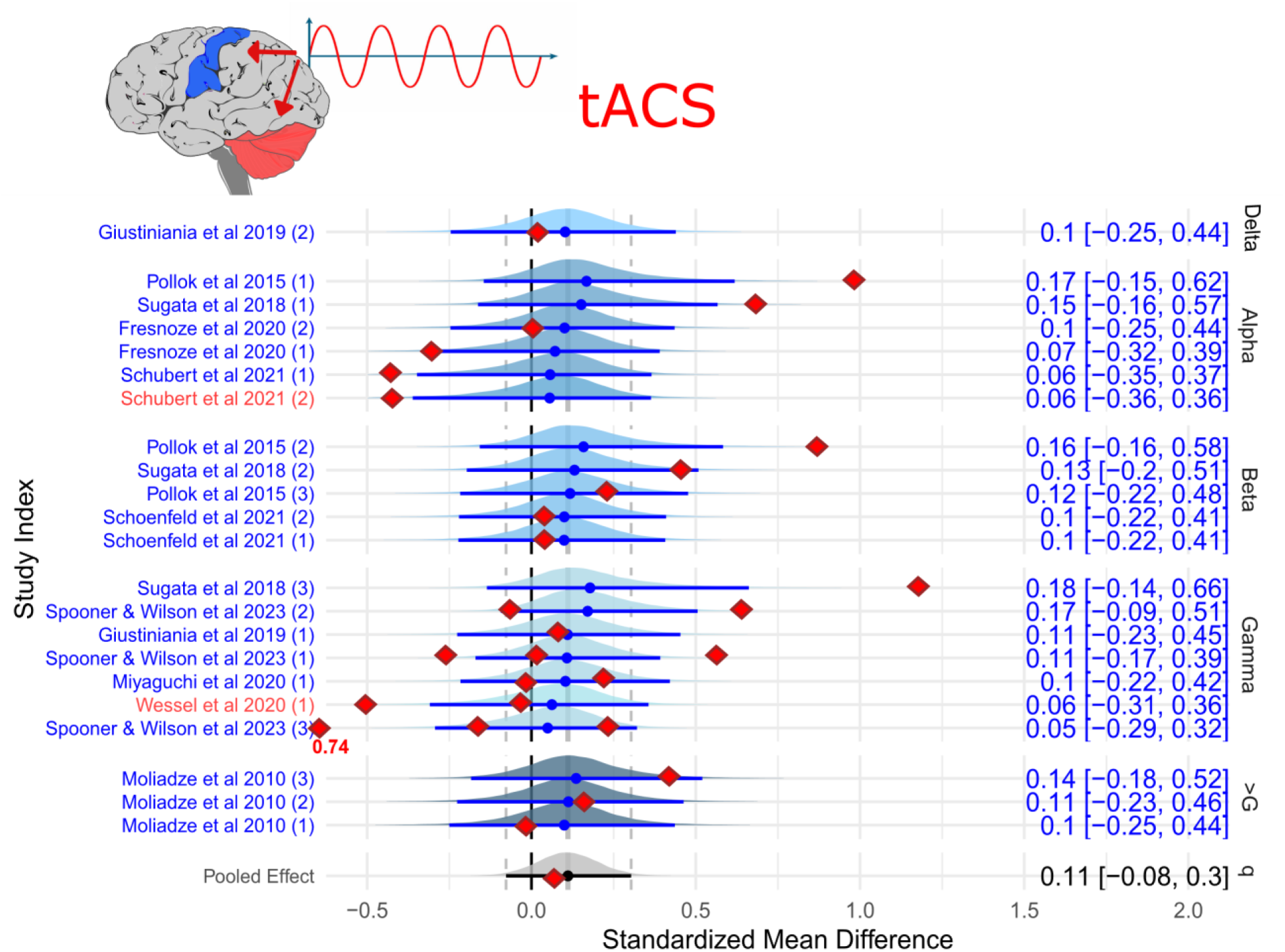
Estimated effect sizes for application of tACS across experimental studies. The forest plot is divided into sections from low-frequency bands to higher-frequency bands. The shaded areas indicate the posterior probability density of each estimate. The blue shaded areas indicate tACS applied to M1. The red shaded areas indicate tACS applied to the CB. The numbers to the right provide the estimated mean effect size (Hedges’ g) and the upper and lower 95% credible intervals. The red squares represent the observed effect size in contrast to the model-based estimated effect size.

### 3.8 | Publication bias of Transcranial Direct Current Stimulation

#### 3.8.1 | Sensitivity Analysis

Figure 10A illustrates the sensitivity analysis results for the meta-analytic effect size estimates in the tDCS analyses. The X-axis represents different levels of publication bias, quantified as the fold increase in the likelihood of significant studies being published compared with non-significant studies. The Y-axis shows the effect size estimate, with the dark-shaded area indicating the 95% credible interval. Our analysis reveals that as the assumed publication bias increases (up to a factor of 200), the estimated effect size decreases, but remains above zero. This finding suggests that the observed effect size is not solely a product of publication bias. The credible intervals widen with increasing publication bias, reflecting greater uncertainty. However, the lower bound of the credible interval consistently remained above the null effect size, indicating robustness of our findings. The red dashed line represents the worst-case scenario estimate based on the “pubbias_meta” analysis, which remained constant and above zero across all levels of publication bias. This finding further supports the robustness of the observed effect size. At a selection ratio of 1 (indicating no publication bias), the effect size estimate was approximately 0.49. When assuming a moderate publication bias (selection ratio of 3), significant studies are three times more likely to be published than non-significant studies. Even in this scenario, the worst-case effect size estimate remained above zero. The analysis indicates that regardless of the severity of publication bias, our meta-analysis provides strong evidence for an average effect in the observed direction, albeit possibly of a smaller magnitude. The fact that the worst-case scenario estimate is above zero suggests that the positive effect observed is robust and unlikely to be null even when accounting for extreme publication bias.

**Figure 10:**
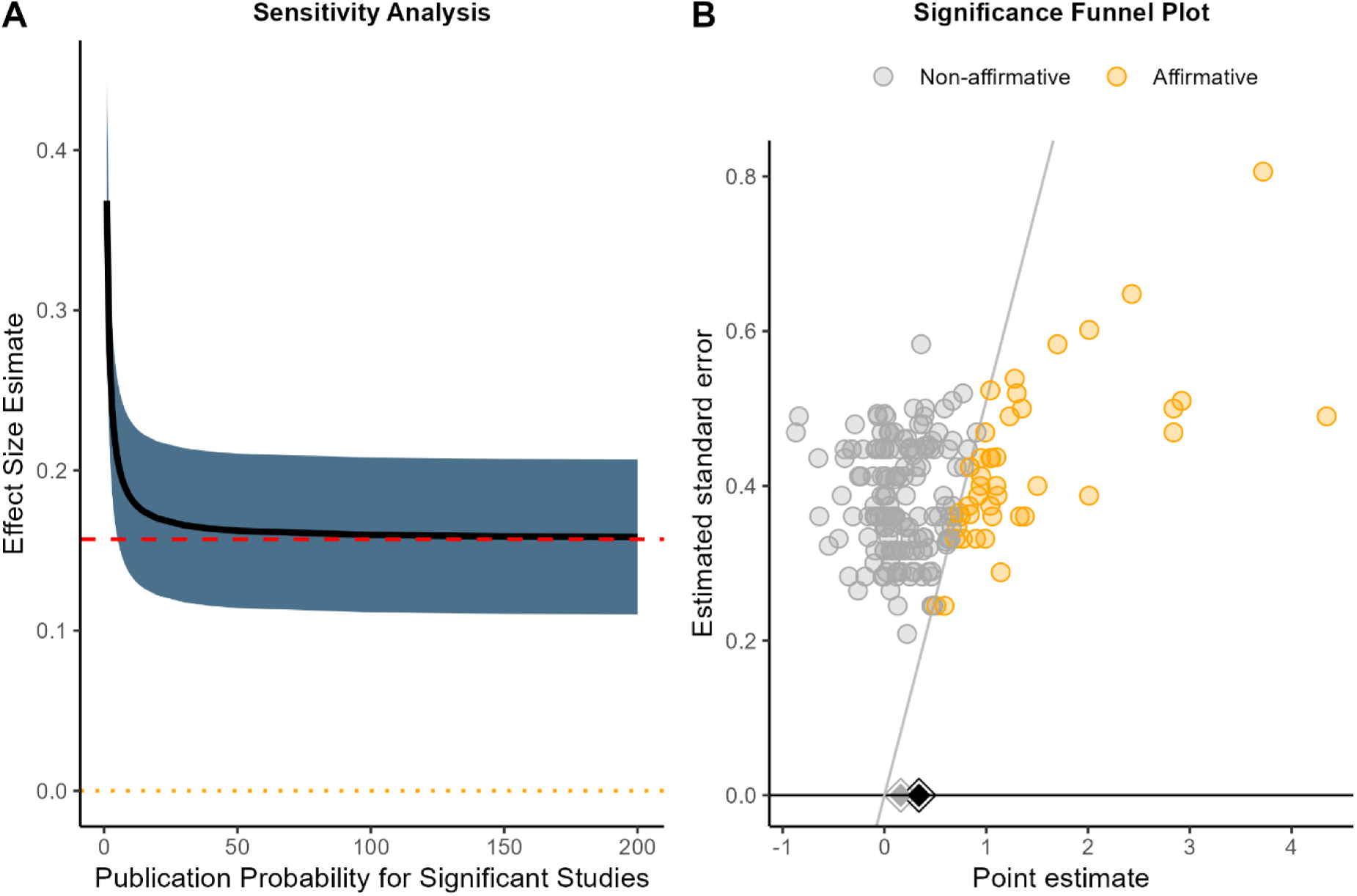
A) Plot of sensitivity analysis showing the effect size estimate as a function of severity of publication Bias. The plot indicates what happens to the effect size if the publication probability is x times higher for significant studies compared to non-significant studies. An effect size estimate of 0.0 is indicated by the orange dotted line and the worst-case point estimate (see below) is indicated by the dashed red line. B) Significance funnel plot of the studies. Studies on the diagonal line have exactly p = 0.05. Grey diamonds denote worst-case estimates of effect size based only on nonsignificant studies. The black diamond denotes an estimate of the effect size for all studies. The significance funnel plot indicates a weak correlation between the point estimates and their standard errors.

By using the inverse of the sum of study variance and a heterogeneity estimate, we can compute the uncorrected worst-case estimate for effect size based solely on non-significant studies 0.157 with 95% CI [0.11, 0.21], as shown in the significance funnel plot in Figure 10B. Although the significance funnel plot indicated a weak correlation between the point estimates and their squared standard errors, the estimates for non-significant studies primarily revolved around the null. This distributional pattern further implies that the meta-analytic estimate is robust to publication biases. Proximity of the diamond-shaped estimates in the visualization suggests consistency across methods, indicating that the meta-analysis results are relatively robust to publication bias, as supported by the quantitative sensitivity analyses conducted. In addition to the results of the sensitivity analysis, these findings indicate that our estimates are less likely to be inflated. Thus, overall, we can conclude that regardless of the severity of publication bias, this meta-analysis provides strong evidence for an average effect in the observed direction, albeit possibly of small size.

## 4 | Discussion

Our systematic review and meta-analysis of 64 tDCS and 9 tACS published studies investigating the effects of weak electrical brain stimulation applied concurrently with motor practice yielded several main findings. First, tDCS demonstrates a greater positive effect on motor skill learning than sham stimulation, with a pooled effect size in the small-to-medium range according to standard classifications. Second, the effects of stimulation on motor skill learning appear to favor within-session gains irrespective of the specific brain region targeted. Third, regarding the specificity of brain region targeting, stimulation was overall most effective, considering both online and offline learning, when the anode electrode was positioned over the primary motor cortex, producing beneficial effects on motor learning, both within and across sessions. Fourth, multiple sessions of stimulation combined with motor practice resulted in greater cumulative effects than single-session interventions. Publication bias analysis revealed that the overall effect size was robust, and a meaningful effect remained even when extreme publication bias was accounted for. Bayesian prevalence suggests a high probability of true efficacy for tDCS, with variation dependent on the brain region targeted, when considered within-study frequentist assessment, i.e., distribution of statistically significant and non-significant effects. Despite methodological heterogeneity in stimulation parameters and motor learning paradigms across studies, we found that tDCS systematically introduces a probable positive effect on motor skill learning.

### 4.1 | Online vs offline skill learning

Building on previous studies, our systematic review and meta-analysis support a positive impact of anodal tDCS on motor learning, with greater learning gains observed compared to sham stimulation. As reviewed, prior research has provided evidence for the ability of tDCS to induce additive learning effects in the motor domain [14], [56], [57]. Hashemirad et al. (2016) [56] included 13 studies and adopted several meta-analyses that quantified the effect of tDCS applications targeting the M1 on speed, accuracy, and skill measures in motor sequence learning. There were positive effects of a single session of tDCS on speed performance at retention (offline learning), but the analyses also revealed that the effects on accuracy were insignificant. Multiple tDCS sessions significantly affected skills both post-intervention (online learning) and retention. The limited number of studies included in the meta-analyses may have introduced some ambiguity. Our findings specifically highlight a more substantial effect of tDCS on online motor learning relative to offline learning, with a small-to-moderate effect size in online learning and a smaller, although still notable, effect in offline learning. These online gains from tDCS may occur independently of session frequency (see Section 4.1.3) and may be attributed to early synaptic potentiation facilitated by tDCS, which enhances pre-existing synaptic networks in the initial stages of skill acquisition [12], [58], [59]. Early studies on anodal tDCS have documented increased cortical excitability, suggesting that LTP-like mechanisms in the cortex could underlie these immediate behavioral improvements [8], [58], [60]. However, no definitive link between the neurophysiological and behavioral effects of tDCS has been established, and changes in one domain do not consistently predict changes in the other, highlighting the need for further exploration of these mechanisms [6]. The difficulty in establishing a direct link between changes in behavior and the underlying neurophysiological mechanisms may partly stem from the nature of externally induced weak electric currents. Such stimulation often produces non-focal electric fields (see Section 4.6), complicates control over the direction of current flow owing to cortical gyrification, and may involve current intensities that are insufficient to robustly influence neuronal activity [25], [61], [62]. Although radially oriented currents have been shown to bias spike timing by modulating ionic flow across the membrane [63], [64], externally applied currents typically generate tangential flows that predominantly affect axon terminals rather than somatic compartments [28]. Consequently, the traditional somatic-doctrine principle may be overly simplistic, as it does not adequately account for the complex factors that constrain the behavior of the induced electric fields in the brain [65]. The offline learning data extracted in this meta-analysis reflect a range of temporal resolutions, capturing both cumulative between-session gains over several days and single-session consolidation phases from post-practice to retention tests, which can vary from hours to days. This heterogeneity in consolidation intervals presents a limitation, as it may obscure a consistent effect of tDCS on neural consolidation processes, even though distant delayed retention testing did not influence the model estimate. Despite these variations, our findings suggest that offline learning gains remain positively influenced by concurrent tDCS application, when anodal tDCS is applied at either M1 or CB, although to a lesser extent than online learning gains.

### 4.2 | Interaction parameters

Our analyses revealed that online learning effects were comparable across tDCS applications targeting different brain areas, with a slightly greater overall effect observed when stimulating the M1. Both within-session and between-session analyses favored M1 stimulation. These findings align with prior quantitative analyses, such as those reported by Hashemirad et al. (2016) [56] and Kumari et al. (2019) [57], who investigated the effects of tDCS on M1 and the cerebellum (CB), respectively. Although our findings regarding M1 stimulation are consistent with those of earlier studies, our results for CB tDCS deviate from those of Kumari et al. (2019) [57]. Specifically, Kumari et al. (2019) [57], who included 17 studies in their review, concluded that CB tDCS enhances short-term (<24 hours) and long-term (>24 hours) motor skill learning, but does not significantly impact online learning gains during and immediately after stimulation. This discrepancy likely stems from differences in the study selection criteria. In this meta-analysis, we included only studies that applied tDCS concurrently with motor practice and imposed stringent inclusion criteria for outcome measures, which may have captured a broader range of online learning effects. The vast methodological diversity in tDCS research—spanning variations in brain area targeting, task characteristics, outcome measures, and session numbers—presents a challenge for establishing consistent effects. No specific combination of stimulation parameters has demonstrated robust, universal efficacy across behavioral outcomes. A recent systematic review of 31 studies suggested that there is a tDCS effect on motor learning when M1 or CB is stimulated [14]. Guimarães and co-workers suggested that tDCS effects do not depend on motor task characteristics or assembly specifications alone but on their interaction. Their systematic review proposed a decision tree to guide future research on selecting stimulation parameters and designing experiments, emphasizing the complexity of these interactions. We employed separate models that incorporated outcome measures and task types as moderators to examine how these variables might influence model predictions. The results indicated no substantial change in the weighted contribution of different outcome measures, suggesting that the tDCS effects were consistent across various outcomes. Similarly, no meaningful differences emerged regarding task characteristics. Hashemirad et al. (2016) [56] also subdivided their meta-analysis based on behavioral outcomes across two motor learning paradigms – specifically movement speed, accuracy, and skill within the SEQTAP/SRTT and SVIPT paradigms – and found no difference in effect sizes between these outcome categories post-intervention. However, it is important to recognize that their sub-analyses included only 2 to 5 studies per category, resulting in substantial uncertainty in the pooled effect estimates. In contrast, our Bayesian analyses, supported by a larger set of included studies, revealed that neither the outcome measure nor the task type meaningfully influenced the model fit, suggesting a consistent effect of tDCS across different outcome categories. Furthermore, stimulation intensity, ranging from 0.3 mA to 4 mA, did not emerge as an explanatory factor for the observed effect sizes, indicating that variations in stimulation intensity within this range do not substantially impact the efficacy of tDCS. Given the vast parameter space associated with tDCS paradigms, we chose not to further investigate how individual stimulation specifications influence the model predictions. While many studies share broadly similar stimulation parameters, subtle variations can introduce substantial heterogeneity, complicating fair comparisons and limiting the ability to draw robust conclusions regarding specific parameter effects. Future research may benefit from systematic factorial designs to disentangle these interactions and establish optimized stimulation protocols.

### 4.3 | Number of sessions

Our analysis revealed that multiple-session tDCS protocols yielded larger effects on motor learning outcomes compared to single-session interventions. This finding offers valuable guidance for optimizing stimulation protocols, suggesting that repeated motor practice sessions with concurrently applied tDCS may be more effective in facilitating motor skill learning. To assess the influence of session number, we employed a Bayesian model that incorporated effect estimates across various time points, capturing both online and offline learning phases, and spanning multiple cortical target regions. While this aggregated approach enhances the precision of our probability estimates and allows for broader generalization, it comes at the cost of reduced specificity. Consequently, the results support a general positive effect of multiple-session stimulation across learning domains, but do not permit conclusions about any one specific protocol or cortical site. We suggest that the model’s generalizability provides a valuable foundation for future work to test more targeted hypotheses. Hashemirad and colleagues [56] demonstrated that multiple sessions had higher effects on motor learning, whereas their meta-analysis indicated that single-session tDCS does not seem to induce significant changes in behavioral outcomes during or immediately after stimulation [56]. Our analysis revealed more pronounced effects with multiple sessions compared to a single session; however, our model also indicated significant within-session learning gains following a single session. This suggests that the application of tDCS concurrently with motor practice likely yields favorable outcomes, surpassing those of sham conditions, after both single and multiple tDCS sessions combined with motor practice.

### 4.4 | tACS

In contrast to direct current stimulation, alternating current stimulation is a method that can entrain endogenous oscillatory brain activity [66]. Our meta-analysis revealed low effects of tACS on online motor learning, with a small SMD of 0.11. Furthermore, incorporating stimulation frequency as a moderator did not meaningfully alter model predictions, suggesting that the frequency applied during tACS did not differentially influence motor learning outcomes. This finding contrasts with prior meta-analyses in this domain, which reported larger effects of tACS on motor performance and function, but employed broader inclusion criteria or investigated different aspects of motor behavior. Hu et al. (2022) [67] conducted a meta-analysis that included studies investigating tACS effects on motor learning and reported a favorable SMD of 0.67 for experimental groups compared to controls. However, it is important to note that only four effect sizes were included in their analysis, which limits the robustness and generalizability of their findings. Additionally, the inclusion criteria of Hu et al. (2022) [67] encompassed studies with potentially varied methodological and experimental conditions, which could have introduced heterogeneity in the observed effects. Rostami et al. (2024) [68] further expanded the scope of tACS research by including studies that examined a wide range of motor functions, including upper and lower limb performance, balance, gait, and functional capacity, alongside motor learning. Their analysis included 16 studies and found that tACS applied over M1 improved motor function irrespective of stimulation frequency. Subgroup analyses revealed that the alpha, beta, and gamma frequencies applied individually positively affected motor learning. Their analysis yielded an overall SMD of 0.95 when tACS was applied concurrently with motor tasks. These findings highlight the potential of tACS as a powerful tool to enhance motor function. However, the inclusion of diverse motor domains makes it challenging to isolate the specific effects of tACS on motor learning. This distinction is crucial because the underlying mechanisms of motor learning differ significantly from those of motor performance or functional capacity. Our meta-analysis focused exclusively on motor learning and included only studies in which tACS was applied concurrently with practice. This stringent criterion enables a more targeted assessment of the effects of tACS on learning-specific processes, such as skill acquisition and retention, rather than on broader motor function. The absence of a strong effect in our findings highlights the potential complexity and variability of tACS efficacy in motor learning paradigms. Differences in study design, task characteristics, and stimulation parameters across the literature further underscore the importance of carefully defining inclusion criteria and outcome measures when interpreting the results. While prior meta-analyses suggest promising applications of tACS in enhancing motor function, our results emphasize the need for further investigation of its role in motor learning.

### 4.5 | Methodological considerations

Conducting a meta-analysis and systematic review involves inherent methodological challenges that must be carefully considered to ensure the validity and robustness of the conclusions. Like any meta-analytic approach, our analysis is only as reliable as the data incorporated. Our focus on the effects of tDCS on motor learning encompasses a wide array of experimental paradigms, brain-stimulation protocols, and outcome measures. This diversity introduces heterogeneity, which, while reflecting the broad applicability of tDCS, also complicates the interpretation of the pooled estimates.

One critical consideration is the variability in stimulation parameters, such as electrode placement, current intensity, and session duration, as well as differences in task types, such as motor sequence and visuomotor tracking tasks. Although our Bayesian framework accommodates heterogeneity by estimating posterior distributions and associated probabilities, it cannot fully overcome the limitations introduced by the inconsistencies across studies. While the Bayesian models account for extreme observed effect sizes and allow the inclusion of potential moderators to explore explanatory parameters, they remain constrained by the inherent limitations of a small number of included studies and the variability introduced by unique experimental designs. For example, the potential interaction between specific task characteristics and stimulation protocols may not be adequately captured, particularly when studies provide insufficient detail or use nonstandardized methodologies. Another important aspect is the selection of the moderator variables. While we incorporated variables such as the number of sessions, outcome measures, and task types, other potentially influential factors such as individual differences in neuroplasticity or variations in motor learning protocols were not included because of data limitations. This highlights the broader challenge of balancing model complexity with available data to avoid overfitting or biased conclusions in meta-analyses. Another key limitation relates to the complex and incompletely understood mechanisms underlying how tDCS modulates motor performance. Although many studies included in our review frame their protocols under the assumption that placing the anode over a target area, such as M1, would lead to modulation of the activity in that particular area, this assumption does not fully capture the actual electric field distribution. Computational modeling has consistently shown that with conventional electrode positions (such as M1 - supraorbital region), the peak electric field intensity often occurs between the anode and cathode rather than directly beneath the anode. This raises questions about whether the target region receives the strongest electric field [69], [70]. Our analyses did not account for the specific placement of the cathode electrode, which could have significant effects on both the direction and intensity of the induced E-field, thereby introducing an important limitation. Furthermore, it remains uncertain whether the magnitude of the local E-field directly translates to greater behavioral effects. The assumption of a simple dose–response relationship, where a stronger electric field leads to greater improvements in motor outcomes, is not consistently supported by the literature [71]. Finally, it is increasingly recognized that the concurrent application of tDCS with motor practice may promote “functional targeting,” whereby the modulation of brain activity occurs primarily within the networks engaged in the ongoing task, rather than at the anatomical site beneath the electrodes [62], [65], [72]. Our results demonstrate that stimulation promotes online learning. This effect did not appear to depend on the stimulation site, further complicating the direct interpretation of how network-level modulation influences behavioral outcomes.

### 4.6 | Perspectives

This research area continues to expand, driven by a dual focus: uncovering the behavioral effects of tDCS and elucidating its underlying neurophysiological mechanisms. Our findings highlight the favorable effects of tDCS on online motor learning, particularly when targeting the primary motor cortex, suggesting that targeting this region may hold potential for enhancing skill acquisition. Notably, the effects of tDCS appear to be amplified when applied during multiple training sessions, underscoring the importance of repeated stimulation sessions and the repeated interaction between motor practice and stimulation. From a practical standpoint, this suggests that concurrent stimulation and motor practice may be incorporated as a tool in everyday training contexts rather than relying on single-session applications to achieve greater and more sustained effects.

Future studies should carefully consider the design of both stimulation and motor learning paradigms. For instance, the interaction between stimulation intensity, duration, and task characteristics remains a critical area for further exploration to optimize the efficacy. Additionally, while concerns persist about whether the induction of weak electric fields, approximately 0.5 V/m within the brain is sufficient to modulate neuronal function meaningfully [61], [73], accumulating evidence from behavioral and neurophysiological studies suggests otherwise. Nevertheless, rigorous investigations using advanced methodologies such as computational modeling and high-resolution imaging are essential to bridge the gap between theoretical predictions and empirical findings. Furthermore, computational modeling and simulations can provide insights into which brain areas are affected by electric fields, thereby optimizing TES applications [70], [74]. By addressing these considerations, future research can contribute to refining stimulation protocols, advancing our understanding of the mechanisms underlying motor learning, and maximizing the translational potential of tDCS in both healthy individuals and clinical populations.

## Author Contributions

Conceptualization: A.L.N., J.L.-J., A.N.K. Methodology: A.L.N., J.L.-J. Validation: A.L.N., J.L.-J., A.N.K. Formal analysis: A.L.N. Investigation: A.L.N., M.I.G., L.J. Writing – original draft: A.L.N., J.L.-J., A.N.K. Writing – review and editing: A.L.N., J.L.-J., A.N.K., M.I.G., L.J., J.R.B., M.M.B., L.C., Visualization: A.L.N. Supervision: J.L.-J., A.N.K., L.C. Funding acquisition: A.L.N., J.L.-J.

## Acknowledgements

A.L.N. and J.L.-J. are supported by a grant from Team Danmark through the Novo Nordisk Foundation under Grant Number NNF.22SA0078293.

## Data Availability Statement

The data that support the findings of this study are available from the corresponding author upon reasonable request.

## References

[1] D. L. Adkins, J. Boychuk, M. S. Remple, and J. A. Kleim, “Motor training induces experience-specific patterns of plasticity across motor cortex and spinal cord,” J. Appl. Physiol., vol. 101, no. 6, pp. 1776–1782, Dec. 2006, doi: 10.1152/japplphysiol.00515.2006.

[2] M. S. Rioult-Pedotti, D. Friedman, and J. P. Donoghue, “Learning-induced LTP in neocortex,” Science, vol. 290, no. 5491, pp. 533–536, Oct. 2000, doi: 10.1126/science.290.5491.533.

[3] S. S. Kantak and C. J. Winstein, “Learning–performance distinction and memory processes for motor skills: A focused review and perspective,” Behav. Brain Res., vol. 228, no. 1, pp. 219–231, Mar. 2012, doi: 10.1016/j.bbr.2011.11.028.

[4] J. W. Krakauer, A. M. Hadjiosif, J. Xu, A. L. Wong, and A. M. Haith, “Motor Learning,” in Comprehensive Physiology, 1st ed., R. Terjung, Ed., Wiley, 2019, pp. 613–663. doi: 10.1002/cphy.c170043.

[5] A. Antal et al., “Non-invasive brain stimulation and neuroenhancement,” Clin. Neurophysiol. Pract., vol. 7, pp. 146–165, Jan. 2022, doi: 10.1016/j.cnp.2022.05.002.

[6] E. R. Buch et al., “Effects of tDCS on motor learning and memory formation: A consensus and critical position paper,” Clin. Neurophysiol., vol. 128, no. 4, pp. 589–603, Apr. 2017, doi: 10.1016/j.clinph.2017.01.004.

[7] A. R. Brunoni et al., “Clinical research with transcranial direct current stimulation (tDCS): Challenges and future directions,” Brain Stimulat., vol. 5, no. 3, pp. 175–195, Jul. 2012, doi: 10.1016/j.brs.2011.03.002.

[8] M. A. Nitsche and W. Paulus, “Excitability changes induced in the human motor cortex by weak transcranial direct current stimulation,” J. Physiol., vol. 527, no. Pt 3, pp. 633–639, Sep. 2000, doi: 10.1111/j.1469-7793.2000.t01-1-00633.x.

[9] A. Antal, K. Boros, C. Poreisz, L. Chaieb, D. Terney, and W. Paulus, “Comparatively weak after-effects of transcranial alternating current stimulation (tACS) on cortical excitability in humans,” Brain Stimulat., vol. 1, no. 2, pp. 97–105, Apr. 2008, doi: 10.1016/j.brs.2007.10.001.

[10] J. Fan, J. Voisin, M.-H. Milot, J. Higgins, and M.-H. Boudrias, “Transcranial direct current stimulation over multiple days enhances motor performance of a grip task,” Ann. Phys. Rehabil. Med., vol. 60, no. 5, pp. 329–333, Sep. 2017, doi: 10.1016/j.rehab.2017.07.001.

[11] J. M. Galea, A. Vazquez, N. Pasricha, J.-J. Orban de Xivry, and P. Celnik, “Dissociating the Roles of the Cerebellum and Motor Cortex during Adaptive Learning: The Motor Cortex Retains What the Cerebellum Learns,” Cereb. Cortex, vol. 21, no. 8, pp. 1761–1770, Aug. 2011, doi: 10.1093/cercor/bhq246.

[12] J. Reis et al., “Noninvasive cortical stimulation enhances motor skill acquisition over multiple days through an effect on consolidation,” Proc. Natl. Acad. Sci., vol. 106, no. 5, pp. 1590–1595, Feb. 2009, doi: 10.1073/pnas.0805413106.

[13] P. Sehatpour et al., “Network-level mechanisms underlying effects of transcranial direct current stimulation (tDCS) on visuomotor learning,” NeuroImage, vol. 223, p. 117311, Dec. 2020, doi: 10.1016/j.neuroimage.2020.117311.

[14] A. N. Guimarães, A. B. Porto, A. J. Marcori, G. M. Lage, L. R. Altimari, and V. H. Alves Okazaki, “Motor learning and tDCS: A systematic review on the dependency of the stimulation effect on motor task characteristics or tDCS assembly specifications,” Neuropsychologia, vol. 179, p. 108463, Jan. 2023, doi: 10.1016/j.neuropsychologia.2022.108463.

[15] P. Besson, M. Muthalib, C. De Vassoigne, J. Rothwell, and S. Perrey, “Effects of Multiple Sessions of Cathodal Priming and Anodal HD-tDCS on Visuo Motor Task Plateau Learning and Retention,” Brain Sci., vol. 10, no. 11, Art. no. 11, Nov. 2020, doi: 10.3390/brainsci10110875.

[16] Á. Foerster, A. Dutta, M.-F. Kuo, W. Paulus, and M. A. Nitsche, “Effects of anodal transcranial direct current stimulation over lower limb primary motor cortex on motor learning in healthy individuals,” Eur. J. Neurosci., vol. 47, no. 7, pp. 779–789, 2018, doi: 10.1111/ejn.13866.

[17] K. Rosenkranz, M. A. Nitsche, F. Tergau, and W. Paulus, “Diminution of training-induced transient motor cortex plasticity by weak transcranial direct current stimulation in the human,” Neurosci. Lett., vol. 296, no. 1, pp. 61–63, Dec. 2000, doi: 10.1016/S0304-3940(00)01621-9.

[18] E. Dayan and L. G. Cohen, “Neuroplasticity Subserving Motor Skill Learning,” Neuron, vol. 72, no. 3, pp. 443–454, Nov. 2011, doi: 10.1016/j.neuron.2011.10.008.

[19] H. K. Ballard, S. M. Eakin, T. Maldonado, and J. A. Bernard, “Using high-definition transcranial direct current stimulation to investigate the role of the dorsolateral prefrontal cortex in explicit sequence learning,” PloS One, vol. 16, no. 3, p. e0246849, 2021, doi: 10.1371/journal.pone.0246849.

[20] T. Kim, J. J. Buchanan, J. A. Bernard, and D. L. Wright, “Improving online and offline gain from repetitive practice using anodal tDCS at dorsal premotor cortex,” Npj Sci. Learn., vol. 6, no. 1, p. 31, Oct. 2021, doi: 10.1038/s41539-021-00109-4.

[21] N. Mizuguchi, T. Katayama, and K. Kanosue, “The Effect of Cerebellar Transcranial Direct Current Stimulation on A Throwing Task Depends on Individual Level of Task Performance,” Neuroscience, vol. 371, pp. 119–125, Feb. 2018, doi: 10.1016/j.neuroscience.2017.11.048.

[22] S. Waters-Metenier, M. Husain, T. Wiestler, and J. Diedrichsen, “Bihemispheric Transcranial Direct Current Stimulation Enhances Effector-Independent Representations of Motor Synergy and Sequence Learning,” J. Neurosci., vol. 34, no. 3, pp. 1037–1050, Jan. 2014, doi: 10.1523/JNEUROSCI.2282-13.2014.

[23] T. Yamaguchi et al., “Transcranial Alternating Current Stimulation of the Primary Motor Cortex after Skill Acquisition Improves Motor Memory Retention in Humans: A Double-Blinded Sham-Controlled Study,” Cereb. Cortex Commun., vol. 1, no. 1, p. tgaa047, Aug. 2020, doi: 10.1093/texcom/tgaa047.

[24] S. T. Grafton, E. Hazeltine, and R. Ivry, “Functional Mapping of Sequence Learning in Normal Humans,” J. Cogn. Neurosci., vol. 7, no. 4, pp. 497–510, Oct. 1995, doi: 10.1162/jocn.1995.7.4.497.

[25] A. N. Karabanov, G. B. Saturnino, A. Thielscher, and H. R. Siebner, “Can Transcranial Electrical Stimulation Localize Brain Function?,” Front. Psychol., vol. 10, 2019, Accessed: May 01, 2023. [Online]. Available: https://www.frontiersin.org/articles/10.3389/fpsyg.2019.00213

[26] S. J. Pelletier and F. Cicchetti, “Cellular and Molecular Mechanisms of Action of Transcranial Direct Current Stimulation: Evidence from In Vitro and In Vivo Models,” Int. J. Neuropsychopharmacol., vol. 18, no. 2, p. pyu047, Feb. 2015, doi: 10.1093/ijnp/pyu047.

[27] A. Bastani and S. Jaberzadeh, “Does anodal transcranial direct current stimulation enhance excitability of the motor cortex and motor function in healthy individuals and subjects with stroke: A systematic review and meta-analysis,” Clin. Neurophysiol., vol. 123, no. 4, pp. 644–657, Apr. 2012, doi: 10.1016/j.clinph.2011.08.029.

[28] A. Rahman et al., “Cellular effects of acute direct current stimulation: somatic and synaptic terminal effects,” J. Physiol., vol. 591, no. 10, pp. 2563–2578, 2013, doi: 10.1113/jphysiol.2012.247171.

[29] T. Maldonado, T. B. Jackson, and J. A. Bernard, “Anodal cerebellar stimulation increases cortical activation: Evidence for cerebellar scaffolding of cortical processing,” Hum. Brain Mapp., vol. 44, no. 4, pp. 1666–1682, Mar. 2023, doi: 10.1002/hbm.26166.

[30] A. Watanabe et al., “Transcranial direct current stimulation to the left dorsolateral prefrontal cortex enhances early dexterity skills with the left non-dominant hand: a randomized controlled trial,” J. Transl. Med., vol. 21, no. 1, p. 143, Feb. 2023, doi: 10.1186/s12967-023-03989-9.

[31] M. J. Page et al., “The PRISMA 2020 statement: an updated guideline for reporting systematic reviews,” Syst. Rev., vol. 10, no. 1, p. 89, Dec. 2021, doi: 10.1186/s13643-021-01626-4.

[32] J. Popay et al., Guidance on the conduct of narrative synthesis in systematic reviews: A product from the ESRC Methods Programme. 2006. doi: 10.13140/2.1.1018.4643.

[33] J. Reis, J. T. Fischer, G. Prichard, C. Weiller, L. G. Cohen, and B. Fritsch, “Time- but Not Sleep-Dependent Consolidation of tDCS-Enhanced Visuomotor Skills,” Cereb. Cortex, vol. 25, no. 1, pp. 109–117, Jan. 2015, doi: 10.1093/cercor/bht208.

[34] J. P. T. Higgins et al., Cochrane Handbook for Systematic Reviews of Interventions. John Wiley & Sons, 2019.

[35] R. A. Ince, A. T. Paton, J. W. Kay, and P. G. Schyns, “Bayesian inference of population prevalence,” eLife, vol. 10, p. e62461, Oct. 2021, doi: 10.7554/eLife.62461.

[36] Michael Borenstein, L. V. Hedges, J. P. T. Higgins, and H. R. Rothstein, Introduction to meta-analysis, Second edition. Hoboken, NJ: John Wiley & Sons, Inc., 2021.

[37] J. Cohen, Statistical power analysis for the behavioral sciences, 2nd ed. Hillsdale, N.J: L. Erlbaum Associates, 1988.

[38] J. A. Durlak, “How to Select, Calculate, and Interpret Effect Sizes,” J. Pediatr. Psychol., vol. 34, no. 9, pp. 917–928, Oct. 2009, doi: 10.1093/jpepsy/jsp004.

[39] T. A. Paterson, P. D. Harms, P. Steel, and M. Credé, “An Assessment of the Magnitude of Effect Sizes: Evidence From 30 Years of Meta-Analysis in Management,” J. Leadersh. Organ. Stud., vol. 23, no. 1, pp. 66–81, Feb. 2016, doi: 10.1177/1548051815614321.

[40] J. K. Kruschke, Ed., “Copyright,” in *Doing Bayesian Data Analysis (Second Edition)*, Boston: Academic Press, 2015, p. iv. doi: 10.1016/B978-0-12-405888-0.09998-0.

[41] R. McElreath, Statistical Rethinking: A Bayesian Course with Examples in R and Stan, Second edition. in Chapman & Hall/CRC Texts in Statistical Science. Milton: CRC Press, 2020. doi: 10.1201/9780429029608.

[42] B. Fernández-Castilla, L. Jamshidi, L. Declercq, S. N. Beretvas, P. Onghena, and W. Van den Noortgate, “The application of meta-analytic (multi-level) models with multiple random effects: A systematic review,” Behav. Res. Methods, vol. 52, no. 5, pp. 2031–2052, Oct. 2020, doi: 10.3758/s13428-020-01373-9.

[43] J. P. T. Higgins, S. G. Thompson, and D. J. Spiegelhalter, “A re-evaluation of random-effects meta-analysis,” J. R. Stat. Soc. Ser. A Stat. Soc., vol. 172, no. 1, p. 137, Jan. 2009, doi: 10.1111/j.1467-985X.2008.00552.x.

[44] D. R. Williams, P. Rast, and P.-C. Bürkner, “Bayesian Meta-Analysis with Weakly Informative Prior Distributions,” Jan. 10, 2018, OSF. doi: 10.31234/osf.io/7tbrm.

[45] L. V. Hedges and J. L. Vevea, “Fixed- and random-effects models in meta-analysis,” Psychol. Methods, vol. 3, no. 4, pp. 486–504, 1998, doi: 10.1037/1082-989X.3.4.486.

[46] P. Jylänki, J. Vanhatalo, and A. Vehtari, “Robust Gaussian Process Regression with a Student-t Likelihood,” J. Mach. Learn. Res., vol. 12, pp. 3227–3257, 2011.

[47] A. Gelman, et al., “Bayesian Workflow,” Nov. 03, 2020, *arXiv*: arXiv:2011.01808.doi: 10.48550/arXiv.2011.01808.

[48] A. Vehtari, A. Gelman, and J. Gabry, “Practical Bayesian model evaluation using leave-one-out cross-validation and WAIC,” Stat. Comput., vol. 27, no. 5, pp. 1413–1432, Sep. 2017, doi: 10.1007/s11222-016-9696-4.

[49] M. B. Mathur and T. J. VanderWeele, “Sensitivity analysis for publication bias in meta-analyses,” J. R. Stat. Soc. Ser. C Appl. Stat., vol. 69, no. 5, pp. 1091–1119, Nov. 2020, doi: 10.1111/rssc.12440.

[50] W. Viechtbauer, “Conducting Meta-Analyses in *R* with the **metafor** Package,” J. Stat. Softw., vol. 36, no. 3, 2010, doi: 10.18637/jss.v036.i03.

[51] P.-C. Bürkner, “**brms** : An *R* Package for Bayesian Multilevel Models Using *Stan*,” J. Stat. Softw., vol. 80, no. 1, 2017, doi: 10.18637/jss.v080.i01.

[52] B. Carpenter et al., “Stan: A Probabilistic Programming Language,” J. Stat. Softw., vol. 76, p. 1, 2017, doi: 10.18637/jss.v076.i01.

[53] G. B. Saturnino, O. Puonti, J. D. Nielsen, D. Antonenko, K. H. Madsen, and A. Thielscher, “SimNIBS 2.1: A Comprehensive Pipeline for Individualized Electric Field Modelling for Transcranial Brain Stimulation,” in Brain and Human Body Modeling: Computational Human Modeling at EMBC 2018, S. Makarov, M. Horner, and G. Noetscher, Eds., Cham: Springer International Publishing, 2019, pp. 3–25. doi: 10.1007/978-3-030-21293-3_1.

[54] Z. Mastrich and I. Hernandez, “Results everyone can understand: A review of common language effect size indicators to bridge the research-practice gap,” Health Psychol., vol. 40, no. 10, pp. 727–736, 2021, doi: 10.1037/hea0001112.

[55] J. K. Kruschke and T. M. Liddell, “The Bayesian New Statistics: Hypothesis testing, estimation, meta-analysis, and power analysis from a Bayesian perspective,” Psychon. Bull. Rev., vol. 25, no. 1, pp. 178–206, Feb. 2018, doi: 10.3758/s13423-016-1221-4.

[56] F. Hashemirad, M. Zoghi, P. B. Fitzgerald, and S. Jaberzadeh, “The effect of anodal transcranial direct current stimulation on motor sequence learning in healthy individuals: A systematic review and meta-analysis,” Brain Cogn., vol. 102, pp. 1–12, Feb. 2016, doi: 10.1016/j.bandc.2015.11.005.

[57] N. Kumari, D. Taylor, and N. Signal, “The Effect of Cerebellar Transcranial Direct Current Stimulation on Motor Learning: A Systematic Review of Randomized Controlled Trials,” Front. Hum. Neurosci., vol. 13, 2019, Accessed: May 24, 2023. [Online]. Available: https://www.frontiersin.org/articles/10.3389/fnhum.2019.00328

[58] B. Fritsch et al., “Direct Current Stimulation Promotes BDNF-Dependent Synaptic Plasticity: Potential Implications for Motor Learning,” Neuron, vol. 66, no. 2, pp. 198–204, Apr. 2010, doi: 10.1016/j.neuron.2010.03.035.

[59] Y. Wang et al., “Neural Mechanism Underlying Task-Specific Enhancement of Motor Learning by Concurrent Transcranial Direct Current Stimulation,” Neurosci. Bull., Jul. 2022, doi: 10.1007/s12264-022-00901-1.

[60] M. A. Nitsche et al., “Facilitation of implicit motor learning by weak transcranial direct current stimulation of the primary motor cortex in the human,” J. Cogn. Neurosci., vol. 15, no. 4, pp. 619–626, May 2003, doi: 10.1162/089892903321662994.

[61] A. Liu et al., “Immediate neurophysiological effects of transcranial electrical stimulation,” Nat. Commun., vol. 9, no. 1, p. 5092, Nov. 2018, doi: 10.1038/s41467-018-07233-7.

[62] E. Morya et al., “Beyond the target area: an integrative view of tDCS-induced motor cortex modulation in patients and athletes,” J. NeuroEngineering Rehabil., vol. 16, no. 1, p. 141, Nov. 2019, doi: 10.1186/s12984-019-0581-1.

[63] M. Bikson et al., “Effects of uniform extracellular DC electric fields on excitability in rat hippocampal slices in vitro,” J. Physiol., vol. 557, no. 1, pp. 175–190, 2004, doi: 10.1113/jphysiol.2003.055772.

[64] B. Lafon, A. Rahman, M. Bikson, and L. C. Parra, “Direct Current Stimulation Alters Neuronal Input/Output Function,” Brain Stimulat., vol. 10, no. 1, pp. 36–45, Jan. 2017, doi: 10.1016/j.brs.2016.08.014.

[65] M. P. Jackson et al., “Animal models of transcranial direct current stimulation: Methods and mechanisms,” Clin. Neurophysiol., vol. 127, no. 11, pp. 3425–3454, Nov. 2016, doi: 10.1016/j.clinph.2016.08.016.

[66] S. Vogeti, C. Boetzel, and C. S. Herrmann, “Entrainment and Spike-Timing Dependent Plasticity – A Review of Proposed Mechanisms of Transcranial Alternating Current Stimulation,” Front. Syst. Neurosci., vol. 16, p. 827353, Feb. 2022, doi: 10.3389/fnsys.2022.827353.

[67] K. Hu, R. Wan, Y. Liu, M. Niu, J. Guo, and F. Guo, “Effects of transcranial alternating current stimulation on motor performance and motor learning for healthy individuals: A systematic review and meta-analysis,” Front. Physiol., vol. 13, 2022, Accessed: Sep. 11, 2023. [Online]. Available: https://www.frontiersin.org/articles/10.3389/fphys.2022.1064584

[68] M. Rostami et al., “Determining the corticospinal, intracortical and motor function responses to transcranial alternating current stimulation of the motor cortex in healthy adults: A systematic review and *meta*-analysis,” Brain Res., vol. 1822, p. 148650, Jan. 2024, doi: 10.1016/j.brainres.2023.148650.

[69] A. Opitz, W. Paulus, S. Will, A. Antunes, and A. Thielscher, “Determinants of the electric field during transcranial direct current stimulation,” NeuroImage, vol. 109, pp. 140–150, Apr. 2015, doi: 10.1016/j.neuroimage.2015.01.033.

[70] G. B. Saturnino, A. Antunes, and A. Thielscher, “On the importance of electrode parameters for shaping electric field patterns generated by tDCS,” NeuroImage, vol. 120, pp. 25–35, Oct. 2015, doi: 10.1016/j.neuroimage.2015.06.067.

[71] Z. Esmaeilpour et al., “Incomplete evidence that increasing current intensity of tDCS boosts outcomes,” Brain Stimulat., vol. 11, no. 2, pp. 310–321, 2018, doi: 10.1016/j.brs.2017.12.002.

[72] M. Bikson and A. Rahman, “Origins of specificity during tDCS: anatomical, activity-selective, and input-bias mechanisms,” Front. Hum. Neurosci., vol. 7, 2013, Accessed: Jul. 19, 2023. [Online]. Available: https://www.frontiersin.org/articles/10.3389/fnhum.2013.00688

[73] J. Modolo, Y. Denoyer, F. Wendling, and P. Benquet, “Physiological effects of low-magnitude electric fields on brain activity: Advances from *in vitro*, *in vivo* and *in silico* models,” Curr. Opin. Biomed. Eng., vol. 8, pp. 38–44, Dec. 2018, doi: 10.1016/j.cobme.2018.09.006.

[74] D. Antonenko et al., “Towards precise brain stimulation: Is electric field simulation related to neuromodulation?,” Brain Stimulat., vol. 12, no. 5, pp. 1159–1168, Sep. 2019, doi: 10.1016/j.brs.2019.03.072.

[75] A. Sriraman, T. Oishi, and S. Madhavan, “Timing-dependent priming effects of tDCS on ankle motor skill learning,” Brain Res., vol. 1581, pp. 23–29, Sep. 2014, doi: 10.1016/j.brainres.2014.07.021.

[76] T. Zandonai, M. Bertucco, N. Graziani, V. Montani, and P. Cesari, “Transcranial direct current stimulation (tDCS) modulates motor execution in a limb reaching task,” Eur. J. Neurosci., vol. 56, no. 4, pp. 4445–4454, 2022, doi: 10.1111/ejn.15756.

[77] O. Seidel and P. Ragert, “Effects of Transcranial Direct Current Stimulation of Primary Motor Cortex on Reaction Time and Tapping Performance: A Comparison Between Athletes and Non-athletes,” Front. Hum. Neurosci., vol. 13, p. 103, Apr. 2019, doi: 10.3389/fnhum.2019.00103.

[78] B. Shah, T. T. Nguyen, and S. Madhavan, “Polarity Independent Effects of Cerebellar tDCS on Short Term Ankle Visuomotor Learning,” Brain Stimulat., vol. 6, no. 6, pp. 966–968, Nov. 2013, doi: 10.1016/j.brs.2013.04.008.

[79] T. Apolinário-Souza et al., “The primary motor cortex is associated with learning the absolute, but not relative, timing dimension of a task: A tDCS study,” Physiol. Behav., vol. 160, pp. 18–25, Jun. 2016, doi: 10.1016/j.physbeh.2016.03.025.

[80] A. Azarpaikan, H. R. T. Torbati, M. Sohrabi, R. Boostani, and M. Ghoshoni, “Timing-Dependent Priming Effects of Anodal tDCS on Two-Hand Coordination,” J. Psychophysiol., vol. 34, no. 4, pp. 224–234, Oct. 2020, doi: 10.1027/0269-8803/a000250.

[81] A. Azarpaikan, H. R. Taherii Torbati, M. Sohrabi, R. Boostani, and M. Ghoshuni, “The Effect of Parietal and Cerebellar Transcranial Direct Current Stimulation on Bimanual Coordinated Adaptive Motor Learning,” J. Psychophysiol., vol. 35, no. 1, pp. 1–14, Jan. 2021, doi: 10.1027/0269-8803/a000254.

[82] J. Ashcroft, R. Patel, A. J. Woods, A. Darzi, H. Singh, and D. R. Leff, “Prefrontal transcranial direct-current stimulation improves early technical skills in surgery,” Brain Stimulat., vol. 13, no. 6, pp. 1834–1841, 2020, doi: 10.1016/j.brs.2020.10.013.

[83] J. Ashcroft, R. Patel, A. J. Woods, A. Darzi, H. Singh, and D. R. Leff, “Prefrontal transcranial direct-current stimulation improves early technical skills in surgery,” Brain Stimul. Basic Transl. Clin. Res. Neuromodulation, vol. 13, no. 6, pp. 1834–1841, Nov. 2020, doi: 10.1016/j.brs.2020.10.013.

[84] A. Azarpaikan, H. R. T. Torbati, M. Sohrabi, R. Boostani, and M. Ghoshoni, “Timing-Dependent Priming Effects of Anodal tDCS on Two-Hand Coordination,” J. Psychophysiol., vol. 34, no. 4, pp. 224–234, Oct. 2020, doi: 10.1027/0269-8803/a000250.

[85] A. Azarpaikan, H. R. Taherii Torbati, M. Sohrabi, R. Boostani, and M. Ghoshuni, “The Effect of Parietal and Cerebellar Transcranial Direct Current Stimulation on Bimanual Coordinated Adaptive Motor Learning,” J. Psychophysiol., vol. 35, no. 1, pp. 1–14, Jan. 2021, doi: 10.1027/0269-8803/a000254.

[86] H. K. Ballard, S. M. Eakin, T. Maldonado, and J. A. Bernard, “Using high-definition transcranial direct current stimulation to investigate the role of the dorsolateral prefrontal cortex in explicit sequence learning,” PLOS ONE, vol. 16, no. 3, p. e0246849, Mar. 2021, doi: 10.1371/journal.pone.0246849.

[87] H. K. Ballard, J. R. M. Goen, T. Maldonado, and J. A. Bernard, “Effects of cerebellar transcranial direct current stimulation on the cognitive stage of sequence learning,” J. Neurophysiol., vol. 122, no. 2, pp. 490–499, Aug. 2019, doi: 10.1152/jn.00036.2019.

[88] G. Cantarero et al., “Cerebellar Direct Current Stimulation Enhances On-Line Motor Skill Acquisition through an Effect on Accuracy,” J. Neurosci., vol. 35, no. 7, pp. 3285–3290, Feb. 2015, doi: 10.1523/JNEUROSCI.2885-14.2015.

[89] P. Ciechanski et al., “Electroencephalography correlates of transcranial direct-current stimulation enhanced surgical skill learning: A replication and extension study,” Brain Res., vol. 1725, p. 146445, Dec. 2019, doi: 10.1016/j.brainres.2019.146445.

[90] M. L. Cox et al., “Utilizing transcranial direct current stimulation to enhance laparoscopic technical skills training: A randomized controlled trial,” Brain Stimulat., vol. 13, no. 3, pp. 863–872, May 2020, doi: 10.1016/j.brs.2020.03.009.

[91] F. Ehsani, A. H. Bakhtiary, S. Jaberzadeh, A. Talimkhani, and A. Hajihasani, “Differential effects of primary motor cortex and cerebellar transcranial direct current stimulation on motor learning in healthy individuals: A randomized double-blind sham-controlled study,” Neurosci. Res., vol. 112, pp. 10–19, Nov. 2016, doi: 10.1016/j.neures.2016.06.003.

[92] J. Fan, J. Voisin, M.-H. Milot, J. Higgins, and M.-H. Boudrias, “Transcranial direct current stimulation over multiple days enhances motor performance of a grip task,” Ann. Phys. Rehabil. Med., vol. 60, no. 5, pp. 329–333, Sep. 2017, doi: 10.1016/j.rehab.2017.07.001.

[93] J. Fan, J. Voisin, M.-H. Milot, J. Higgins, and M.-H. Boudrias, “Transcranial direct current stimulation over multiple days enhances motor performance of a grip task,” Ann. Phys. Rehabil. Med., vol. 60, no. 5, pp. 329–333, Sep. 2017, doi: 10.1016/j.rehab.2017.07.001.

[94] Á. Foerster, A. Dutta, M. Kuo, W. Paulus, and M. A. Nitsche, “Effects of anodal transcranial direct current stimulation over lower limb primary motor cortex on motor learning in healthy individuals,” Eur. J. Neurosci., vol. 47, no. 7, pp. 779–789, Apr. 2018, doi: 10.1111/ejn.13866.

[95] B. Fritsch et al., “Direct Current Stimulation Promotes BDNF-Dependent Synaptic Plasticity: Potential Implications for Motor Learning,” Neuron, vol. 66, no. 2, pp. 198–204, Apr. 2010, doi: 10.1016/j.neuron.2010.03.035.

[96] Y. Gao et al., “Decreasing the Surgical Errors by Neurostimulation of Primary Motor Cortex and the Associated Brain Activation via Neuroimaging,” Front. Neurosci., vol. 15, 2021, Accessed: Sep. 24, 2023. [Online]. Available: https://www.frontiersin.org/articles/10.3389/fnins.2021.651192

[97] Y. Gao et al., “Decreasing the Surgical Errors by Neurostimulation of Primary Motor Cortex and the Associated Brain Activation via Neuroimaging,” Front. Neurosci., vol. 15, 2021, Accessed: Sep. 24, 2023. [Online]. Available: https://www.frontiersin.org/articles/10.3389/fnins.2021.651192

[98] B. Greeley, J. S. Barnhoorn, W. B. Verwey, and R. D. Seidler, “Multi-session Transcranial Direct Current Stimulation Over Primary Motor Cortex Facilitates Sequence Learning, Chunking, and One Year Retention,” Front. Hum. Neurosci., vol. 14, 2020, Accessed: Sep. 24, 2023. [Online]. Available: https://www.frontiersin.org/articles/10.3389/fnhum.2020.00075

[99] Z. Hadi, A. Umbreen, M. N. Anwar, and M. S. Navid, “The effects of unilateral transcranial direct current stimulation on unimanual laparoscopic peg-transfer task,” Brain Res., vol. 1771, p. 147656, Nov. 2021, doi: 10.1016/j.brainres.2021.147656.

[100] F. Hashemirad, P. B. Fitzgerald, M. Zoghi, and S. Jaberzadeh, “Single-Session Anodal tDCS with Small-Size Stimulating Electrodes Over Frontoparietal Superficial Sites Does Not Affect Motor Sequence Learning,” Front. Hum. Neurosci., vol. 11, 2017, Accessed: Sep. 24, 2023. [Online]. Available: https://www.frontiersin.org/articles/10.3389/fnhum.2017.00153

[101] G. Hsu, A. D. Shereen, L. G. Cohen, and L. C. Parra, “Robust enhancement of motor sequence learning with 4 mA transcranial electric stimulation,” Brain Stimulat., vol. 16, no. 1, pp. 56–67, Jan. 2023, doi: 10.1016/j.brs.2022.12.011.

[102] A. Iannone, I. Santiago, S. T. Ajao, J. Brasil-Neto, J. C. Rothwell, and D. A. Spampinato, “Comparing the effects of focal and conventional tDCS on motor skill learning: A proof of principle study,” Neurosci. Res., vol. 178, pp. 83–86, May 2022, doi: 10.1016/j.neures.2022.01.006.

[103] B. J. Jongkees, M. A. Immink, O. D. Boer, F. Yavari, M. A. Nitsche, and L. S. Colzato, “The Effect of Cerebellar tDCS on Sequential Motor Response Selection,” The Cerebellum, vol. 18, no. 4, pp. 738–749, Aug. 2019, doi: 10.1007/s12311-019-01029-1.

[104] N.-G. Jo, G.-W. Kim, Y. H. Won, S.-H. Park, J.-H. Seo, and M.-H. Ko, “Timing-Dependent Effects of Transcranial Direct Current Stimulation on Hand Motor Function in Healthy Individuals: A Randomized Controlled Study,” Brain Sci., vol. 11, no. 10, p. 1325, Oct. 2021, doi: 10.3390/brainsci11101325.

[105] E. Kaminski, M. Engelhardt, M. Hoff, C. Steele, A. Villringer, and P. Ragert, “TDCS effects on pointing task learning in young and old adults,” Sci. Rep., vol. 11, no. 1, Art. no. 1, Feb. 2021, doi: 10.1038/s41598-021-82275-4.

[106] E. Kaminski et al., “Transcranial direct current stimulation (tDCS) over primary motor cortex leg area promotes dynamic balance task performance,” Clin. Neurophysiol., vol. 127, no. 6, pp. 2455–2462, Jun. 2016, doi: 10.1016/j.clinph.2016.03.018.

[107] S. Karok and A. G. Witney, “Enhanced Motor Learning Following Task-Concurrent Dual Transcranial Direct Current Stimulation,” PLoS ONE, vol. 8, no. 12, p. e85693, Dec. 2013, doi: 10.1371/journal.pone.0085693.

[108] N. Kunaratnam et al., “Transcranial direct current stimulation leads to faster acquisition of motor skills, but effects are not maintained at retention,” PloS One, vol. 17, no. 9, p. e0269851, 2022, doi: 10.1371/journal.pone.0269851.

[109] M. Liebrand et al., “Beneficial effects of cerebellar tDCS on motor learning are associated with altered putamen-cerebellar connectivity: A simultaneous tDCS-fMRI study,” NeuroImage, vol. 223, p. 117363, Dec. 2020, doi: 10.1016/j.neuroimage.2020.117363.

[110] P. G. Lindberg, M. Verneau, Q. L. Boterff, M. Cuenca-Maia, J.-C. Baron, and M. A. Maier, “Age- and task-dependent effects of cerebellar tDCS on manual dexterity and motor learning–A preliminary study,” Neurophysiol. Clin., vol. 52, no. 5, pp. 354–365, Oct. 2022, doi: 10.1016/j.neucli.2022.07.006.

[111] V. Lopez-Alonso et al., “A Preliminary Comparison of Motor Learning Across Different Non-invasive Brain Stimulation Paradigms Shows No Consistent Modulations,” Front. Neurosci., vol. 12, 2018, Accessed: Sep. 24, 2023. [Online]. Available: https://www.frontiersin.org/articles/10.3389/fnins.2018.00253

[112] J. A. G. Lum et al., “Transcranial direct current stimulation enhances retention of a second (but not first) order conditional visuo-motor sequence,” Brain Cogn., vol. 127, pp. 34–41, Nov. 2018, doi: 10.1016/j.bandc.2018.09.006.

[113] P. Maceira-Elvira et al., “Dissecting motor skill acquisition: Spatial coordinates take precedence,” Sci. Adv., vol. 8, no. 29, p. eabo3505, Jul. 2022, doi: 10.1126/sciadv.abo3505.

[114] C. Saucedo Marquez, X. Zhang, S. Swinnen, R. Meesen, and N. Wenderoth, “Task-Specific Effect of Transcranial Direct Current Stimulation on Motor Learning,” Front. Hum. Neurosci., vol. 7, 2013, Accessed: Sep. 24, 2023. [Online]. Available: https://www.frontiersin.org/articles/10.3389/fnhum.2013.00333

[115] R. A. Mooney, J. Cirillo, and W. D. Byblow, “Neurophysiological mechanisms underlying motor skill learning in young and older adults,” Exp. Brain Res., vol. 237, no. 9, pp. 2331–2344, Sep. 2019, doi: 10.1007/s00221-019-05599-8.

[116] C. Nguemeni, A. Stiehl, S. Hiew, and D. Zeller, “No Impact of Cerebellar Anodal Transcranial Direct Current Stimulation at Three Different Timings on Motor Learning in a Sequential Finger-Tapping Task,” Front. Hum. Neurosci., vol. 15, 2021, Accessed: Sep. 24, 2023. [Online]. Available: https://www.frontiersin.org/articles/10.3389/fnhum.2021.631517

[117] N. H. Pixa, A. Berger, F. Steinberg, and M. Doppelmayr, “Parietal, but Not Motor Cortex, HD-atDCS Deteriorates Learning Transfer of a Complex Bimanual Coordination Task,” J. Cogn. Enhanc., vol. 3, no. 1, pp. 111–123, Mar. 2019, doi: 10.1007/s41465-018-0088-x.

[118] N. H. Pixa, F. Steinberg, and M. Doppelmayr, “Effects of High-Definition Anodal Transcranial Direct Current Stimulation Applied Simultaneously to Both Primary Motor Cortices on Bimanual Sensorimotor Performance,” Front. Behav. Neurosci., vol. 11, 2017, Accessed: Sep. 24, 2023. [Online]. Available: https://www.frontiersin.org/articles/10.3389/fnbeh.2017.00130

[119] G. Prichard, C. Weiller, B. Fritsch, and J. Reis, “Effects of Different Electrical Brain Stimulation Protocols on Subcomponents of Motor Skill Learning,” Brain Stimulat., vol. 7, no. 4, pp. 532–540, Jul. 2014, doi: 10.1016/j.brs.2014.04.005.

[120] R. K. Raw, R. J. Allen, M. Mon-Williams, and R. M. Wilkie, “Motor Sequence Learning in Healthy Older Adults Is Not Necessarily Facilitated by Transcranial Direct Current Stimulation (tDCS),” Geriatr. Basel Switz., vol. 1, no. 4, p. 32, Dec. 2016, doi: 10.3390/geriatrics1040032.

[121] J. Reis, J. T. Fischer, G. Prichard, C. Weiller, L. G. Cohen, and B. Fritsch, “Time-but Not Sleep-Dependent Consolidation of tDCS-Enhanced Visuomotor Skills,” Cereb. Cortex, vol. 25, no. 1, pp. 109–117, Jan. 2015, doi: 10.1093/cercor/bht208.

[122] J. Reis et al., “Noninvasive cortical stimulation enhances motor skill acquisition over multiple days through an effect on consolidation,” Proc. Natl. Acad. Sci., vol. 106, no. 5, pp. 1590–1595, Feb. 2009, doi: 10.1073/pnas.0805413106.

[123] G. N. Rivera-Urbina, A. Molero-Chamizo, and M. A. Nitsche, “Discernible effects of tDCS over the primary motor and posterior parietal cortex on different stages of motor learning,” Brain Struct. Funct., vol. 227, no. 3, pp. 1115–1131, Apr. 2022, doi: 10.1007/s00429-021-02451-0.

[124] A. Sánchez-Kuhn, C. Pérez-Fernández, M. Moreno, P. Flores, and F. Sánchez-Santed, “Differential Effects of Transcranial Direct Current Stimulation (tDCS) Depending on Previous Musical Training,” Front. Psychol., vol. 9, 2018, Accessed: Sep. 24, 2023. [Online]. Available: https://www.frontiersin.org/articles/10.3389/fpsyg.2018.01465

[125] H. M. Schambra, M. Abe, D. A. Luckenbaugh, J. Reis, J. W. Krakauer, and L. G. Cohen, “Probing for hemispheric specialization for motor skill learning: a transcranial direct current stimulation study,” J. Neurophysiol., vol. 106, no. 2, pp. 652–661, Aug. 2011, doi: 10.1152/jn.00210.2011.

[126] M. Sevilla-Sanchez, T. Hortobágyi, N. Fogelson, E. Iglesias-Soler, E. Carballeira, and M. Fernandez-del-Olmo, “Small Enhancement of Bimanual Typing Performance after 20 Sessions of tDCS in Healthy Young Adults,” Neuroscience, vol. 466, pp. 26–35, Jul. 2021, doi: 10.1016/j.neuroscience.2021.05.001.

[127] “Shimizu et al. - 2017 - The impact of cerebellar transcranial direct curre.pdf.” Accessed: Sep. 24, 2023. [Online]. Available: https://regroup-production.s3.amazonaws.com/documents/ReviewReference/776621757/11.pdf?response-content-type=application%2Fpdf&X-Amz-Algorithm=AWS4-HMAC-SHA256&X-Amz-Credential=AKIAYSFKCAWYQ4D5IUHG%2F20230924%2Fus-east-1%2Fs3%2Faws4_request&X-Amz-Date=20230924T142851Z&X-Amz-Expires=604800&X-Amz-SignedHeaders=host&X-Amz-Signature=8d78ee7df90d9d1e0c2202680d497a205b6ee14107908b3245c473d18a02d81a

[128] R. E. Shimizu, A. D. Wu, J. K. Samra, and B. J. Knowlton, “The impact of cerebellar transcranial direct current stimulation (tDCS) on learning fine-motor sequences,” Philos. Trans. R. Soc. B Biol. Sci., vol. 372, no. 1711, p. 20160050, Jan. 2017, doi: 10.1098/rstb.2016.0050.

[129] A. Talimkhani, I. Abdollahi, M. A. Mohseni-Bandpei, F. Ehsani, S. Khalili, and S. Jaberzadeh, “Differential Effects of Unihemispheric Concurrent Dual-site and Conventional Primary Motor Cortex Transcranial Direct Current Stimulation on Motor Sequence Learning in Healthy Individuals: A Randomized Sham-Controlled Study,” *Basic Clin*. Neurosci. J., Oct. 2018, doi: 10.32598/bcn.9.10.350.

[130] A. Talimkhani, I. Abdollahi, M. Zoghi, E. Talebi Ghane, and S. Jaberzadeh, “The Effects of Unihemispheric Concurrent Dual-Site Transcranial Direct Current Stimulation on Motor Sequence Learning in Healthy Individuals: A Randomized, Clinical Trial,” Iran. Red Crescent Med. J., vol. 20, no. 7, May 2018, doi: 10.5812/ircmj.64147.

[131] S.-C. Tseng, S.-H. Chang, K. M. Hoerth, A.-T. A. Nguyen, and D. Perales, “Anodal Transcranial Direct Current Stimulation Enhances Retention of Visuomotor Stepping Skills in Healthy Adults,” Front. Hum. Neurosci., vol. 14, 2020, Accessed: Sep. 24, 2023. [Online]. Available: https://www.frontiersin.org/articles/10.3389/fnhum.2020.00251

[132] S. Waters-Metenier, M. Husain, T. Wiestler, and J. Diedrichsen, “Bihemispheric Transcranial Direct Current Stimulation Enhances Effector-Independent Representations of Motor Synergy and Sequence Learning,” J. Neurosci., vol. 34, no. 3, pp. 1037–1050, Jan. 2014, doi: 10.1523/JNEUROSCI.2282-13.2014.

[133] P. Ciechanski et al., “Effects of Transcranial Direct-Current Stimulation on Neurosurgical Skill Acquisition: A Randomized Controlled Trial,” World Neurosurg., vol. 108, pp. 876–884.e4, Dec. 2017, doi: 10.1016/j.wneu.2017.08.123.

[134] P. Ciechanski et al., “Effects of transcranial direct-current stimulation on laparoscopic surgical skill acquisition,” BJS Open, vol. 2, no. 2, pp. 70–78, Apr. 2018, doi: 10.1002/bjs5.43.

[135] A. Tiksnadi, T. Murakami, W. Wiratman, H. Matsumoto, and Y. Ugawa, “Direct comparison of efficacy of the motor cortical plasticity induction and the interindividual variability between TBS and QPS,” Brain Stimulat., vol. 13, no. 6, pp. 1824–1833, Nov. 2020, doi: 10.1016/j.brs.2020.10.014.

[136] “TDCS of the Primary Motor Cortex: Learning the Absolute Dimension of a Complex Motor Task.” Accessed: Feb. 19, 2024. [Online]. Available: https://www.tandfonline.com/doi/epdf/10.1080/00222895.2020.1792823?needAccess=true

[137] F. F. Zhu, A. Y. Yeung, J. M. Poolton, T. M. C. Lee, G. K. K. Leung, and R. S. W. Masters, “Cathodal Transcranial Direct Current Stimulation Over Left Dorsolateral Prefrontal Cortex Area Promotes Implicit Motor Learning in a Golf Putting Task,” Brain Stimulat., vol. 8, no. 4, pp. 784–786, Jul. 2015, doi: 10.1016/j.brs.2015.02.005.

[138] F. F. Zhu, A. Y. Yeung, J. M. Poolton, T. M. C. Lee, G. K. K. Leung, and R. S. W. Masters, “Cathodal Transcranial Direct Current Stimulation Over Left Dorsolateral Prefrontal Cortex Area Promotes Implicit Motor Learning in a Golf Putting Task,” Brain Stimulat., vol. 8, no. 4, pp. 784–786, Jul. 2015, doi: 10.1016/j.brs.2015.02.005.

[139] S. Karok, D. Fletcher, and A. G. Witney, “Task-specificity of unilateral anodal and dual-M1 tDCS effects on motor learning,” Neuropsychologia, vol. 94, pp. 84–95, Jan. 2017, doi: 10.1016/j.neuropsychologia.2016.12.002.

[140] T. Kim, H. Kim, and D. L. Wright, “Improving consolidation by applying anodal transcranial direct current stimulation at primary motor cortex during repetitive practice,” Neurobiol. Learn. Mem., vol. 178, p. 107365, Feb. 2021, doi: 10.1016/j.nlm.2020.107365.

[141] T. Kim and D. L. Wright, “Transcranial Direct Current Stimulation of Supplementary Motor Region Impacts the Effectiveness of Interleaved and Repetitive Practice Schedules for Retention of Motor Skills,” Neuroscience, vol. 435, pp. 58–72, May 2020, doi: 10.1016/j.neuroscience.2020.03.043.

[142] A. W. Meek, D. Greenwell, B. Poston, and Z. A. Riley, “Anodal tDCS accelerates on-line learning of dart throwing,” Neurosci. Lett., vol. 764, p. 136211, Nov. 2021, doi: 10.1016/j.neulet.2021.136211.

[143] G. Naros et al., “Enhanced motor learning with bilateral transcranial direct current stimulation: Impact of polarity or current flow direction?,” Clin. Neurophysiol., vol. 127, no. 4, pp. 2119–2126, Apr. 2016, doi: 10.1016/j.clinph.2015.12.020.

[144] N. H. Pixa, F. Steinberg, and M. Doppelmayr, “High-definition transcranial direct current stimulation to both primary motor cortices improves unimanual and bimanual dexterity,” Neurosci. Lett., vol. 643, pp. 84–88, Mar. 2017, doi: 10.1016/j.neulet.2017.02.033.

[145] P. Sehatpour et al., “Network-level mechanisms underlying effects of transcranial direct current stimulation (tDCS) on visuomotor learning,” NeuroImage, vol. 223, p. 117311, Dec. 2020, doi: 10.1016/j.neuroimage.2020.117311.

[146] R. L. S. Summers, M. Chen, A. Hatch, and T. J. Kimberley, “Cerebellar Transcranial Direct Current Stimulation Modulates Corticospinal Excitability During Motor Training,” Front. Hum. Neurosci., vol. 12, p. 118, Apr. 2018, doi: 10.3389/fnhum.2018.00118.

[147] A. Voegtle, C. Terlutter, K. Nikolai, A. Farahat, H. Hinrichs, and C. M. Sweeney-Reed, “Suppression of Motor Sequence Learning and Execution Through Anodal Cerebellar Transcranial Electrical Stimulation,” The Cerebellum, vol. 22, no. 6, pp. 1152–1165, Dec. 2023, doi: 10.1007/s12311-022-01487-0.

[148] H. Vollmann et al., “Anodal transcranial direct current stimulation (tDCS) over supplementary motor area (SMA) but not pre-SMA promotes short-term visuomotor learning,” Brain Stimulat., vol. 6, no. 2, pp. 101–107, Mar. 2013, doi: 10.1016/j.brs.2012.03.018.

[149] S. Waters, T. Wiestler, and J. Diedrichsen, “Cooperation Not Competition: Bihemispheric tDCS and fMRI Show Role for Ipsilateral Hemisphere in Motor Learning,” J. Neurosci., vol. 37, no. 31, pp. 7500–7512, Aug. 2017, doi: 10.1523/JNEUROSCI.3414-16.2017.

[150] M. J. Wessel et al., “Multifocal stimulation of the cerebro-cerebellar loop during the acquisition of a novel motor skill,” Sci. Rep., vol. 11, no. 1, p. 1756, Jan. 2021, doi: 10.1038/s41598-021-81154-2.

[151] M. J. Wessel, M. Zimerman, J. E. Timmermann, K. F. Heise, C. Gerloff, and F. C. Hummel, “Enhancing Consolidation of a New Temporal Motor Skill by Cerebellar Noninvasive Stimulation,” Cereb. Cortex, vol. 26, no. 4, pp. 1660–1667, Apr. 2016, doi: 10.1093/cercor/bhu335.

[152] M. A. Wilson, D. Greenwell, A. W. Meek, B. Poston, and Z. A. Riley, “Neuroenhancement of a dexterous motor task with anodal tDCS,” Brain Res., vol. 1790, p. 147993, Sep. 2022, doi: 10.1016/j.brainres.2022.147993.

[153] M. A. Wilson, D. Greenwell, A. W. Meek, B. Poston, and Z. A. Riley, “Neuroenhancement of a dexterous motor task with anodal tDCS,” Brain Res., vol. 1790, p. 147993, Sep. 2022, doi: 10.1016/j.brainres.2022.147993.

[154] R. E. Shimizu, A. D. Wu, J. K. Samra, and B. J. Knowlton, “The impact of cerebellar transcranial direct current stimulation (tDCS) on learning fine-motor sequences,” Philos. Trans. R. Soc. B Biol. Sci., vol. 372, no. 1711, p. 20160050, Jan. 2017, doi: 10.1098/rstb.2016.0050.

[155] N. Mizuguchi, T. Katayama, and K. Kanosue, “The Effect of Cerebellar Transcranial Direct Current Stimulation on A Throwing Task Depends on Individual Level of Task Performance,” Neuroscience, vol. 371, pp. 119–125, Feb. 2018, doi: 10.1016/j.neuroscience.2017.11.048.

[156] S. Miyaguchi et al., “Effects on motor learning of transcranial alternating current stimulation applied over the primary motor cortex and cerebellar hemisphere,” J. Clin. Neurosci., vol. 78, pp. 296–300, Aug. 2020, doi: 10.1016/j.jocn.2020.05.024.

[157] J. Reis et al., “Noninvasive cortical stimulation enhances motor skill acquisition over multiple days through an effect on consolidation,” Proc. Natl. Acad. Sci., vol. 106, no. 5, pp. 1590–1595, Feb. 2009, doi: 10.1073/pnas.0805413106.

