## Supplementary material for "Effects of concurrent transcranial direct and alternating current stimulation on human motor skill learning: a systematic review and Bayesian meta-analysis"

**S | Supplementary**

The supplementary material provides additional details supporting the main findings of the study. It includes: (1) a graphical overview of the included studies; (2) a risk of bias assessment; (3) quantitative data extracted for both online and offline learning phases; (4) detailed study characteristics (Tables S4–S7); and (5) results from the Bayesian prevalence analysis.

***S1 - Graphical overview of the included studies***

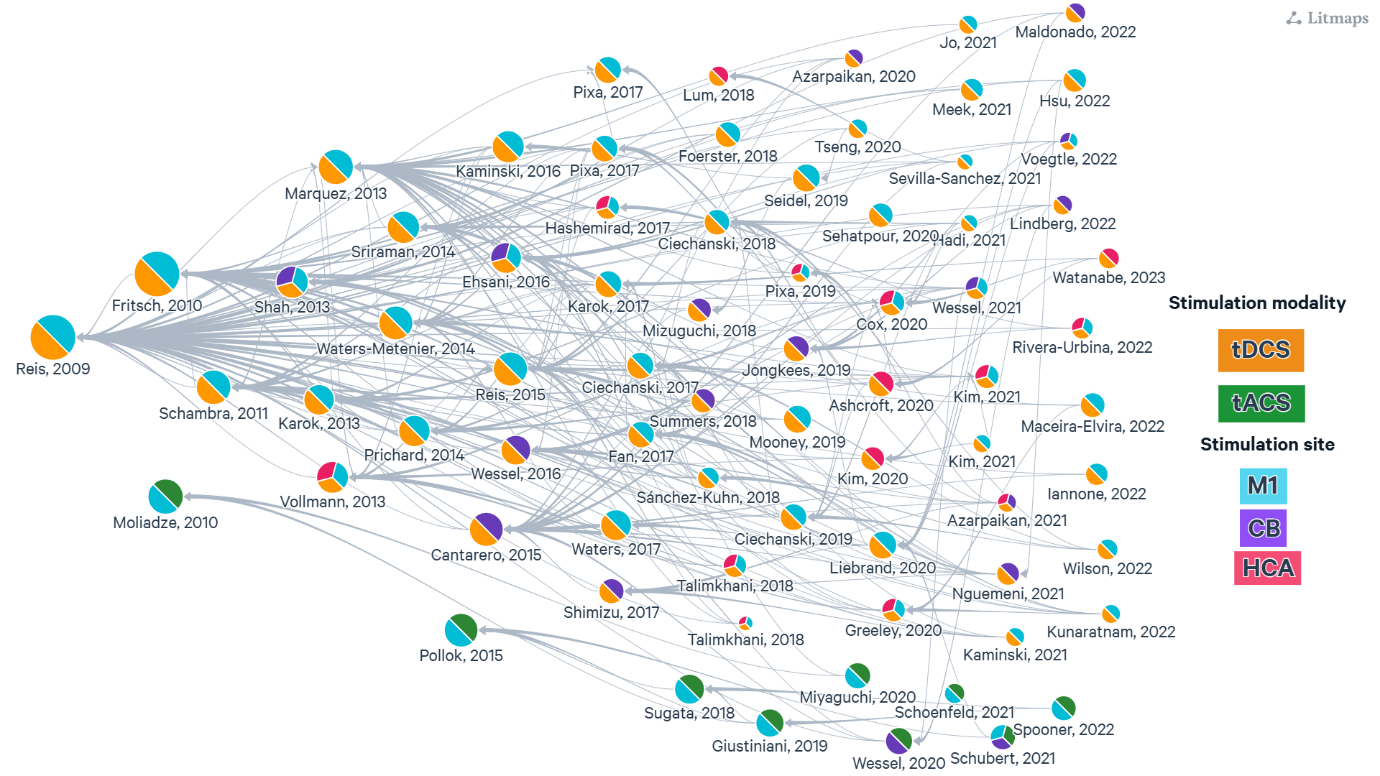

*Figure S1:* *Graphical overview of the included studies. Orange nodes represent studies investigating the effects of tDCS on motor learning, while green nodes represent tACS studies. Node colors (blue, purple, red) indicate the targeted brain regions: primary motor cortex (M1), cerebellum (CB), or other higher cortical areas (HCA), respectively. Grey lines and arrows indicate citation links between studies, and circle size reflects the number of times a study has been cited. Figure created via litmaps.com*

***S2 – Quality assessment***

We assessed the methodological quality of included trials using the Cochrane Risk of Bias tool. Overall, reporting was frequently insufficient, leading to many domains being rated as “unclear.” Below is a summary of our findings:

1. **Sequence Generation**
   - **Unclear risk:** 45% of studies did not describe the method used to generate the random allocation sequence (e.g., computer‐randomization, random number tables).
   - **Low risk:** 55% of trials explicitly detailed an appropriate randomization procedure.
2. **Allocation Concealment**
   - **Unclear risk:** 73% of studies failed to report how allocation was concealed from investigators prior to participant enrollment (e.g., sealed opaque envelopes, centralized randomization).
   - **Low risk:** 27% provided clear descriptions of adequate concealment, minimizing selection bias.
3. **Blinding of Participants and Personnel**
   - **Unclear risk:** 53% did not specify whether participants and trial personnel were blinded to group assignment or the methods used to achieve blinding (e.g., identical sham devices).
   - **Low risk:** 47% clearly described effective blinding procedures.
4. **Blinding of Outcome Assessment**
   - **Unclear risk:** 63% lacked sufficient detail on whether outcome assessors were blinded, or how blinding was maintained for subjective endpoints.
   - **Low risk:** 37% explicitly noted assessor blinding, reducing detection bias.
5. **Incomplete Outcome Data**
   - **Unclear risk:** 48% did not adequately report attrition rates or account for all randomized participants in the analysis (e.g., missing data, dropouts).
   - **Low risk:** 52% provided comprehensive information on withdrawals and used appropriate methods (e.g., intention‐to‐treat) to handle missing data.
6. **Selective Reporting**
   - **Low risk:** 95% of trials reported all prespecified outcomes with sufficient detail, and there was no evidence of outcome‐reporting bias.
   - **Unclear/high risk:** 5% had minor discrepancies between protocols and published results, but these did not materially affect the synthesis.
7. **Other Sources of Bias**
   - Across all studies, there were no consistent concerns regarding baseline comparability or funding-related conflicts of interest that would systematically compromise validity.

Most studies were judged at low risk of bias for selective reporting and were adequately described for randomization, blinding, and handling of missing data. However, substantial gaps in reporting methods for sequence generation, allocation concealment, and blinding procedures resulted in a predominance of “unclear” judgments.

***S3 – Quantitative data extraction for online and offline learning periods***

**********************************************************************************

Information data extraction (Online- & offline learning)

**********************************************************************************

###TDCS###

| *Ashcroft et al.,2020* | -Online: Baseline vs IMR |
| --- | --- |
| *Azarpaikan et al.,2020* | -Online: Baseline vs IMR : Day1 + day2  -Offline: IMR vs 24R-72R : Day3-2 + day4-3 + day8-4 |
| *Azarpaikan et al.,2021* | -Online: Baseline vs IMR : Day1 + day2  *-*Offline: IMR vs 24R |
| *Cantarero et al.,2015* | -Online: Baseline vs IMR : Day1 + day2 + day3  -Offline: IMR vs 24R : Day2-1 + day3-2 |
| *Ciechansk et al.,2018* | -Online: Baseline vs IMR  -Offline: IMR vs 6W-R |
| *Ciechansk et al.,2017* | -Online: Baseline vs IMR |
| *Ciechansk et al.,2019* | -Online: Baseline vs IMR |
| *Cox et al.,2020* | -Online: Total learning: Baseline vs post-practice 3days. |
| *Ehsani et al.,2016* | -Online: Baseline vs IMR  *-*Offline: IMR vs 48R |
| *Fan et al.,2017* | -Online: Total learning: Baseline vs post-practice 5days. |
| *Foerster et al.,2018* | -Online: Baseline vs IMR  -Offline: IMR vs 24R |
| *Fritsch et al.,2010* | -Online: Total learning: Baseline vs post-practice 5days. |
| *Greeley et al.,2020* | -Not enough information to be included in the meta-analysis |
| *Hadi et al.,2021* | -Online: Baseline vs IMR |
| *Hashemirad et al.,2017* | -Online: Baseline vs IMR  -Offline: IMR vs 24R |
| *Hsu et al.,2022* | -Online: Baseline vs IMR  -Offline: IMR vs 1H-R |
| *Iannone et al.,2022* | -Online: Baseline vs IMR  -Offline: IMR vs 24R |
| *Jongkees et al.,2019* | -Online: Baseline vs IMR |
| *Jo et al.,2021* | -Online: Total learning: Baseline vs post-practice 5days. |
| *Kaminski et al.,2021* | -Online: Baseline vs IMR : Day1 + day2 + day3  -Offline: IMR vs 24R : Day2-1 + day3-2 |
| *Kaminski et al.,2016* | -Online: Baseline vs IMR  -Offline: IMR vs 24R |
| *Karok et al.,2013* | -Online: Baseline vs IMR |
| *Kunaratnam et al.,2022* | -Online: Baseline vs IMR  -Offline: IMR vs 24R |
| *Liebrand et al.,2020* | -Online: Baseline vs IMR |
| *Lindberg et al.,2022* | -Online: Baseline vs IMR : Day1 + day2 + day3  -Offline: IMR vs 24R : Day2-1 + day3-2 |
| *Lum et al.,2018* | -Online: Baseline vs IMR |
| *Pablo Maceira-Elvira et al.,2022* | -Online: Total learning: Baseline vs post-practice 5days. |
| *Maldonado et al.,2022* | -Online: Baseline vs IMR |
| *Saucedo Marquez et al.,2013* | -Online: Total learning: Baseline vs post-practice 3days.  -Offline: Post-test vs 7D-R |
| *Mooney et al.,2019* | -Online: Baseline vs IMR  -Offline: IMR vs 24R |
| *Nguemeni et al.,2021* | -Online: Baseline vs IMR  -Offline: IMR vs 24R |
| *Pixa et al.,2018* | -Online: Total learning: Baseline vs post-practice 3days.  -Offline: post-practice 3days vs 24R |
| *Pixa et al.,2017* | -Online: Total learning: Baseline vs post-practice 3days.  -Offline: post-practice 3days vs 24R |
| *Prichard et al.,2014* | -Online: Baseline vs IMR : Day1 + day2 + day3  -Offline: IMR vs 24R : Day2-1 + day3-2 |
| *Reis et al.,2015* | -Online: Baseline vs IMR : Day1 + day2 + day3  -Offline: IMR vs 24R : Day2-1 + day3-2 |
| *Reis et al.,2009* | -Online: Baseline vs IMR : Day1 + day2 + day3 + day4 + day5  -Offline: IMR vs 24R : Day2-1 + day3-2 + day4-3 + day5-4 |
| *Rivera‐Urbina et al.,2022* | -Online: Baseline vs IMR |
| *Sánchez-Kuhn et al.,2018* | -Online: Total learning: Baseline vs post-practice 3days.  *-*Offline: post-practice 3days vs 8D-R |
| *Schambra et al.,2011* | -Online: Total learning: Baseline vs post-practice 3days. |
| *Sevilla-Sanchez et al.,2021* | -Online: Total learning: Baseline vs post-practice 20days. |
| *Shimizu et al.,2016* | -Online: Baseline vs IMR |
| *Talimkhani et al.,2019* | -Online: Total learning: Baseline vs post-practice 3days.  -Offline: post-practice 3days vs 4W-R |
| *Talimkhani et al.,2018* | -Online: Total learning : Baseline vs post-practice 3days.  -Offline: post-practice 3days vs 1W-R |
| *Tseng et al.,2020* | -Online: Baseline vs IMR  -Offline: IMR vs 30m-R |
| *Waters-Metenier et al.,2014* | -Online: Baseline vs IMR : Day1 + day2 + day3 + day4  -Offline: IMR vs 24R : Day2-1 + day3-2 + day4-3 |
| *Shah et al.,2013* | -Online: Baseline vs IMR |
| *Seidel & Ragert,2019* | -Online: Baseline vs IMR |
| *Sriraman et al.,2014* | -Online: Baseline vs IMR  -Offline: IMR vs 24R |
| *Mizuguchi et al., 2018* | -Online: Baseline vs IMR |
| *Karok et al.,2017* | -Online: Baseline vs IMR : Day1 + day2 + day3  -Offline: IMR vs 24R : Day2-1 + day3-2 |
| *Kim et al.,2021* | -Online: Baseline vs IMR  -Offline: IMR vs 24R |
| *Kim et al.,2021* | -Online: Baseline vs IMR  -Offline: IMR vs 24R |
| *Kim et al.,2020* | -Online: Baseline vs IMR  -Offline: IMR vs 24R |
| *Meek et al.,2021* | -Online: Baseline vs IMR |
| *Pixa et al.,2017* | -Online: Total learning : Baseline vs post-practice 3days.  -Offline: post-practice 3days vs 1W-R |
| *Sehatpour et al.,2020* | -Online: Baseline vs IMR |
| *Summers et al.,2018* | -Online: Baseline vs IMR |
| *Voegtle et al.,2022* | -Online: Baseline vs IMR |
| *Vollmann et al.,2022* | -Online: Baseline vs IMR |
| *Watanabe et al.,2023* | -Online: Baseline vs IMR |
| *Waters et al.,2017* | -Online: Total learning : Baseline vs post-practice 4days.  -Offline: post-practice 4days vs 24h-R |
| *Wessel et al.,2021* | -Online: Baseline vs IMR : S1+ S2 + S3 + S4  -Offline: IMR vs 24R : S2-1 + S3-2 + S4-3 |
| *Wessel et al.,2016* | -Online: Baseline vs IMR  -Offline: IMR vs 24R |
| *Wilson et al.,2022* | -Online: Baseline vs IMR |

###TACS###

| Spooner & Wilson et al 2023 | -Online: Baseline vs IMR |
| --- | --- |
| Wessel et al., 2020 | -Online: Baseline vs IMR  -Offline: IMR vs 24R |
| Sugata et al., 2018 | -Online: Baseline vs IMR |
| Schubert et al., 2021 | -Online: Baseline vs IMR |
| Schoenfeld et al., 2021 | -Online: Baseline vs IMR |
| Pollok et al., 2015 | -Online: Baseline vs IMR |
| Moliadze et al., 2010 | -Online: Baseline vs IMR |
| Miyaguchi et al., 2020 | -Online: Baseline vs IMR  -Offline: IMR vs 24R |
| Giustiniania et al., 2019 | -Online: Baseline vs IMR |

***S4: tDCS Assemble Specifications.***

| **tDCS Assemble Specifications** | | | | | | | | | |
| --- | --- | --- | --- | --- | --- | --- | --- | --- | --- |
| **Task** | **Author & publish year** | **Study design** | **number of participants + range/mean (SD ±) age** | **Groups** | **Target Area (Region of interest, ROI)** | **Montage (A/C)** | **Electrode size (A/C)** | **Current intensity (mA)** | **Duration (min)** |
| SVIPT | Cantarero et al., 2015 | Double-blinded | 33 (a-tDCS: 23.18 ± 1.20; c-tDCS: 27.90 ± 2.56; sham: 23.82 ± 1.36) | a-tDCS/ sham | CB | CB-R/ ipsilateral buccinator | 25 cm2 | 2mA | 20 min |
| SVIPT | Fan et al., 2017 | Double-blinded Randomzed | 30 (22.1 ± 3.3) | a-tDCS/ sham | M1 | M1-L/ SO-R | 35 cm2 | 2mA | 20 min |
| SVIPT | Fritsch et al., 2010 | NA | 34 | a-tDCS/ sham | M1 (hand area) | M1-L/ SO-R | NA | 1mA | 20 min |
| SVIPT | Hashemirad et al., 2017 | Parallel single-blinded randomized | 48 (25,82 ± 6,14) | M1-L a-tDCS/ DLPFC/ PPC/ sham | M1/ DLPFC/ PPC | M1-L/ SO-R; DLPFC (F3)/ SO-R; PPC (P3)/ SO-R | 3 cm2/ 12 cm2 | 0.3mA | 20 min |
| SVIPT | Iannone et al., 2022 | Double-blinded randomized | 30 (Conventional: 24.3 ± 1.41; Focal: 24.5 ± 1.71; Sham: 23.7 ± 1.52) | Conventional / focal / sham | M1 hand area | M1-R/ SO-L; HD-tDCS 4x1 (M1/surrounding) | conventional: 25 cm2; focal: 3.14cm2 | 2mA | 20 min |
| SVIPT | Kunaratnam et al., 2022 | Randomized single-blinded | 52 (a-tDCS: 24.23 ± 4.73; Sham: 26.31 ± 4.31) | a-tDCS / sham | M1 hand area | C3/ SO-R | 25/25cm2 | 1mA | 20 min |
| SVIPT | Marquez et al., 2013 | Double-blinded cross-over design | 27 (23.9 ± 3.13) | a-tDCS/ sham | M1 arm area | FDI hotspot/ ipsilateral deltoid | 25 cm2/ 99 cm2 | 1mA | 20 min |
| SVIPT | Mooney et al., 2019 | Pseudorandomized double-blinded crossover design | 16 (19–34) | a-tDCS/ sham | M1 hand area | M1-R/ SO-L | 25 cm2 / 35 cm2 | 1mA | 20 min |
| SVIPT | Reis et al., 2015 | Randomized double-blinded | 115 (18-53) | a-tDCS/ sham | M1 hand area | M1-L/ SO-R | 16 cm2 | 1mA | 20 min |
| SVIPT | Reis et al., 2009 | Double-blinded | 36 (Sham: 30.8 ± 3.0; a-tDCS: 28.3 ± 2.2; c-tDCS: 28.3 ± 1.3) | a-tDCS/ c-tDCS/ sham | M1 hand area (FDI) | M1-L/ SO-R | 25 cm2 | 1mA | 20 min |
| SVIPT | Schambra et al., 2011 | Parallel fractional single-blinded | 87 (27.8 ± 0.6) | a-tDCS / sham | M1 hand area | M1-L or M1-R/ ipsilateral deltoid | 25 cm2 | 1mA | 20 min |
| SRTT | Ehsani et al., 2016 | Parallel, randomized double-blinded, sham-controlled | 59 (a-tDCS: 22.77 ± 1.32; M1 a-tDCS: 23.35 ± 0.81; sham: 23.44 ± 0.78) | M1 a-tDCS / CB a-tDCS / sham | M1 / CB | C3/ SO-R; CB-R/ ipsilateral deltoid | 25 cm2 | 2mA | 20 min |
| SRTT | Jongkees et al., 2019 | Randomized | 72 | CB a-tDCS / CB c-tDCS / sham | CB | CB-C/ bilaterally over the mastoids | 35 cm2 | 1mA | 20 min |
| SRTT | Liebrand et al., 2020 | Cross-over design | 24 (22.6) | M1 a-tDCS/ CB a-tDCS/ sham | M1/ CB | CB-R/ mandibula-R; FC3/ CP3 | 9 cm2 | 1mA | 20 min |
| SRTT | Lum et al., 2018 | Single-blinded randomized | 36 (a-tDCS: 22.8 ± 3.3; Sham: 23.3 ± 3.0) | a-tDCS / sham | Inferior frontal gyrus | IFG-L/ SO-R | 35 cm2 | 2mA | 16 min |
| SRTT | Rivera‐Urbina et al., 2022 | Mixed partial crossover randomized and counterbalanced sham-controlled single-blind | 48 (24 ± 8) | a-tDCS / sham | M1/ PPC | P3/ SO-R; C3/ SO-R | 20 cm2/ 35 cm2 | 0.5mA | 14 min |
| SRTT | Talimkhani et al., 2019 | Randomized single-blinded | 53 (Dual-a-tDCS: 27.90 ± 0.88; a-tDCS: 28.19 ± 1; Sham: (27.70 ± 0.88) | dual-a-tDCS/ a- tDCS/ sham | M1/ DLPFC | a-tDCS-dual: M1 + DLPFC/ SO-R; a-tDCS: M1-L/ SO-R. | 6 cm2/ 12 cm2 | 1mA | 20 min |
| SRTT | Talimkhani et al., 2018 | Randomized Clinical Trial single-blinded | 37 ( 19-35) | dual-a-tDCS/ sham | M1/ DLPFC | C3 + F3/ SO-R | 6 cm2 / 12 cm2 | 1mA | 20 min |
| DSB (Discrete Sequence Production task) (FFT) | Greeley et al., 2020 | Randomized single-blind design | 65 (20.5 ± 2.4) | a tDCS/ sham | M1/ PFC/ SMA | M1-L/ SO-R; PFC-L (F3)/SO-R; PFC-R (F4)/SO-L; SMA/ Fpz | PFC/M1: 25 cm2/ 25 cm2 & SMA; 25 cm2/ 35 cm2 | 2mA | 20 min |
| Sequential finger-tapping task (FTT) | Hsu et al., 2022 | Single-blinded randomized | 108 (24.6 ± 6.42) | a-tDCS/ sham | M1 hand area | P4, CP4, CP2, P2/ F4, F2, AF4, and Fz; c-tDCS reverse montage | 2.7 cm2 | Total current: 4 mA; 1 mA per electrode | 12 min |
| Finger-tapping task (FTT) (random) | Jo et al., 2021 | Single-center prospective randomized controlled trial | 38 (a-tDCS online: 27,58 ± 9,12); a-tDCS prior: 25,69 ± 4,13); Sham: 28,23 ± 12,54) | a-tDCS/ sham | M1 hand area | M1-R/ SO-L | 35cm2 | 2mA | 20 min |
| Explicit sequential finger-tapping task (FFT) | Karok et al., 2013 | Sham-controlled repeated-measures design | 20 ( 25.6 ± 4.5) | Conventional a-tDCS/ dual-tDCS (bihemispheric) / sham | M1 hand area | C4/ C3; C4/ SO-L (F3) | Conventional: 25 cm2/ 35 cm2; dual: 25 cm2/ 25 cm2 | 1.5mA | 10 min |
| FTT | Maceira-Elvira et al., 2022 | Double-blind placebo-controlled parallel design | 19 (24.4) | a-tDCS / sham | M1 hand area | FDI hotspot/ SO-R | 25 cm2 | 1 mA | 20 min |
| Explicit sequence learning task (SEQTAP)(FFT) | Maldonado et al., 2022 | Randomized single-blinded | 71 (22.03 ± 3.44) | CB a-tDCS/ sham | CB | CB-R/ Deltoid-R; Deltoid-R/ CB-R | 25 cm2 | 2mA | 20 min |
| Sequential finger-tapping task (FTT) | Nguemeni et al., 2021 | Randomized double-blinded | 112 (22.5 ± 2.2) | CB a-tDCS/ sham | CB | CB-L / ipsilateral buccinator | 25 cm2 | 2mA | 20 min |
| Sequential Finger Tapping Task (SEQTAP) | Sánchez-Kuhn et al., 2018 | NA | 40 (20.77 ± 3.50) | a-tDCS/ sham | M1 hand area | C4/ Contralateral trapeze | 20 cm2 | 2mA | 20 min |
| Typing task (FFT) | Sevilla-Sanchez et al., 2021 | Longitudinal randomized single-blinded | 60 (21 ± 2) | a-tDCS/ sham/ control | M1 hand area | C3/ SO-R | 25 cm2 | 1.5mA | 15 min |
| Sequential finger-tapping task | Shimizu et al., 2016 | NA | 99 (Ex1: 21.11 ± 3.91; Ex2: 20.32 ± 1.82) | a-tDCS / sham | CB | CB-R/ ipsilateral cheek | 35 cm2 | 2mA | 20 min |
| Sequential finger-tapping task | Waters-Metenier et al., 2014 | Randomized double-blinded | 52 (21.90 ± 0.40) | Bi-tDCS/ sham | M1 hand area | M1-R/ M1-L | 35 cm2 | 2mA | 25 min |
| SRTT | Seidel & Ragert, 2019 | Sham-controlled, double-blinded, cross-over design | 46 (13 Football players ( 24.00 ± 3.89), 12 handball players (22.50 ± 4.32) and 21 Non-trained (26.95 ± 3.43) | a-tDCS / sham | M1 leg area | M1/Fpz | 35 cm2 | 2 mA | 20 min |
| FTT | Kim et al., 2021 | Single-blinded | 64 (19-23) | PMd-L tDCS / PMd-R tDCS/ Sham | PMd | F3/ SO-R & F4/SO-L | 25/35 cm2 | 2 mA | 20 min |
| FTT | Kim et al., 2021 | Single-blinded | 54 | a-tDCS / sham | M1 hand area | C4/SO-L | 35 cm2 | 2 mA | 20 min |
| Discrete sequence production task | Kim et al., 2020 | Single-blinded | 69 | a-tDCS / sham | SMA-R | FC2/SO-M | 35 cm2 | 2 mA | 20 min |
| SRTT | Sehatpour et al., 2020 | NA | 17: aged 19.4–54.0 (mean = 35.2, SD ± 10.8) | a-tDCS / sham | M1 hand area | C5/Fp2 | 9 cm2 | 2 mA | 30 min |
| SRTT | Voegtle et al., 2022 | Randomized double-blinded between-subject design | 60: 19–34 (M = 26.20, SD= 3.32; 35 females) | CB a-tDCS/ M1 a-tDCS/ sham | CB & M1 hand area | CB-R/ ipsi-deltoid & C3/ ipsi-deltoid | 35 cm2 | 2 mA | 15 min |
| FTT | Waters et al., 2017 | Double-blinded | 64: (54.69% females; average age 22.84 ± 0.56 years) | sham/ unihemispheric/ bihemispheric/ RP bihemispheric tDCS | M1 hand area | M1-SO/ M1-M1 | 35 cm2 | 2 mA | 25 min |
| SFTT | Wessel et al., 2021 | Randomized, double-blind, sham controlled, parallel design | 40: (age 25.93 ± 3.47, 23 female). | Sham/ M1 / CB / M1+ CB | M1 & CB | M1/ SO & CB/ Buccinator | 25/25 cm2 | 1-2 mA | 20 min |
| Auditory tapping task | Wessel et al., 2016 | Double-blind, sham-controlled, parallel design | 31: (25.0 ± 2.7) | CB tDCS/ sham | CB | CB-R/ Buccinator-R | 25 cm2 | 2 mA | 20 min |
| Surgical task | Ashcroft et al., 2020 | Randomized sham-controlled double-blind parallel design | 40 (21) | a-tDCS/ sham | PFC | F3/ F4 | 35 cm2 | 2mA | 15 min |
| Purdue Pegboard Test (PPT) & Fundamentals of Laparoscopic Surgery (FLS) | Ciechansk et al., 2018 | Double-blind sham-controlled randomized trial | 39 (Sham: 24,7 ± 3,3; a-tDCS: 26,3 ± 4,1) | a-tDCS / sham | M1 | C3/ SO-R; C4/ SO-L | 25 cm2 | 1mA | 20 min |
| Purdue Pegboard Test (PPT) & Fundamentals of Laparoscopic Surgery (FLS) | Ciechansk et al., 2017 | Double-blind randomized sham-controlled single-center pilot trial | 22 (Sham: 24.6 ± 2.1; a-tDCS: 25.8 ± 3.0) | a-tDCS / sham | M1 | C3/ SO-R; C4/ SO-L | 25 cm2 | 1mA | 20 min |
| Purdue Pegboard Test (PPT) & Fundamentals of Laparoscopic Surgery (FLS) | Ciechansk et al., 2019 | Parallel-design randomized sham-controlled double-blind | 22 (sham: 25.5 ± 4.7; a-tDCS: 25.9 ± 3.6) | a-tDCS / sham | M1 | CP4/ F2, FC2, F6, AF4; CP3/F1, FC1, F5, AF3 | 1 cm (diameter) | 1mA | 20 minutes |
| FLS peg transfer task | Cox et al., 2020 | Double-blinded randomized, and sham-controlled | 60 ( 22.7 ± 4.8) | a-tDCS/ c-tDCS/ sham | M1/ SMA | C3/ C4; Cz/ Fpz | 15 cm2 | 2mA | 40 min (2x20min) |
| Unimanual laparoscopic peg-transfer task | Hadi et al., 2021 | Randomized double-blinded crossover | 15 (26.24 ± 2.44) | a-tDCS/ sham | M1 | C3/ Fp2 | 9 cm2 | 1mA | 20 min |
| PPT | Pixa et al., 2017 | Multiple-day, double-blind study | 31: (age M = 23.42; SD = 2.45; range = 20–29) | HD a-tDCS/ sham | M1 hand area | M1-L+M1-R/ FC5,T7,CP5,FC6,T8,CP6 | 3.14 cm2 | 1 mA | 15 min |
| PPT | Watanabe et al., 2023 | Randomized, double-blind, sham-controlled trial | 66 | DLPFC a-tDCS/ sham | DLPFC-L | F3/ Fp2 | 35 cm2 | 2 mA | 20 min |
| PPT | Wilson et al., 2022 | Randomized between-subjects, Sham controlled design | 40: ( 22.18 ± 3.73) | a-tDCS/ sham | M1 hand area | M1-R or L/Fp2 or Fp3 | 25 cm2 | 1 mA | 20 min |
| Bimanual coordination task (Vienna) | Azarpaikan et al., 2020 | Randomized single-blind, and sham-control parallel design | 64 | a-tDCS prior/ a-tDCS online/ a-tDCS post/ sham tDCS | CB | CB-R/ ipsilateral deltoid | 25 cm2 | 1.5mA | 15 min |
| Bimanual coordination task (Vienna) | Azarpaikan et al., 2021 | Double-blind randomiseret | 64 (24.36 ± 2.51) | PA-a-tDCS/ CB-a-tDCS/ sham-tDCS/ Control-nostim | CB/ PA | P4 or CB/ right arm (deltoid muscle) | 25cm2 | 1,5 mA | 15 min |
| Arc pointing task | Kaminski et al., 2021 | Double‐blinded sham‐controlled | 30 (27.7 ± 3.8) | a-tDCS / sham | M1 hand area | M1-R/ SO-R | 35 cm2/ 100cm2 | 1mA | 20 min |
| Finger force manipulandum | Lindberg et al., 2022 | Randomized double-blinded | 20 (24 ± 2.8) | CB a-tDCS / sham | CB | CB/ buccinator muscle | 35 cm2 | 2mA | 20 min |
| Bimanual coordination task (Vienna) | Pixa et al., 2018 | Randomized double-blinded | 27 (23.18 ± 2) | HD M1 a-tDCS/ HD PA a-tDCS/ sham | M1/ PA | HD-a-tDCS M1: C1 and C2 /FC5, T7, CP5, FC6, T8, and CP6; HD-a-tDCS PA: CPz and Pz/ PO3, PO4, CP5, and CP6 | (Ag/AgCl, 3.14cm2) | 1mA pr anode (in all 2mA) | 15 min |
| Tracing tasks (Handwriting) | Prichard et al., 2014 | Randomized double-blinded | 90 ( 25.7 ± 4.6) | tRNS/ a-tDCS/ sham | M1 | Bi: M1-R hotspot/ contralateral M1-L; Conventional: M1-R hotspot/ SO-L; tRNS: M1-R hotspot/ SO-L; tRNS: T6/SO-L | 16 cm2 | 1mA | 20 min |
| VMT | Foerster et al., 2018 | Randomized single-blind sham-controlled | 33 (25.81 ± 3.85) | a-tDCS/ sham | M1 (right leg TA muscle) | M1-L/ SO-R | 8 cm2/ 88 cm2 | 0.5mA | 15 min |
| Ankle tracking task | Shah et al., 2013 | Double-blinded randomized repeated measures design | 8: (5 males and 3 females; age range 18-26 years) | CB a-tDCS/ M1 a-tDCS/ sham | M1 leg area & CB | M1-R/ SO-L & CB-L/ buccinator muscle-L | 8/ 35 cm2 | 1 mA | 15 min |
| VMT | Sriraman et al., 2014 | Crossover design | 12: (4 males, 8 females, age range 22–32 years) | a-tDCS/ sham | M1 leg area | M1/ SO | 8/ 35 cm2 | 1 mA | 15 min |
| Tracking | Summers et al., 2018 | Double-blinded, randomized, pre-test/post-test study design | 12: (Mean ± SD: 28.8 ± 10.5, 8 male) | a-tDCS/ sham | CB | CB/Buccinator muscle | 70/ 35 cm2 | 2 mA | 2x15 min |
| Tracking | Vollmann et al., 2013 | NA | 48 ( 23 ± 3 years, 25 female and 23 male participants) | M1/ sham/ SMA/ preSMA | SMA & M1 | M1/SO & SMA/SO & preSMA/SO | 10.7/100 cm2 | 0.75 mA | 20 min |
| Tracking | Karok et al., 2017 | Sham-controlled, mixed design | 30 (mean age 27.0 years ± 5.4 SD) | a-tDCS/ dual-a-tDCS/ Sham | M1 hand area | M1 / SO & M1-R/ M1-L | 25/48 & 25/25 cm2 | 1.5 mA | 15 min |
| Stepping task | Tseng et al., 2020 | Randomized-controlled single-blinded | 20 (27.3 ± 4.1) | a-tDCS/ sham | M1 leg area | lateral to Cz/ SO | 35 cm2 | 2mA | 20 min |
| Dart | Mizuguchi et al., 2018 | Double-blind, counter-balanced cross-over design | 24 (23 ± 3) | a-tDCS/ sham | CB | CB-R/ Buccinator-R | 25 cm2 | 2 mA | 20 min |
| Dart | Meek et al., 2021 | Randomized between-subjects, Sham controlled design | 58 (23.3 ± 3.9 yrs) | a-tDCS/ sham | M1 hand area | M1/ SO | 25 cm2 | 1 mA | 20 min |
| Cup stacking | Pixa et al., 2017 | Randomized multiple-session double-blinded | 32 (24.25 ± 2.75) | HD atDCS / sham | M1 | C1 & C2/ FC5, T7, CP5, FC6, T8 & CP7 | Ag/AgCl, 3.14 cm3 | 1mA (2mA in all) | 16 min |
| Whole-body dynamic balancing task - DBT | Kaminski et al., 2016 | Randomized sham-controlled single-blinded parallel design | 24 (26.08 ± 3.19) | a-tDCS / sham | M1 leg area | M1 leg area/SO-R | 25 cm2/ 50cm2 | 1mA | 20 min |

*Table S4: tDCS Assemble Specifications.*

***S5: tDCS Procedures of motor skill learning and motor learning outcome.***

| **Procedure of Motor Skill Learning** | | | | | | | | |
| --- | --- | --- | --- | --- | --- | --- | --- | --- |
| **Task** | **Author & publish year** | **Body part** | **Practice trials (amount and time)** | **Training sessions (days)** | **Behavioral measure(s)** | **Testing (Baseline & retentions tests)** | **Online effects on motor learning (tDCS vs sham)** | **Offline effects on motor learning (tDCS vs sham)** |
| SVIPT | Cantarero et al., 2015 | Hand | 6 blocks of 30 trials per day | 3 | Speed-accuracy | Baseline, post-practice x 3 & 7d-R | CB a-tDCS: Performance ↑ (speed → & accuracy ↑) | CB a-tDCS: Performance ↓ |
| SVIPT | Fan et al., 2017 | Hand | 6 blocks of 40-30 trials per day | 5 | Speed-accuracy & SAF | Continuously | a-tDCS: SAF performance ↑ & skill performance → | NA |
| SVIPT | Fritsch et al., 2010 | Hand | 200 trials per day | 5 | Speed-accuracy | Continuously over 5 days | a-tDCS: Skill performance ↑ | NA |
| SVIPT | Hashemirad et al., 2017 | Hand | 8 blocks and 8 trials | 1 | Movement time, error rate, & skill | baseline, IMR & 24h-R | a-tDCS: No difference | a-tDCS: No difference |
| SVIPT | Iannone et al., 2022 | Hand | 6 blocks of 30 trials | 1 | Speed-accuracy | baseline & 24h-R | a-tDCS: No difference | Focal a-tDCS vs sham: Focal performance ↑: Focal a-tDCS vs conventional a-tDCS: No difference |
| SVIPT | Kunaratnam et al., 2022 | Hand | a-tDCS: 6 blocks of 40-30 trials (1 day)/ sham: 6 blocks of 40-30 trials (5 days) | a-tDCS: 1 / sham: 5 | Speed-accuracy | baseline, 24h-R & 5d-R | a-tDCS: Skill performance day 1 ↑ | 24h-R: No difference |
| SVIPT | Marquez et al., 2013 | Hand | SEQTAP: 20 blocks of 40 sec; SVIPT: 7 blocks of 2 min | 3 for each task | Speed-accuracy | Baseline, 3d-R, & 1w-R | a-tDCS: SEQTAP performance ↑ (correct seq/mean response time) | 1w-R: a-tDCS SVIPT performance ↑ (error rate ↓/ duration ↓) |
| SVIPT | Mooney et al., 2019 | Wrist | 9 blocks of 12 trials | 1 | Speed-accuracy function (SAF) | Baseline, IMR, 24h-R, & 7d-R | No difference | No difference |
| SVIPT | Reis et al., 2015 | Hand | 5 blocks of 40-30 trials per day (480 in all) | 3 | Speed-accuracy | Baseline, 15min-R, 3h-R, 6h-R & 24h-R | No difference | Group 15min-R: 24h-R performance offline ↑; group 3h-R & 6h-R offline performance ↑; a-tDCS-after group: performance → |
| SVIPT | Reis et al., 2009 | Hand | 6 blocks of 40-30 trials (200 trials) per day | 5 | Speed-accuracy | Baseline, continuously & 5 follow-up sessions. | a-tDCS: Performance ↑ | Between-day offline learning ↑; 85D-R: a-tDCS Performance maintained → |
| SVIPT | Schambra et al., 2011 | Hands | 6 blocks of 40-30 trials | 3 | Speed-accuracy | Baseline & post-practice (IMR) | a-tDCS: M1-L-group performance ↑ (Skill ↑ & error rate ↓) | NA |
| SRTT | Ehsani et al., 2016 | Hand | 8 blocks of 10 trials | 1 | RT & error | baseline, 35min-R (imr) & 48h -R | RT: No difference →; CB a-tDCS: Error ↓ | M1 a-tDCS: short-term offline RT & Error learning ↑; CB a-tDCS: short-term offline Error learning ↑; M1- & CB a-tDCS long-term offline RT & Error learning ↑. |
| SRTT | Jongkees et al., 2019 | Hand | 13 blocks of 12 trials | 1 | RT & accuracy | Baseline & 24h-R | CB a-tDCS: Performance ↓ (RT ↑) | 24h-R: CB a-tDCS performance ↓ (RT ↑) |
| SRTT | Liebrand et al., 2020 | Hand/fingers | 11 blocks of 120 trials | 1 | RT & error rate | Baseline & IMR | CB-R a-tDCS performance ↑ (RT ↓ & correct guessed button press ↑): M1 a-tDCS: performance → | NA |
| SRTT | Lum et al., 2018 | Hand/fingers | 8 blocks of 96 trials | 1 | RT | Baseline & 5min-R | IMR: a-tDCS performance of a complex task ↑. Simple task performance → | NA |
| SRTT | Rivera‐Urbina et al., 2022 | Hand/fingers | 8 blocks of 120 trials | 1 | RT & error rate | Continuously | PPC a-tDCS SRTT performance ↑ (reaction time ↓) | No difference |
| SRTT | Talimkhani et al., 2019 | Hands | 8 blocks of 10 trials | 3 | Speed & accuracy | Continuously & 4w-R | No difference | 4w-R: dual M1+DLPFC a-tDCS skill performance ↑ |
| SRTT | Talimkhani et al., 2018 | Hand | 8 blocks of 120 key-presses per day | 3 | Speed & accuracy | Continuously & 1w-R | No difference | 1w-R: Dual M1 & DLPFC a-tDCS performance ↑ |
| DSB (Discrete Sequence Production task) | Greeley et al., 2020 | Hand | Day 1 & 2: 6 blocks (96 trials per seq); Day 3: 2 blocks (32 trials per seq) | 3 | RT, number of (motor) chunks, & number of errors in the complex sequences. | Baseline, 3d-R & 1y-R | PFC a-tDCS day 1 performance ↓ (nr of chunks ↓); SMA a-tDCS RT ↓; M1 a-tDCS day 2 RT ↓; PFC c-tDCS: Performance ↓ (nr of chunks ↓ & RT ↑) | 1y-R: M1- & L-PFC a-tDCS performance → |
| Sequential finger-tapping task (FTT) | Hsu et al., 2022 | Hand | 36 trials | 1 | Correct sequences & tapping speed | Practice session & 1h-R | a-tDCS: Performance ↑ (correct seq. ↑ & tap speed ↓) | a-tDCS: Performance ↑ (correct seq. ↑) |
| Finger-tapping task (FTT) (random) | Jo et al., 2021 | Hand | 120 finger taps per day | 5 | Speed & Accuracy | Baseline & post-practice (IMR) | NA | FFT: No differences; Prior a-tDCS GPT performance ↑ |
| Explicit sequential finger-tapping task (FFT) | Karok et al., 2013 | Hand/fingers | 12 trials of 36 tappings | 1 | RT & accuracy | Baseline, IMR, & post-15min-R | Bi-a-tDCS: Performance ↑ (RT ↓ & accuracy →) | NA |
| FTT | Maceira-Elvira et al., 2022 | hands/fingers | 6 blocks of 90 sec | 5 | % correct sequence | Baseline, continuously, 10d-R & 60d-R | No difference | No difference |
| Explicit sequence learning task (SEQTAP)(FFT) | Maldonado et al., 2022 | Hand/fingers | 12 blocks of 6 trials | 1 | RT & accuracy | Continuously | a-tDCS: Performance ↓ (accuracy ↓) | NA |
| Sequential finger-tapping task (FTT) | Nguemeni et al., 2021 | fingers | 20 blocks of 40 sec | 1 | Correct sequences tapped during each block | Baseline & 24h-R | No difference | No difference |
| Sequential Finger Tapping Task (SEQTAP) | Sánchez-Kuhn et al., 2018 | Fingers | 40 blocks of 20 sec tap | 3 | Skill Index (correct sequence/mean response time pr 20sec trial) | Baseline, 20min-R & 8d-R | Non-musicians: a-tDCS performance ↑ (correct sequence/mean response time=SI scores ↑ ) & Musicians no difference | 8D-R: non-musicians a-tDCS performance maintained → |
| Typing task (FFT) | Sevilla-Sanchez et al., 2021 | Hands | 15 min typing | 20 | Speed & accuracy (typing speed & error typing) | Baseline, post 10 sessions & post 20 sessions | NA | Session 0-10: a-tDCS performance ↑ (Error rate ↓); Session 10-20: All groups performance ↓; SAF = NA - inter-individual difference |
| Sequential finger-tapping task | Shimizu et al., 2016 | Hands | 6 blocks of 144 seq | 1 | RT & errors | Continuously | No difference | NA |
| Sequential finger-tapping task | Waters-Metenier et al., 2014 | Hands | 384 trials | 4 | Speed (execution time/ET + RT) & error | Baseline, continuously during practice, 5d-R, 12d-R & 33d-R | Synergy task: tDCS-related ET and RT performance ↑ & error →; Untrained task configuration & untrained hand performance ↑; Sequence task: tDCS ET performance ↑ & error →; Untrained sequences & hand ET performance ↑ | Synergy task 4w-R: a-tDCS maintained RT, ET & accuracy performance →; Sequence task 4w-R: a-tDCS ER performance ↑; a-tDCS ET performance maintained → |
| SRTT | Seidel & Ragert, 2019 | Lower limb & hands | 16 trials | 1 | RT | Baseline & IMR | No difference | NA |
| FTT | Kim et al., 2021 | Hand | 189 trials | 1 | Completion time | Baseline, post-practice & 24h-R | PMd L&R a-tDCS group: Performance score ↑ (completion time ↓) | No difference |
| FTT | Kim et al., 2021 | Hand | 189 trials | 1 | Completion time | Baseline, post-practice & 24h-R | No difference | a-tDCS group: Performance score ↑ (completion time ↓) |
| Discrete sequence production task | Kim et al., 2020 | Hand | 189 trials | 1 | Completion time | Baseline, post-practice & 24h-R | No difference | a-tDCS group: Performance score ↑ (completion time ↓) |
| SRTT | Sehatpour et al., 2020 | Hand | 2 x 12 min | 1 | RT | Baseline & Post-practice | No difference | NA |
| SRTT | Voegtle et al., 2022 | Hand | 15 min | 1 | RT | Baseline & Post-practice | CB-tDCS: Performance ↓; M1-tDCS: No difference | NA |
| FTT | Waters et al., 2017 | Hands | 384 sequence executions | 4 | Completion time | Baseline & Post-practice | Conv, Bi-tDCS, Bi-RP-tDCS: Performance ↑ | Conv, Bi-tDCS, Bi-RP-tDCS: Performance ↑ |
| SFTT | Wessel et al., 2021 | Hand | 7 blocks of 90 sec each session | 4 | Accuracy | Baseline & Post-practice | M1 & M1+CB tDCS: Performance ↑ | No difference |
| Auditory tapping task | Wessel et al., 2016 | Hand | 7 blocks of 140 sec each session | 1 | ER | Baseline, post-practice & 24h-R | No difference | 24h-R: Performance ↑ |
| Surgical task | Ashcroft et al., 2020 | Hands | 9 repetitions | 1 | performance score (sum of errors & completion time) | Baseline & IMR | a-tDCS group: Performance score ↑ (error score ↓ & completion time →) | NA |
| Purdue Pegboard Test (PPT) & Fundamentals of Laparoscopic Surgery (FLS) | Ciechansk et al., 2018 | Hands | 8 blocks of 1 repetition of each task | 1 | Performance score (sum of errors & completion time) | Baseline, post-practice & 6W-R | a-tDCS: Performance ↑ (cutting score ↑ & GPT →) | 6W-R: No difference |
| Purdue Pegboard Test (PPT) & Fundamentals of Laparoscopic Surgery (FLS) | Ciechansk et al., 2017 | Hands | 8 blocks of 3 min tumor resection | 1 | Brain resected, resection effectiveness, duration of excessive forces (EF) applied, & resection efficiency | Baseline, post-practice & 6W-R | a-tDCS: Performance ↑ (Tumor resected ↑, duration of excessive forces ↓, resection efficiency ↑ & effectiveness of resection →) | 6W-R: Performance maintained → |
| Purdue Pegboard Test (PPT) & Fundamentals of Laparoscopic Surgery (FLS) | Ciechansk et al., 2019 | Hands | 8 blocks | 1 | Error score and completion time | Baseline & IMR, 6h-R | a-tDCS: Performance ↑ (pattern cutting ↑ & PPT →) | NA |
| FLS peg transfer task | Cox et al., 2020 | hands | 2 blocks per day of 20 min | 3 | Performance score (completed peg transfer error deduction) | Pre-test & post-test | bM1 a-tDCS: Performance ↑ (Object moved ↑ & error ↓) | NA |
| Unimanual laparoscopic peg-transfer task | Hadi et al., 2021 | Hands | 10 min | 1 | Performance score (error score and completion time) | Baseline & IMR | a-tDCS: No difference | NA |
| PPT | Pixa et al., 2017 | Hands | 3 trials per day | 3 | Peg score | Baseline, Post-practice & Follow-up | HD-tDCS: Performance right hand ↑. The other tasks (left hand, both hands, assembly): No difference | HD-tDCS: Performance right hand ↑. The other tasks (left hand, both hands, assembly): No difference |
| PPT | Watanabe et al., 2023 | Hand | 6 blocks of 60 sec | 1 | Speed-accuracy | Baseline & post-practice | DLPFC-tDCS: Performance ↑ | NA |
| PPT | Wilson et al., 2022 | Hand | 10 pegs + 30-sec rest for 20 min | 1 | Speed-accuracy | Baseline & post-practice | a-tDCS group: Performance score ↑ | NA |
| Bimanual coordination task (Vienna) | Azarpaikan et al., 2020 | Hands | 2 blocks of 10 trials | 2 | Error and movement time | Baseline, IMR, 24h-R, 48h-R,4d-R & 8d-R | Concurrently a-tDCS and motor practice & a-tDCS after practice: Performance ↑ (Error ↓ & movement time ↓) | a-tDCS after motor practice: Performance ↑ (Error ↓ & movement time ↓) |
| Bimanual coordination task (Vienna) | Azarpaikan et al., 2021 | Hands | 15 min as many runs as possible | 2 | Error and movement time | Baseline, IMR, 24h-R and 48h-R | PA a-tDCS: Speed ↑ & CB a-tDCS: Error ↓ | CB a-tDCS: Performance ↑ (error ↓ & mean duration ↓); PA a-tDCS: mean duration ↓ (Retention 1 & 2) |
| Arc pointing task | Kaminski et al., 2021 | Hand | 20 trials per day | 3 | Accuracy & movement time | Continuous through practice | a-tDCS: No difference | a-tDCS: No difference |
| Finger force manipulandum | Lindberg et al., 2022 | fingers | Multi-finger tapping (MFT): 208 trials; Finger force-tracking (FFT): 32 trials per day | 3 | RT & error taps | Baseline, IMR & 10d-R | CB a-tDCS: No difference | No difference |
| Bimanual coordination task (Vienna) | Pixa et al., 2018 | Hands | 12 trials per day | 3 | Movement time & error | Baseline & 5-7d-R | No difference | 7D-R: PA HD-a-tDCS performance ↓ (movement time ↑) |
| Tracing tasks (Handwriting) | Prichard et al., 2014 | Hand | 12 blocks of 15 trials per day | 2 | Accuracy | Continuously | Uni-, Bi-tDCS & tRNS Performance ↑ (accuracy ↑) | No difference |
| VMT | Foerster et al., 2018 | Leg | 45 trials | 1 | Movement time, RT & Accuracy | Baseline, IMR & 24h-R | No difference | a-tDCS: Performance 24h-R ↑ (movement time ↓ & accuracy ↑) |
| Ankle tracking task | Shah et al., 2013 | Lower limb | 15 min | 1 | Accuracy | Baseline, 10min-R, 30min-R & 60min-R | Improvements in Accuracy Index with cerebellar anodal (10.85%), and anodal M1 (8.34%) | NA |
| VMT | Sriraman et al., 2014 | Lower limb | 15 min | 1 | Accuracy | Baseline, 10min-R, 25min-R & 24h-R | a-tDCS-online improved accuracy compared to the other groups during practice and 10min-R and 25min-R | 24h-R: a-tDCS prior & online had higher accuracy compared to sham |
| Tracking | Summers et al., 2018 | Hand | 6 blocks | 1 | Accuracy | Baseline & post-practice | No difference | NA |
| Tracking | Vollmann et al., 2013 | Hand | 20 trials | 1 | Accuracy | Baseline & post-practice | M1 & SMA tDCS: Performance ↑ | NA |
| Tracking | Karok et al., 2017 | Hands | 4 Blocks per day | 3 | Skill, accuracy & movement time | Baseline, post-practice & 10d-R | a-tDCS & dual-tDCS: Performance score ↑ (PPT ↑ / Accuracy ↑ / Movement time ↑) | dual-tDCS: Performance score ↑ (PPT ↑ / Accuracy ↑ / Movement time ↑); a-tDCS: PPT performance ↑ |
| Stepping task | Tseng et al., 2020 | Lower limbs | 50 steps | 1 | RT, movement time & accuracy | Baseline, IMR, & 30min-R | No online effects | 30min-R: a-tDCS performance ↑ (RT ↓) & accuracy → |
| Dart | Mizuguchi et al., 2018 | Hand | 6 blocks of 25 throw | 1 | Accuracy | Baseline, post-practice & 24h-R | No difference | No difference |
| Dart | Meek et al., 2021 | Hand | 60-100 throw | 1 | Accuracy | Baseline & Post-practice IMR | No difference | NA |
| Cup stacking | Pixa et al., 2017 | Hands | 4 blocks of 6 trials per day | 3 | Speed | Baseline, 24-48h-R, & 5-7d-R | Bi-HD-a-tDCS performance ↑ (Speed ↓) | 1w-R: Performance maintained → |
| Whole-body dynamic balancing task - DBT | Kaminski et al., 2016 | Legs | 15 trials of 30 sec. | 2 | Time in balance & RMSE | Baseline & 24h-R | a-tDCS: DBT performance ↑ (time in balance ↑ and RMSE ↑) | a-tDCS: DBT skill consolidation → & a-tDCS learning rate day 2 ↑ |

*Table S5: Procedures of motor skill learning and motor learning outcome.*

| **tACS Assemble Specifications** | | | | | | | | | | |
| --- | --- | --- | --- | --- | --- | --- | --- | --- | --- | --- |
| **Task** | **Author & publish year** | **Study design** | **number of participants + range/mean (SD ±) age** | **Groups** | **Target Area (Region of interest, ROI)** | **Montage (A/C)** | **Electrode size (A/C)** | **Current intensity (mA)** | **Duration (min)** | **Frequencies applied** |
| Sequential tapping task | Spooner & Wilson et al 2023 | Double-blind randomized crossover design | 25 ( range: 21–32) | Low-, peak-, high-gamma frequency, and sham (± 10 to peak) | M1 hand area | C3/ C1, C5, FC3, CP3 | NA | 2 mA (peak to peak) | 20 min | 60-100Hz (gamma) |
| SVIPT | Wessel et al., 2020 | Double-blind, sham controlled, cross over design | 15 (26.20± 3.34) | 50Hz-tACS/ sham | CB-L | CB-L/ Buccinator muscle ipsi | 25 cm2 | 2 mA (peak to peak) | 20 min | 50Hz |
| Sequential tapping task | Sugata et al., 2018 | Randomized single blinded | 52 (32.7 ± 6.8 years; 22 females) | 10Hz, 20Hz, 70Hz/ sham | M1-L hand area | C3/SO-R | 35 cm2 | 1 mA (peak to peak) | 10 min | 10/20/70Hz |
| SRTT | Schubert et al., 2021 | single-blinded | 24 (mean age: 24.8 years, range 2031; 11 males) | 10Hz M1/ 10Hz CB/ sham | CB-R or M1-L | CB-R/mandibula & FC3/CP3 | 15 cm2 (ring) | 1 mA (peak to peak) | 20 min | 10Hz |
| Bimanual task | Schoenfeld et al., 2021 | double-blind sham-controlled study | 54 (28 females, mean age = 24.05 years, SD = 4.76 years) | Indiv. peak beta-tACS (In phase or out of phase) / sham | M1/ ipsilateral shoulder x2 | C3 & C4/ ipsilateral shoulder x2 | 35 cm2 | 2 mA (peak to peak) | 20 min | indiv. Beta |
| SRTT | Pollok et al., 2015 | sham-controlled, double-blind design | 13+13(extra control) (6 male, 7 female with an average age of 22.08 ±.71; mean ± standard error of the mean, SEM) | 10Hz, 20Hz, 35Hz & Sham | M1-L hand area | M1-L/SO-R | 35 cm2 | 1 mA (peak to peak) | avg 12min + 12 sec (±4.7 s) | 10, 20, 35Hz |
| SRTT | Moliadze et al., 2010 | Single-blinded randomized crossover design | 13 (age 25.9 ± 2.35 S.D. years, range: 23–30 years) | 80Hz, 140z, 250Hz & Sham | M1-L hand area | M1-L/SO-R | 16 cm2/ 84 cm2 | 1 mA (peak to peak) | 10 min | 80, 140, 280Hz |
| Visuomotor control task | Miyaguchi et al., 2020 | NA | 30 (14 females, 16 males; mean age: 21. 0 ± 0.36 years) | 70Hz/ sham | M1-R/CB-L | M1-R/ CB-L | 25 cm2 | 1 mA (peak to peak) | 8x1 min | 70Hz |
| SRTT | Giustiniania et al., 2019 | NA | 17 (mean age 24.5 ± 3.5 years) | 40Hz/1Hz/sham | M1-L | M1-L/SO-R | 25 cm2 | 2 mA (peak to peak) | 5.08 ± 1.13 min | 40Hz, 1Hz |

***S6: tACS Assemble Specifications.***

*Table S6: tACS Assemble Specifications.*

***S7: tACS Procedures of motor learning outcome.***

| **tACS Procedures of motor learning outcome** | | | | | | | | |
| --- | --- | --- | --- | --- | --- | --- | --- | --- |
| **Task** | **Author & publish year** | **Body part** | **Practice trials (amount and time)** | **Training sessions (days)** | **Behavioral measure(s)** | **Testing (Baseline & retentions tests)** | **Online effects on motor learning (tDCS vs sham)** | **Offline effects on motor learning (tDCS vs sham)** |
| Sequential tapping task | Spooner & Wilson et al 2023 | Right hand | 480 trials | 1 | RT, accuracy, & movement time | Continuously | Accuracy: Low ↑: RT: High ↑ ; MT: Low & peak ↑ | NA |
| SVIPT | Wessel et al., 2020 | Right hand | 9 blocks of 90 s | 1 | Error | Baseline, R24H & R10D | No difference | No difference |
| Sequential tapping task | Sugata et al., 2018 | Right hand | 12 blocks | 1 | RT | Pre and post tACS | 70Hz tACS: motor performance ↑ | NA |
| SRTT | Schubert et al., 2021 | Right hand | 8 blocks of 120 seq | 1 | RT & ER (learning index) | Continuously | RT 10Hz CB tACS: Performance ↓ | NA |
| Bimanual task | Schoenfeld et al., 2021 | Both hands | 40 min | 1 | MT & ER | Baseline & IMR | No difference | NA |
| SRTT | Pollok et al., 2015 | Right hand | 6 blocks of 5 trials | 1 | RT/learning index | Continuously | 10Hz & 20Hz tACS: RT motor performance ↑ | NA |
| SRTT | Moliadze et al., 2010 | Right hand | 8 blocks of 120 trials | 1 | RT & ER | Continuously | No difference | No difference |
| Visuomotor control task | Miyaguchi et al., 2020 | Left hand | 15 trials | 2 | ER | Baseline, IMR, & R24h | No difference | R24h: 70Hz tACS performance ↑ (less increase in ER) |
| SRTT | Giustiniania et al., 2019 | Right hand | 8 blocks of 6 trials | 1 | RT | Baseline & IMR | 40Hz-tACS: motor performance ↓ | No difference |

*Table S7: Procedures of motor skill learning and results from the application of tACS.*

**S8: *| Bayesian Prevalence Estimation***

To assess the population prevalence of statistically significant effects, we applied a Bayesian prevalence method, inspired by Ince et al. (2021). This approach estimates the proportion of the population, from which the sample of studies was drawn, that exhibits a statistically significant effect. Each unit in the analysis represents a study that conducted a statistical test within the Null-Hypothesis Statistical Testing (NHST) framework, yielding a binary outcome (positive or negative). The analysis assumes a beta prior distribution combined with a binomial likelihood function. A uniform prior was employed, reflecting the absence of prior information regarding the distribution of effects across the population. This uniform prior implies that all population prevalence values are considered equally likely a priori. This analysis focused on the prevalence of true positive results, defined as the proportion of studies in the population that reported a statistically significant effect under the given experimental conditions. Under the uniform prior, the maximum a posteriori (MAP) estimate of the prevalence proportion of true positives (number of studies showing an above-threshold effect out of *n* total studies included in the analysis) is equivalent to the maximum likelihood estimate. Furthermore, the 96% highest posterior density intervals (HPDIs) were calculated to quantify the uncertainty of the MAP estimate. It is important to note that HPDIs are not equivalent to frequentist confidence intervals and should not be interpreted as such. Instead, they provide a probabilistic range that reflects the most credible values of population prevalence, given the observed data (Ince et al., 2021; McElreath, 2020).

***Results:***

Figures S8.1 and S8.2 present the Bayesian posterior distributions for the study prevalence of true positive results in both online and offline learning, as well as across different tDCS applications. It is important to clarify that these estimates do not reflect the study prevalence of the true binary status of the tested effect, which means that the analysis does not directly measure how often the tested effect truly occurs in the population in each study. Rather, our analysis focused on the binarization of statistical outcomes derived from each study, which involved a range of statistical tests considered suitable for the particular study designs. These tests were chosen based on a subjective frequentist approach by each study's researchers.

***Online motor learning***

The results in Figure 8 represent strong evidence of a non-zero effect in many studies in the different domains, albeit approximately balanced across studies between positive and negative directions. Considering the inference at the study level, 39/88 was significant within the analysis of M1 (Fig. 8B). The Bayesian prevalence approach provides a full posterior distribution from which we can obtain the maximum a posteriori (MAP) estimate, together with measures of uncertainty and highest posterior density intervals (HPDIs). Figure 8A (orange) shows Bayesian posteriors, MAPs, and HPDIs. The MAP prevalence estimate was 0.414 (96% HPDI: [0.308 0.523]) for studies that used M1 as the target area. Given the data, the probability that the study prevalence is greater than 30.8 is higher than 96%. Based on this result, it is likely that more than 30.8% of the population would show a true positive effect if tested under the same conditions (i.e., online learning and M1 as the target area).

The Bayesian prevalence of tDCS application to target CB (Fig. 8A & 8C) also suggests evidence of a non-zero effect within studies, albeit approximately sleight overweight across studies' acceptance of the NULL hypothesis. With regard to inferences at the study level, 7/25 were significant. The MAP prevalence estimate was 0.24 (96% HPDI: [0.089 0.44]). Given the data, the probability that the study prevalence was greater than 0.089  was higher than 96%. Based on this result, it is unlikely that the population would show a true positive effect if tested under the same conditions.

The results for higher cortical areas represent evidence of a non-zero effect within studies but overweight across studies' acceptance of the NULL hypothesis. With regard to inference at the study level, 8/27 were significant. The MAP prevalence estimate was 0.259 (96% HPDI: [0.1 0.45]). Given the data, the probability that the study prevalence is greater than 0.1 is higher than 96%. Based on this result, it is unlikely that the population would show a true positive effect if tested under the same conditions.

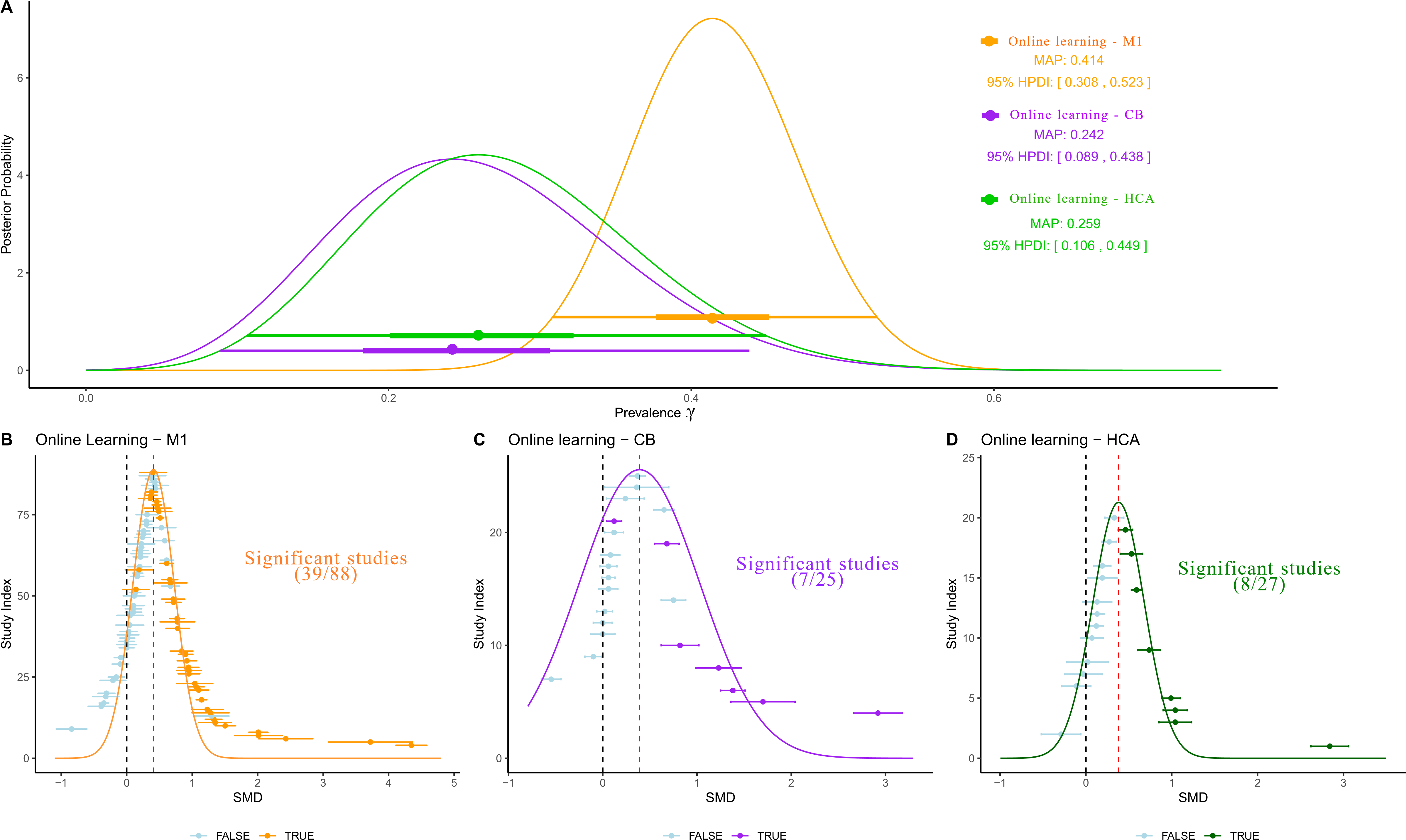

*Figure S8.1: Bayesian Posterior Prevalence for online motor learning. A, Bayesian posterior distributions of the study prevalence of true positive results for online learning and different tDCS applications. Circles show Bayesian maximum posteriori (MAP) estimates. Thick and thin horizontal lines indicate the 50% and 96% highest posterior density intervals (HPDIs), respectively. MAP (96%  HPDI) values are shown in the legend. B, C, D, Orange, purple, green, and blue indicate, respectively, exceeding or not exceeding a p=0.05, threshold for a given statistical test at the individual study level (on the within-study SMD). Orange = M1, Purple = CB, green = HCA.*

***Offline motor learning***

The results in Figure 9 show the Bayesian Prevalence estimate for the offline motor learning analysis. Considering the inference for tDCS application targeting M1 at the study level, 21/56 cases were significant. Figure 9A (orange) shows the Bayesian posteriors, MAPs, and HPDIs. The MAP prevalence estimate is 0.34 (96% HPDI: [0.218 0.48]). Given the data, the probability that the study prevalence is greater than 21.8 is higher than 96%. Based on this result, we would consider it likely that more than 21.8% of the population would show a true positive effect if tested under the same conditions.

The results for online learning and CB application (purple) demonstrate that 5/12 cases are significant. The MAP prevalence estimate was 0.38 (96% HPDI: [0.14 0.6]). Given the data, the probability that the study prevalence is greater than 14 is higher than 96%. Based on this result, we would consider it likely that more than 14% of the population would show a true positive effect if tested under the same conditions. The results for HCA (green) indicate that 1/13 of the cases are significant. The MAP prevalence estimate was 0.028 (96% HPDI: [0 0.27]).

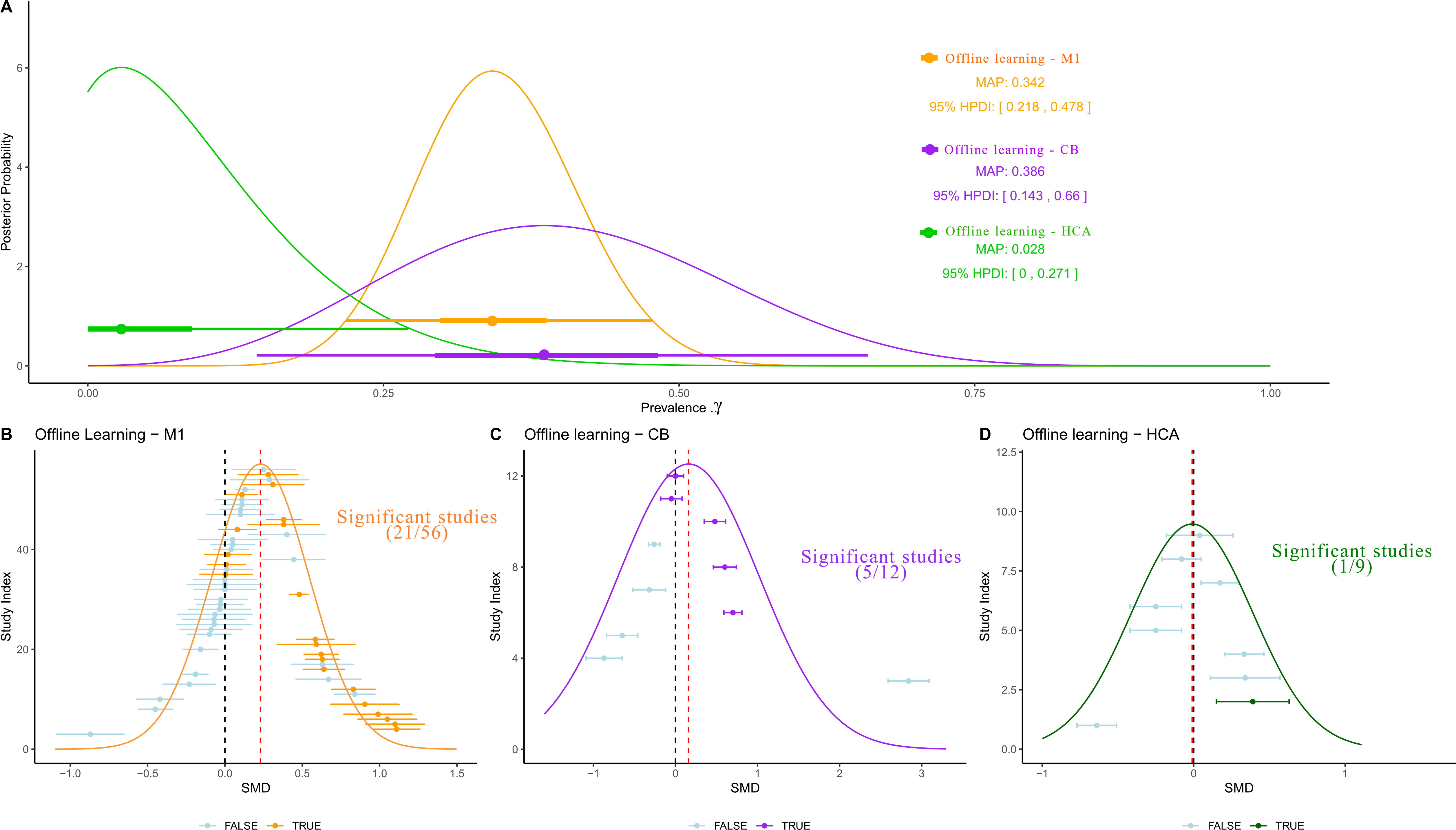

*Figure S8.2: Bayesian Posterior Prevalence for offline motor learning. A, Bayesian posterior distributions of study prevalence of true positive results for online learning and different tDCS applications. Circles show Bayesian maximum posteriori (MAP) estimates. Thick and thin horizontal lines indicate 50% and 96% highest posterior density intervals (HPDIs), respectively. MAP (96%  HPDI) values are shown in the legend. B, C, D, Orange, purple, or green and blue indicate, respectively, exceeding or not exceeding a p=0.05 threshold for a given statistical test at the individual study level (on the within-study SMD). Orange = M1, Purple = CB, and green = HCA.*
